# Suicide attempts: genetic and environmental risk factors, impact, and healthcare utilization—an analysis of nationwide data

**DOI:** 10.1101/2024.06.14.24308922

**Authors:** Thuy-Dung Nguyen, Kejia Hu, Karen Borges, Ralf Kuja-Halkola, Agnieszka Butwicka, Isabell Brikell, James J. Crowley, Zheng Chang, Brian M. D’Onofrio, Henrik Larsson, Paul Lichtenstein, Christian Rück, Cynthia M. Bulik, Patrick F. Sullivan, Fang Fang, Yi Lu

## Abstract

**Background:** Suicide is a major public health challenge, and a suicide attempt is an indicator of future mortality. This study provides a comprehensive analysis of initial suicide attempts.

**Methods:** Using Swedish national registers, we conducted a population-based cohort study of 3.7 million individuals followed from age 10 to a maximum age of 57. Suicide attempts were identified in hospital and death registers using ICD self-harm codes (intentional, with lethal methods, or leading to hospitalization or death). We investigated incidence, risk factors, outcomes, and familial aggregation, heritability, genetic correlations with psychiatric disorders, and healthcare visits in the month before and after initial suicide attempt.

**Findings:** The lifetime risk of suicide attempt in the study population was 4.6%, with greater risk in females and highest risk among ages 18-24. Overdose/poisoning were the most common methods. Prior history of psychiatric disorders, general medical diseases, and adverse life events were associated with increased risk of initial suicide attempt, while higher socioeconomic status was protective. Individuals with an initial suicide attempt were at substantially elevated risks of subsequent attempts (hazard ratio, HR, 23.4), suicide mortality (HR 16.4), and all-cause mortality (HR 7.3). One in ten families in Sweden had at least one individual who attempted suicide, and it tended to aggregate within families. The estimate of heritability was 42%, and genetic correlations of suicide attempts with psychiatric disorders ranged 0.48-0.85. At least 60% of those who made an initial suicide attempt had a healthcare contact in the month preceding the attempt.

**Interpretation:** The study provides comprehensive insights into suicidal behavior. Suicide attempts are major markers of poor mental health and risk for subsequent morbidity and mortality; indeed, they may carry the greatest mortal risk seen in clinical psychiatry. Our results underscore the need for systematic prevention efforts for individuals who have recently attempted suicide.

## INTRODUCTION

Globally, nine of every 100,000 people die by suicide, resulting in ∼700,000 deaths yearly.^1^ Many more attempt suicide. A 17-country study reported a lifetime prevalence of suicide attempts of 2.7%.^2^ In Sweden, the prevalence is higher (3.3%).^3^ Suicide attempts are among the strongest predictors of death by suicide.^4,5^ Effective suicide prevention requires thorough profiling of individuals who have attempted suicide to better understand the diverse characteristics of these people, associated risks and outcomes, and pathways to care.

Despite the corpus of research on suicide^4,6^, knowledge gaps remain. Previous studies have focused on death by suicide^7^ or suicidal behaviors in sub-populations.^8,9^ There are few large-scale population studies that specifically address suicide attempts (literature review in *Table S1*), leading to suboptimal understanding of suicide attempts in the general population. This is partly due to the scarcity of available data: death by suicide is often captured in mortality databases, while no official record for non-fatal suicide attempts exits.^5^ To thoroughly profile suicide attempts, a national population and longitudinal follow-up focus is needed.

Using data from nationwide registers in Sweden, we followed 3.7 million individuals for up to 47 years, identifying over 100,000 who had attempted suicide at least once. We focused on severe cases that required specialist care, aiming to deliver a comprehensive overview of medically serious suicide attempts in Sweden including incidence, risk factors, outcomes, familial patterns, and healthcare utilization.

## METHODS

This study was approved by the Swedish Ethical Review Authority (No. 2020-06540). Methodology is summarized here with full details in the *Supplement*.

### Nationwide Registers

This study used data from Swedish national registers, updated until 2020-12-31. To mitigate the impact of the COVID-19 pandemic, we included data up to 2019-12-31. We linked individual register data using Swedish personal identity numbers (assigned at birth or upon immigration): National Patient Register (NPR)^10^ for suicide attempts, psychiatric disorders, and general medical diseases using International Classification of Diseases (ICD) codes; Causes of Death Register (CDR)^11^ for deaths by suicide and other causes; total Population Register and Longitudinal Integrated Database for Health Insurance and Labor Market Studies (LISA) for sociodemographic factors and risk factors; National Prescribed Drug Register for dispensed medications; and familial information using the Multi-Generation and the Medical Birth Registers. Primary care data from public and private providers operating in Stockholm County were available from 2003-01-01 to 2013-12-31 (*Table S2*).

### Study population

The cohort comprised individuals who were born in Sweden from 1963-01-01 to 1998-12-31 (i.e., from ten years before the systematic capture of inpatient care in NPR in 1973 to a minimum age of 21 at the end of the follow-up). These individuals were followed from their tenth birthday to 2019-12-31. Suicide before age 10 is uncommon in Sweden^12^ and accidental ingestions by a child (98% of such presentations occur before age 10) is classified as “self-harm of undetermined intent”. For the genetic epidemiology analyses, we included all available relatives to ensure sufficient sample size but excluded: adoptees; or if both parents could not be confidently identified; or if younger than 10 on 2019-12-31 (*Methods S1*). We defined inclusive families as all individuals connected to a family by at least one of their parents, and nuclear families as parents and their offspring (*Methods S2*). For healthcare utilization, we used Stockholm County data for primary care, specialist care, and redeemed prescriptions (2005-08-01 to 2013-11-30).

### Definition of suicide attempt

Suicide is “the action or an act of taking one’s own life”^13^ and a suicide attempt is therefore the act of deliberate self-harm with the intention of causing death. Our main interest in this study is suicide attempts, especially initial attempts. We derived operational definitions of suicide attempts using the Swedish NPR and CDR, and followed prior Swedish studies^3,14^ with some modifications. We reviewed ICD codes for self-harm and/or suicidal behavior (ICD-CM codes are not used in Sweden) and consulted clinicians about Swedish diagnostic practices:

- ICD-10 codes X60-X84 capture suicidal actions as “intentional self-poisoning” or “intentional self-harm” and include purposely self-inflicted poisoning or injury
- ICD-10 codes Y10-Y34 capture events with insufficient data to distinguish accident, self-harm, and assault. Y87.0 and Y87.2 capture sequalae of self-harm of intentional or undetermined intent
- ICD-8/9 codes E950-E959 combine suicide attempts and other intentions (“suicide and self-inflicted poisoning/injury”).
- ICD-8/9 codes E980-E989 code for causes for self-harm but combine deliberate and other intentions (“undetermined whether accidentally or purposely inflicted”) and thus capture both suicide attempts and non-suicidal self-injury (e.g., shallow cutting to diminish strong negative emotions).

Following review and consultation, we defined <u>suicide attempt</u> as meeting one of three criteria (*Table S3*):

1. Patient records with ICD-10 codes X60-X84
2. Patient records with ICD codes E980-E989, Y10-Y34, Y87.0, Y87.2, E950-E959 with:
  a. A notably lethal method of self-harm (firearm, jumping from heights, motor vehicle crash, suffocation, or poisoning by domestic gas) and/or
  b. Led to inpatient care
3. Any deaths by self-harm or suicidal behavior (ICD X, Y, E codes)

We considered an individual who died in an initial suicide attempt—defined as death by suicide with no prior record of a treated suicide attempt or death during treatment for the initial attempt—as meeting the definition, as suicide death is the ultimate form of a medically serious suicide attempt.

To facilitate comparison with other studies^3,14^, we also defined a broader phenotype of <u>any self-harm including suicide attempts</u>, which included the ICD-8, −9, and −10 codes listed above from both NPR and CDR (*Table S3*). Figure S1 shows the numbers of people meeting the main definition of suicide attempt and the broader definition.

### Other variables

We investigated a range of <u>risk factors</u> for suicide attempts based on the literature.^15^ We derived >30 measures of socioeconomic status, comorbidities, adverse life events, and history of psychiatric disorders and general medical diseases from Swedish registers (*Table S4*). For <u>outcomes</u> of suicide attempts, we studied subsequent suicide attempts and mortality among those survived the first attempt. We concatenated all records with overlapping admission and discharge dates into one attempt. A subsequent attempt was then identified as the first record that did not overlap with the previous one. “Subsequent attempts” usually refers to the second attempts (except for *Figures S9, S10* and S13). We defined suicide mortality, non-suicide mortality, and all-cause mortality using the CDR records (*Tables S3-S4*).

### Study designs

We employed multiple designs (***Table 1***). A *<u>retrospective</u> <u>cohort</u> <u>design</u>* was used for suicide attempt incidence, genetic epidemiology, and healthcare utilization. We used all information since birth to capture medical conditions and demographic factors. For incident suicide attempt, we started follow-up at the age of 10 and followed individuals until the earliest occurrence of suicide attempt, death, emigration, or 2019-12-31 (Methods S1.3).

**Table 1.**
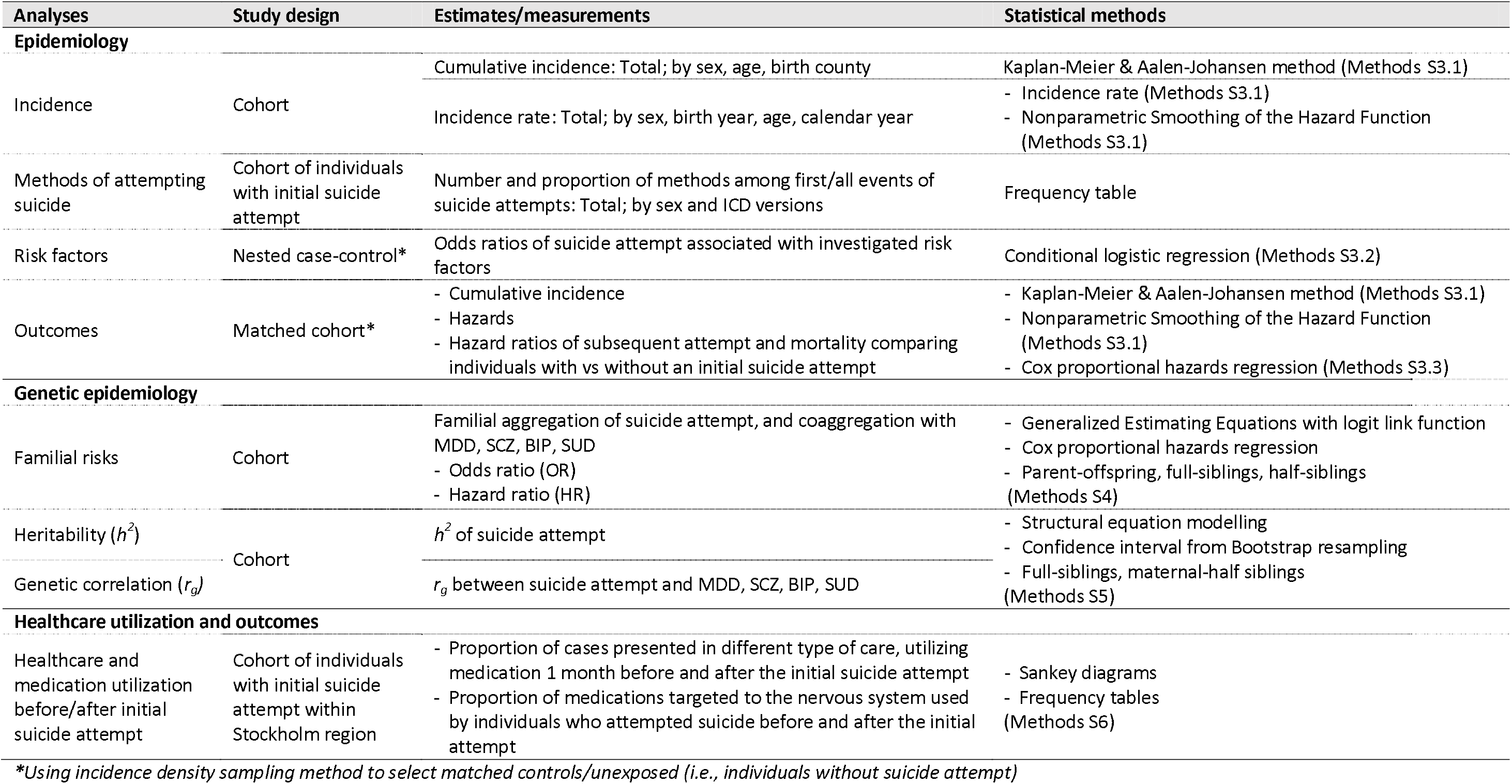
Overview of methods.

We applied a *<u>nested</u> <u>case-control</u> <u>design</u>* for risk factor analyses. For each individual who had a suicide attempt during the follow-up period (“cases”), we randomly selected five “matched controls” with the same sex and birth year and without a recorded suicide attempt by the index date (i.e., the date of first suicide attempt of the index case). Risk factors were defined from birth to the index date.

We applied a *<u>matched</u> <u>cohort</u> <u>design</u>* and followed the individuals from the date of an initial suicide attempt for four outcomes: subsequent suicide attempt, suicide mortality, non-suicide mortality, and all-cause mortality. Individuals with suicide attempts who survived the initial attempt (“exposed”) were matched with five randomly selected individuals with the same sex and birth year, and without a recorded suicide attempt by the index date (“unexposed”). Both groups were followed from the index date until each outcome of interest or being censored.

### Statistical methods

We estimated the cumulative incidence using the Kaplan-Meier method, and calculated incidence rates of suicide attempt for the full population and by subgroups (*Methods S3*.*1*). To study risk factors for suicide attempts, we applied conditional logistic regression to estimate the odds ratio (OR) associated with each risk factor. Model 1 conditioned on the matching clusters which, by design, controlled for sex and birth year. Model 2 adjusted Model 1 for socioeconomic status (education and income in the year before the initial suicide attempt; *Methods S3*.*2*).

For outcomes, Kaplan-Meier methods estimated cumulative incidence of a subsequent suicide attempt and mortality for individuals who attempted suicide and those who did not. We presented the cumulative incidence curve and the hazard curves, employing nonparametric smoothing of the hazard function.^16^ Competing risks were considered in sensitivity analyses.

We employed stratified Cox proportional hazards regression models to estimate cause-specific hazard ratios (HR) of subsequent attempt and mortality in relation to suicide attempt status, stratifying by matching clusters (Model 1) and adjusting for socioeconomic status (Model 2). We assessed effect modification by sex by including an interaction term and statistical significance using Wald tests. Data were censored at the end of follow-up, first emigration, or mortality due to causes other than the one of interest, and thus the estimated HRs were cause-specific (*Methods S3*.*3*).

To estimate the familial aggregation of suicide attempt and its coaggregation with four psychiatric disorders that are among the top risk factors for suicide^3^, we conducted analyses considering suicide attempt as binary and as time-to-event, adjusting for sex and birth year. For binary outcome, we fitted generalized estimating equations with logit link function. For time-to-event outcomes, we applied the Cox proportional hazards model with chronological age as the time scale and censoring for end-of-follow-up, emigration, and all-cause death. To account for family relatedness, we estimated robust standard errors by including each family as a cluster (*Methods S4*). With comparable results from binary and time-to-event outcomes (*Table S11*), subsequent quantitative genetic analyses considered suicide attempt as binary.

We applied structural equation models to estimate heritability (*h*^*2*^) of suicide attempt (univariate model) and genetic correlations (*r*_*g*_) between suicide attempt and psychiatric disorders (bivariate models), comparing full-siblings and maternal half-siblings. Model settings and assumptions were provided in *Methods S5*. We described path-to-care among individuals who were treated for the initial suicide attempt in Stockholm county using Sankey diagrams (*Method S6*).

## RESULTS

We present results for suicide attempts in the main text, and the majority of results for any self-harm including suicide attempts in the *Supplement*. We defined a birth cohort of 3,696,620 Sweden-born individuals (*Figure S2;* 48.6% female). The total population of Sweden in 2019 was 10.2 million. The age at the end of follow-up of the cohort ranged from 21 to 57 (mean 39.3, SD 10.5, median 39.4) (*Table S6*). In this cohort, 113,890 (3.1%) individuals had at least one suicide attempt (self-harm: 4.3%, *Figure S3*), among whom 55.2% were females. Among all individuals who attempted suicide, 8.8% died at the initial attempt (Characteristics in *Table S6*), and 30% had multiple suicide attempts during the follow-up (*Figure S3*).

### Epidemiology

#### Cumulative incidence

Among those who survived to age 57 (maximum age of our cohort), the cumulative incidence of specialist-treated suicide attempt was 4.58% (95% CI=4.54-4.62%). The percentage was higher among females (4.91% vs. 4.27% in males), and varied across Sweden (*Figure S4*). Females had higher cumulative incidence of attempting suicide than males after the age 13 (***Figure 1a***, *dashed lines*). Sensitivity analysis showed that the cumulative incidence of the initial suicide attempt was largely consistent when considering all-cause deaths as competing risk (*Figure S5*).

**Figure 1:**
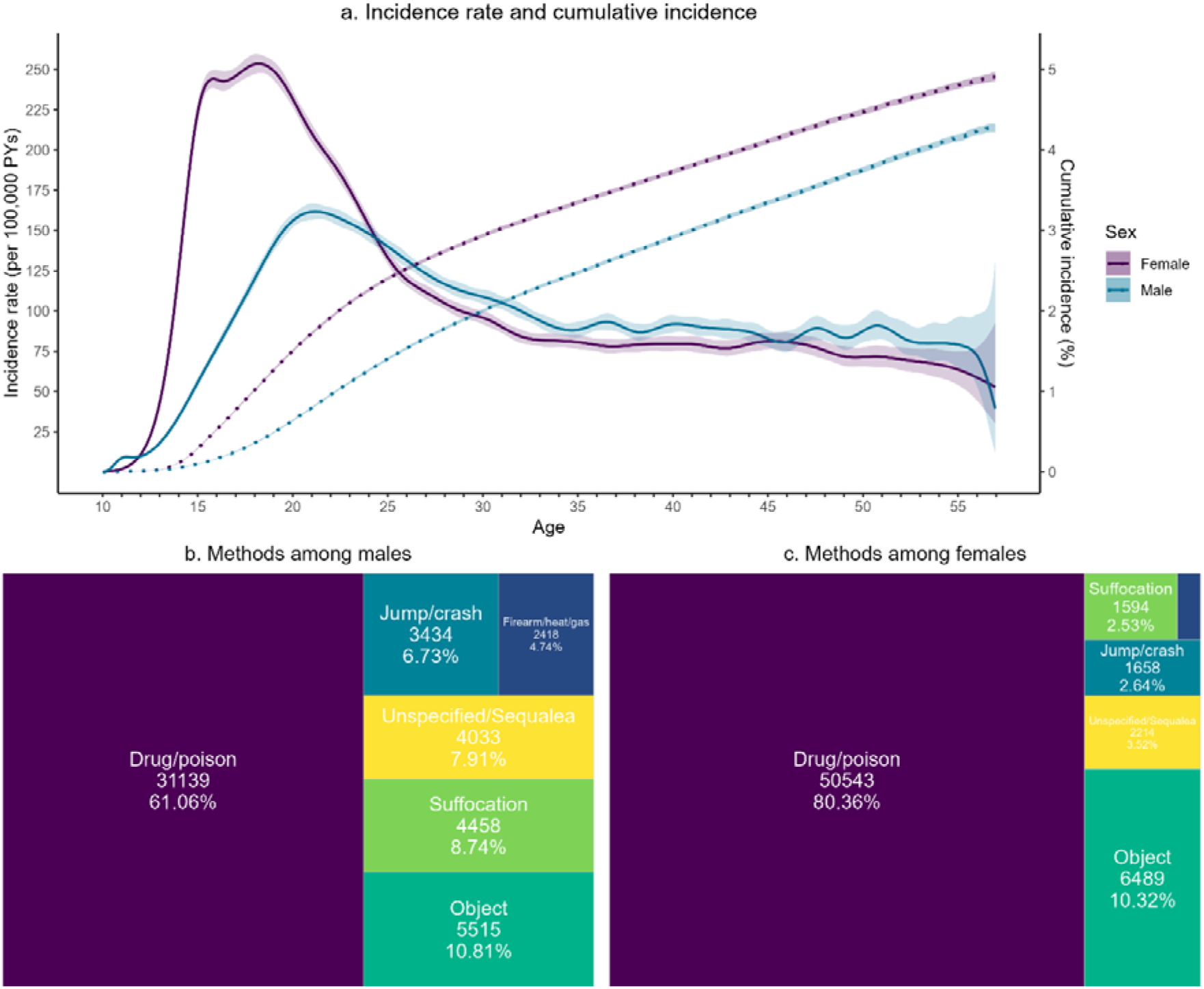
Incidence and methods of initial suicide attempts. **a) Incidence rate over age** (solid lines, y axis on the left) and **Cumulative incidence across age** (dashed line, y-axis on the right). Colors represent data separately by sexes. Shading regions show 95% confidence intervals of the estimates. Cumulative incidence was estimated using the Kaplan-Meier method. The lines show proportion (%) of individuals with a suicide attempt among individuals who were still at risk at corresponding age on the x-axis **b)-c) Methods used in the initial suicide attempt among males and females**. Data from the first record of each individual. Methods that accounted for <1% were collapsed to a single category of Firearm/heat/gas. The areas are proportional to the number of individuals within each sex. Definition of suicide methods is in Table S4.

#### Incidence rate

During birth cohort follow-up (102,391,191 person-years), there were 113,890 incident cases of suicide attempts. The overall incidence rate was 111.2 events per 100,000 person-years (95% CI=110.6-111.9; or 154.7 events for any self-harm). Females had higher incidence rates than males (127.0 CI=126.0-128.0 vs. 96.4 CI=95.6-97.3 events per 100,000 person-years).

The incidence rate for initial suicide attempts was lowest at age 10-12 but rose sharply thereafter to peak at around age 18 for females and age 20 for males, and then decreased with age. Females had a much higher incidence rate than males before age 24, after which the rates converged, and were slightly higher for males (***Figure 1a***, *solid lines*). Females also had a higher incidence rate in both inpatient and outpatient settings (*Figures S6*).

The overall incidence rate of initial suicide attempt was stable for adults throughout the year. For adolescents (notably for the 15-19 age group), the incidence rate tracked with the academic year with lower rates during summer/winter holidays and greater rates around the end of term in May and December (*Figure S7*).

#### Methods of suicide attempt

For both sexes, the most common method used in an initial suicide attempt was ingesting drugs or poison (males 61% and females 80%). Additional methods included: use of a blunt or sharp object; jumping or crashing; suffocation; firearm, heat, or gas; and unclear for 4-8% of attempts (***Figure 1b-c***). We observed a similar pattern of methods for any self-harm (*Figure S8*) as well as in all initial and subsequent attempts for an individual (*Figures S9, S10*).

#### Risk factors for initial suicide attempt

We investigated 31 risk factors for suicide using data collected prior to an initial suicide attempt (Figure S11). All investigated risk factors—demographics, adverse life events, psychiatric disorders, and general medical diseases—were associated with suicide attempts (Figure 2, Table S7). The largest associations were for psychiatric disorders, especially major depressive disorder (MDD, OR=12.85, 95% CI=12.49-13.21). Having any of six psychiatric disorders was associated with a nearly 14-fold greater risk of attempting suicide (OR=13.82, CI=13.52-14.13). For general medical diseases, sleep disorders (also a symptom of MDD) had the greatest risk for initial suicide attempts (OR=3.22, CI=3.05-3.40) and a prior history of obesity, diabetes, chronic pain, or cardiovascular diseases showed ORs of 1.84-2.03. Among adverse life events, being assaulted or victimized had the largest effect (OR=4.38, CI=4.23-4.53). Higher personal and parental socio-economic status were associated with lower risk of suicide attempts. For instance, those with higher educational levels had 60-75% lower odds of attempting suicide (versus those with <9 years of education). Compared to individual socioeconomic status, parental education and income had comparatively weaker protective effects. Compared to unmarried individuals, those who were married had lower odds of attempting suicide, whereas individuals who were divorced or widowed showed higher odds of attempting suicide.

**Figure 2:**
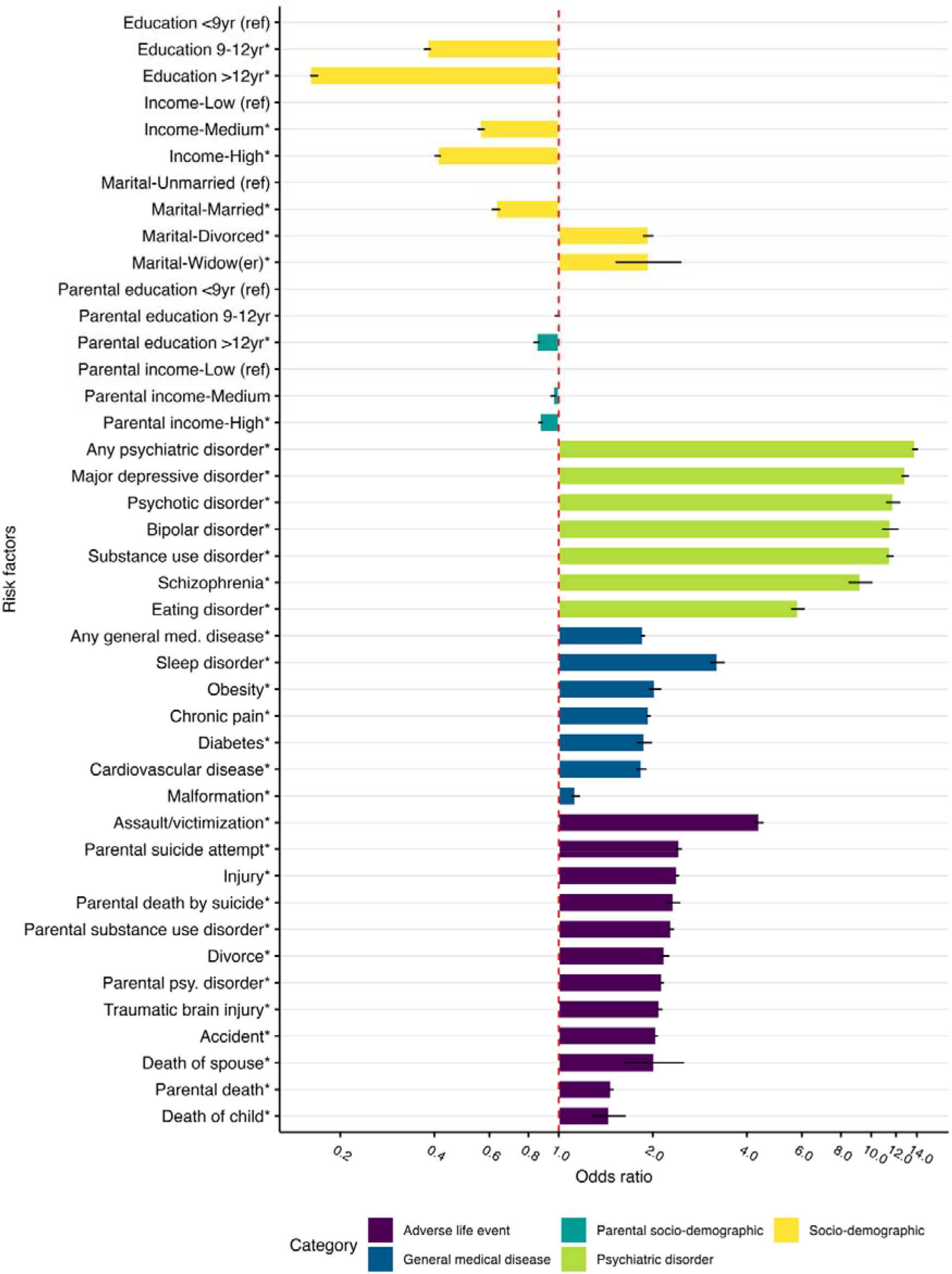
Odds ratios of suicide attempts associated with risk factors. Results from the conditional logistic regression, conditioning on matching clusters (i.e., controlled for sex and birthyear by design) and further adjusted for education and income. Bars show odds ratios of having a suicide attempt. Error bars show 95% confidence intervals. Asterisks (*) indicate OR was statistically significantly different from 1 (Bonferroni-corrected p-value ≤ 3.88*10^-4^)

The sex-stratified analyses showed similar patterns to the results from the whole population but with some notable differences between sexes. For instance, substance use disorders showed a stronger association among males (OR=12.05; 95% CI=11.62-12.50) than females (OR=10.86; CI=10.43-11.30) whereas eating disorders had a stronger effect among females (OR=6.04; CI=5.73-6.37) compared with males (OR=3.54; CI=2.96-4.23) (*Table S7*). For any self-harm, we found consistent but attenuated effects of these risk factors (*Table S8*).

#### Outcomes

During the follow-up, the HR of having a subsequent suicide attempt or death was higher among those with an initial suicide attempt compared to those without (***Figure 3***; sex-specific in *Figure S12*). The HRs were extremely high in the weeks or months immediately after the initial attempt but decreased rapidly after one year. For those who attempted suicide, the incidence rates of subsequent suicide attempt, suicide mortality, non-suicide mortality and all-cause mortality were: 3182.0, 264.6, 273.6, and 552.2 events per 100,000 person-years, respectively (*Table S9a*). The risk of making a subsequent attempt or death increased with the number of previous attempts (*Figures S13-14*).

**Figure 3:**
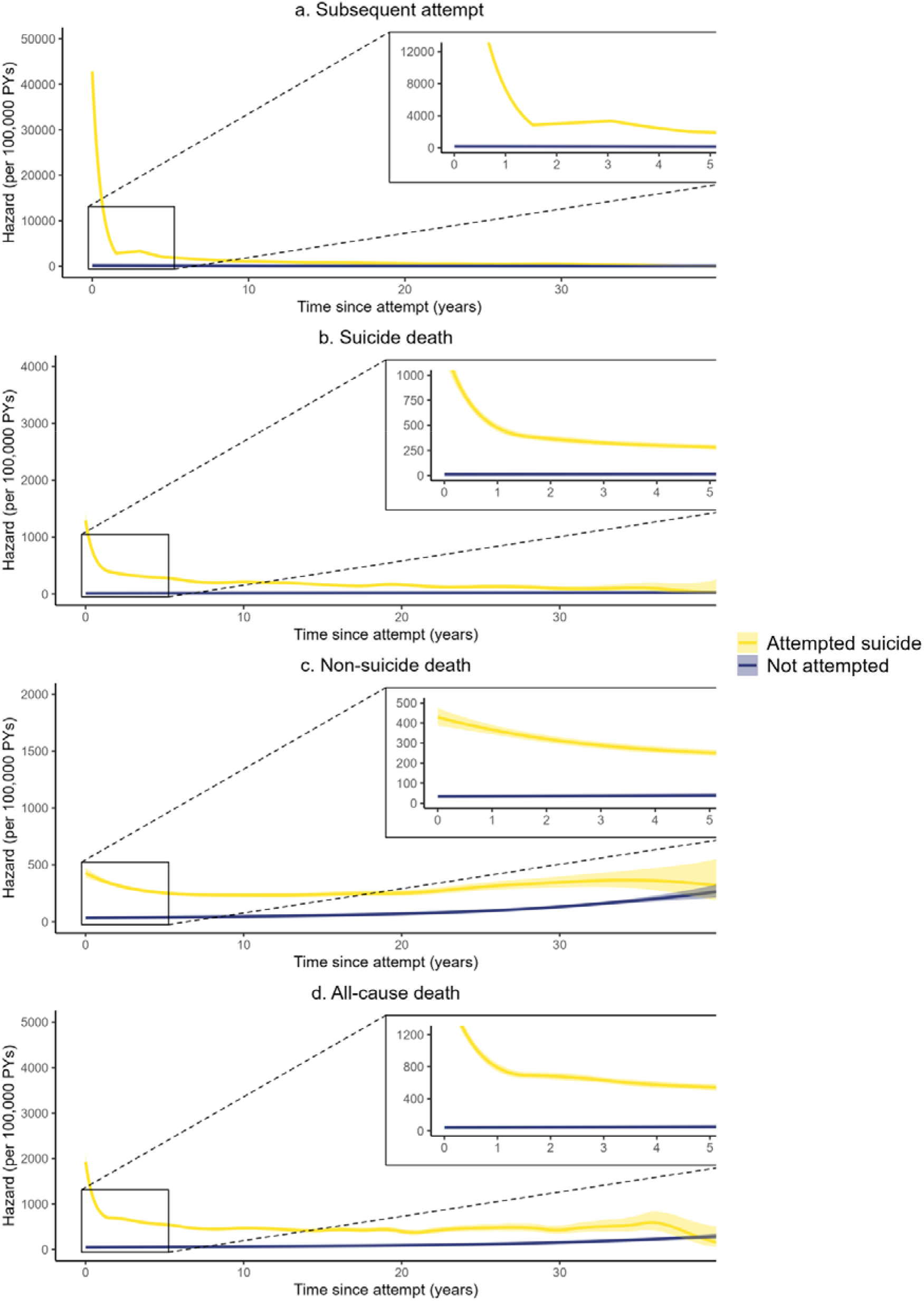
Hazards of subsequent suicide attempt and mortality. The lines show hazards (number of events per 100,000 person-years) of four outcomes among individuals who had and had not an initial suicide attempt from a matched cohort. Shaded areas represent 95% CI of the hazard.

Individuals who had an initial suicide attempt were at substantially elevated risk of making another attempt (HR=23.39, 95% CI=22.78-24.02), with higher risks in males (HR=24.71) than females (HR=22.66; exposure-sex interaction P=2.1×10^-3^ ) (Table S9a-b). Experiencing an initial suicide attempt was associated with a seven-fold increased risk of all-cause mortality (HR=7.31, CI=7.04-7.59). The increased risk was higher for death by suicide than death due to other causes (HR=16.40, CI=15.29-17.60, versus HR=4.41, CI=4.21-4.63) (*Table S9a*). However, males had higher risk increase in all-cause mortality (HR=8.39 in males vs 6.04 in females, interaction P=2.0×10^-17^), and non-suicide mortality (HR=5.48 in males vs 3.21 in females, interaction P=1.9×10^-28^). Males and females had similar risk increase in death by suicide (HR=15.51 in males vs. 17.82 in females, interaction P=0.06) (*Table S9b*). For self-harm, the HRs were lower, but the pattern was similar to suicide attempt (*Table S9c*).

When competing risks were considered, we still observed higher cumulative incidences of subsequent attempts and mortality among those with an initial suicide attempt compared to those without (*Figure S15*). For example, when considering non-suicide death as competing risk, the absolute risk of subsequent death by suicide within 1, 5, 10 years after the initial attempt was 0.7% vs. 0.008%, 2.1% vs. 0.06%, 3.2% vs. 0.1% for individuals with and without suicide attempt in the matched cohort.

### Genetic epidemiology

Of the inclusive family groupings comprising this cohort (Methods), 10.02% had at least one family member who had attempted suicide (*Table S10*). Suicide attempts aggregated within families. Relatives of individuals with suicide attempts had elevated risks of attempting suicide, with the highest HR of 2.87 (95% CI=2.81-2.93) for mother-offspring pairs. First-degree relatives (parents-offspring and full-siblings) had higher risks compared to second-degree relatives, suggesting a complex genetic influence on risk of suicide attempt. Aggregation of suicide attempts among maternal half-siblings was higher than paternal half-siblings with non-overlapping CI, suggesting the influence of shared environment (in Sweden, maternal-half siblings are more likely to live together than paternal half-siblings^17^; ***Figure 4a***, *Table S11a*). Relatives of those who attempted suicide had significantly increased risk for MDD, schizophrenia, bipolar disorder, and substance use disorders (HR range 1.21-2.54) with higher HRs among the first-degree compared to second-degree relatives. For MDD and substance use disorders, the HR was higher for mother-offspring pairs compared with father-offspring pairs (***Figure 4b***, *Table S11a*).

**Figure 4:**
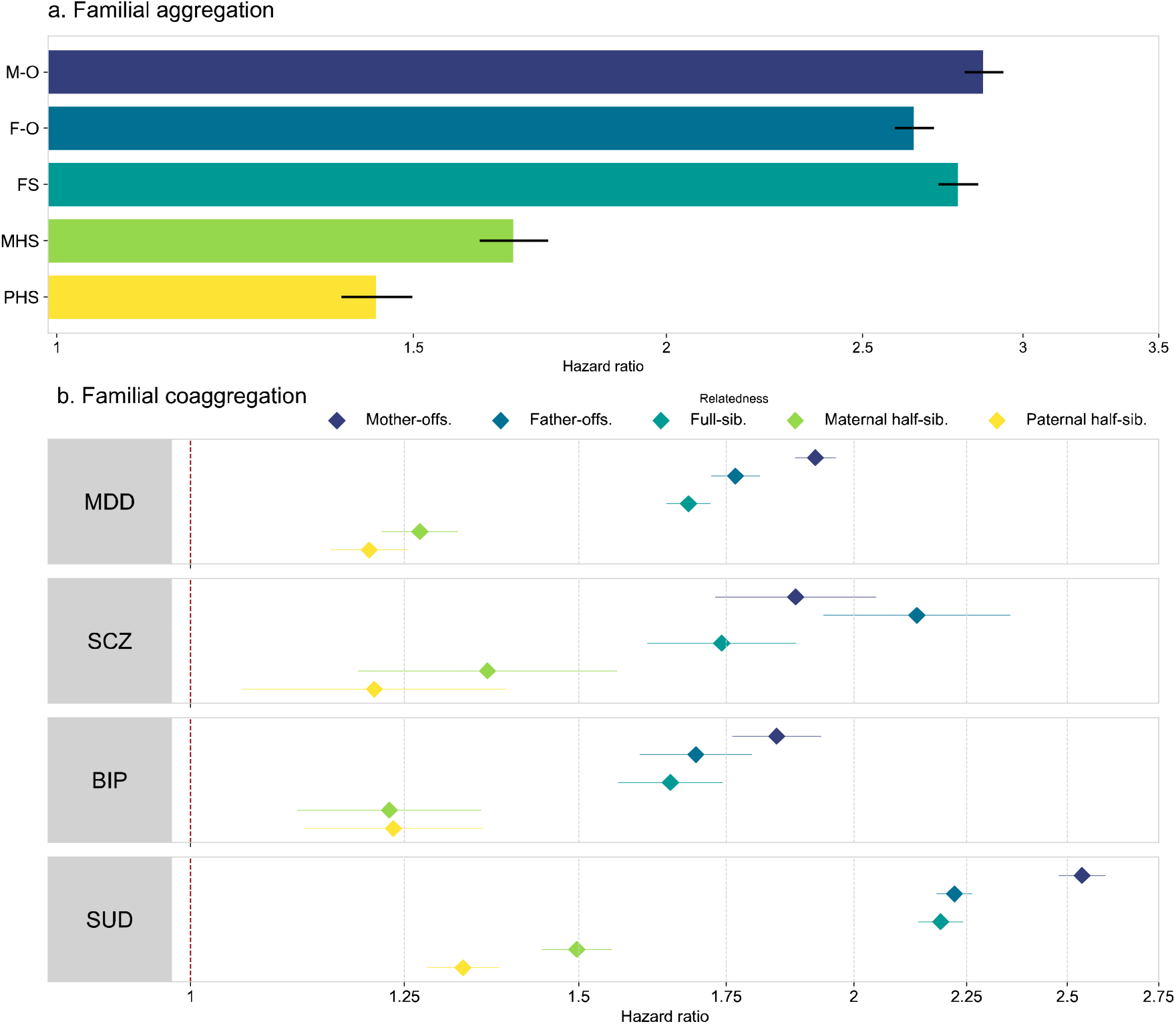
Familial aggregation and coaggregation of suicide attempts. **a. Familial aggregation** of suicide attempt; **b. Familial coaggregation** of suicide attempt with major depressive disorder (MDD), schizophrenia (SCZ), bipolar disorder (BIP) and substance use disorders (SUD) within family separately for types of relatives. Results from analyses of time-to-event outcome. Bars and dots show hazard ratio, error bars show 95% confidence interval for the estimates. Colors represent different types of relatedness (same color in plots a and b). M-O: Mother-offspring | F-O: Father-offspring | FS: Full-siblings | MHS: Maternal half-siblings | PHS: Paternal half-siblings.

The pedigree-heritability of suicide attempts was 41.9% (95% CI=35.9-48.2%). We observed a small but significant influence of shared environmental factors (3.6%, 0.8-6.4%). There were moderate to high pedigree genetic correlations with four psychiatric disorders (range 0.48-0.85) with the highest for substance use disorders (*r*_*g*_=0.85, 95% CI 0.83-0.96) (*Table S12a*). Estimates for self-harm were comparable (pedigree-heritability 42.3%; *Tables S11b*,*12b*).

### Healthcare utilization

To depict paths-to-care for individuals who attempted suicide, we extracted healthcare contact information for 3,646 individuals with an initial suicide attempt and for whom we also had primary care data in Stockholm County. For an initial suicide attempt, 7% were treated in primary care, 56% at outpatient specialist care, and 36% had an inpatient care. In the month preceding an initial suicide attempt, 60% of individuals had contact with healthcare: 46% visited a pharmacy to obtain prescription medications, 38% had outpatient specialist care visits, 11% were hospitalized, and 12% had primary care visits (***Figure 5***). For the month after an initial suicide attempt, we observed increase in visits made in each category, with the greatest increase in outpatient specialist care (18.2%). Within specialist care there was a higher increase in psychiatric visits compared to non-psychiatric (*Table S13*). Similarly, the use of psychotropic medications also increased (*Table S14*). Notably, 23% of those who attempted suicide had no healthcare contacts in the month after the event.

**Figure 5:**
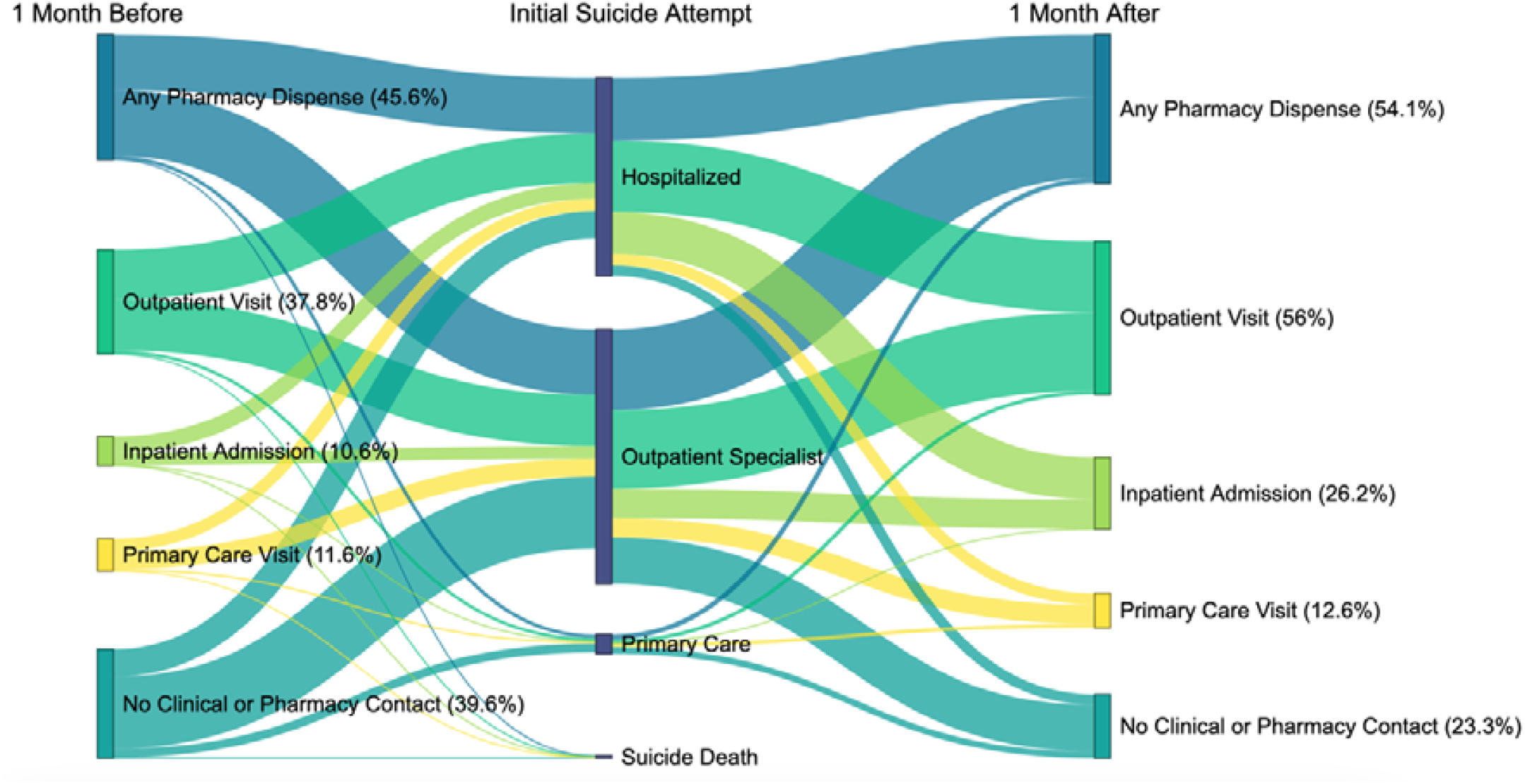
Healthcare utilization and outcome after initial suicide attempt. Data from 3646 individuals with an initial suicide attempt recorded in Stockholm region during 1/10/2005-30/9/2013 in both primary and specialist care. Node 1 describes clinical contacts and pharmacy visits within 30 days before the initial suicide attempt incidence. Node 2 describes the location where the initial suicide attempt presented. Node 3 describes clinical contacts and pharmacy visits within 30 days after the initial suicide attempt. Pharmacy visits included prescribed medications. Medication dispensed on the day of suicide attempt was considered 1 month after. Groups in node 1, 3 are not mutually exclusive. % presented in group labels of node 1 is among 3646 individuals. % presented in group labels of node 3 is among 3629 individuals who survived the initial attempt.

## DISCUSSION

Many studies have centered on death by suicide or suicidal behaviors in population segments (e.g., by ethnicity, sex, or age) or based on specific general medical or psychiatric diagnoses. There remains a critical knowledge gap in describing those who attempt suicide for the first time in a population. In this Swedish national study, we provide comprehensive insights into those who make an initial suicide attempts (summarized in ***Table 2***). Our findings emphasize the importance of focusing on this group for suicide prevention.

**Table 2.** Summary of key findings.

| **Result** | **Key findings from a Swedish birth cohort (1963-1998)** |
| --- | --- |
| Population risk of suicide attempt | • 3.1% (113,890 out of 3,696,620 individuals) made a suicide attempt<br>• Cumulative incidence: 4.6% lifetime risk (up to age 57)<br>• Incidence rate: 111 per 100,000 person-years (i.e., ~1 initial suicide attempt per 1000 people each year)<br>• 10.0% of families in Sweden had a family member attempted suicide |
| Age and sex | • Incidence rates peaked earlier in females (age 18) than males (age 20)<br>• Females had higher incidence rates than males before age 25<br>• After age 25, incidence rates were comparable but slightly higher in males |
| Major risk factors | • Lower risk: higher personal and parental social economic status (OR 0.16-0.96), being married (OR 0.63)<br>• Higher risk: having psychiatric disorders (OR 6-13), general medical diseases (OR ~2-3), and adverse life events (OR ~1.5-4), being divorce/widow (OR ~2)<br>• Major risk factors: MDD (OR 12.9), being assaulted/victimized (OR 4.4), sleep disorder (OR 3.2) |
| Outcomes | • 8.8% died at initial suicide attempt<br>• 30% had multiple suicide attempts during follow-up<br>• Relative risks of subsequent death by suicide 16.4 times<br>• Absolute risk of subsequent death by suicide within 1, 5, 10 years after the initial attempt was 0.7% vs. 0.008%, 2.1% vs. 0.06%, 3.2% vs. 0.1% for individuals with and without suicide attempt in the matched cohort<br>• Hazards for subsequent attempts and death were markedly greater in the weeks and months following the initial attempt<br>• Most people who make a suicide attempt are alive 10 years later |
| Genetic epidemiology | • Suicide attempts aggregated in families. Familial aggregation was partly due to inheritance (pedigree heritability 41.9%). The main component of environmental effects was nonshared within families (54.5%).<br>• Moderate-high genetic correlations with major psychiatric disorders (range 0.5-0.9) |
| Healthcare utilization | • In the month prior to an initial suicide attempt: 60.4% had a health care contact: pharmacy (45.6%), outpatient specialist care (37.8%), primary care (11.6%), or hospital (10.6%).<br>• In the month after an initial suicide attempt, there were multiple indications of increased contact with health care; however, 23% of those who attempted suicide had no health care contacts in the month after the event |

In our cohort with up to 47-year follow-up, 3.1% of the population were found to have a suicide attempt (lifetime prevalence of 4.6% if everyone survived to age 57). Our estimates were within the range of lifetime prevalence reported in a global survey (0.5-5%), and in line with a previous Swedish estimate.^2,3^ This translates to one initial suicide attempt for every 1,000 individuals each year. In addition to the individual effects, there are likely profound familial impact as one in ten families in Sweden had at least one individual who attempted suicide. These figures underscore the pressing need to address this significant issue faced by individuals and families as well as the healthcare system and society as a whole.

Our findings demonstrated that a suicide attempt is a marker for substantially increased mortality. Therefore, a robust and systematic focus on initial suicide attempts could be profoundly important to the societal imperative of suicide prevention. An initial suicide attempt carried multiple important subsequent health risks with 30% of individuals making additional attempts during the follow-up, and the risks for a subsequent attempt and dying by suicide increasing with the number of previous attempts. Risk of mortality was substantially elevated due to suicide and all-cause deaths. The first three months following an initial attempt was associated with particularly high mortality rate: suicide mortality of 1222 (vs. 10 among those without an attempt) and all-cause mortality of 1855 (vs. 45 among those without attempt) per 100,000 person-years. For comparison, the cumulative, long-term mortality after an initial suicide attempt was 82.4%, on par with that of 82% in cancer patients.^18^ The exceptionally high hazard ratios for mortality (17.8 for females and 15.5 for males) associated with suicide attempts underscore the critical clinical importance of an initial suicide attempt. We would argue that initial suicide attempts are the greatest single risk of mortality seen in clinical psychiatry.^19^ Although there are proven clinical interventions specifically targeted at those who survive a significant suicide attempt^20,21^, there is an implementation gap as these and other interventions are far from universally available, and certainly not commensurate with the risks. Indeed, despite of the clinical guidelines emphasizing follow-up care^22,23^, in our data, 23% of those who survived a suicide attempt did not have any record of healthcare contact within the month after the event. In our view, this critical time window for intervention represents an opportunity to save lives.

Identifying individuals at risk prior to an initial suicide attempt is also critical for suicide prevention. We identified key risk factors contributing to the first suicide attempt, including psychiatric disorders, insomnia and other general medical conditions, adverse life events, and family history. Our findings also highlight the protective effect of higher socioeconomic status, suggesting that addressing socioeconomic disparities via equitable access to mental health resources and support services may mitigate the profound societal impact. Emerging research have demonstrated the potential for developing clinically useful prediction algorithms to identify individuals at-risk for subsequent attempt and death, especially among individuals with psychiatric disorders.^24^ Further research is needed to focus on individuals who were treated at any healthcare unit for initial suicide attempts, as our finding suggest this is equally important target population for suicide prevention.

Similar to a previous study focused on suicide death^25^, we found that most individuals had contact with healthcare (60%) in the month before the first suicide attempt. Implementation of universal screening, targeted screening and clinical protocols can be beneficial in identifying at-risk individuals and providing timely interventions. We also note that nearly half (46% in Stockholm county and 47% across Sweden) visited pharmacy within the month before suicide attempt. Pharmacies were often neglected in studies investigating healthcare contacts before suicide.^26^ However, our data, together with observations that a large proportion of community pharmacists reported encounters with patients displaying suicide warning signs^27,28^, suggest that pharmacies can also play an important role in suicide prevention.

To our knowledge, this is one of the few studies that provide a comprehensive epidemiological overview of suicide attempts in the whole population. Leveraging the high-quality longitudinal registers, we capitalized on the unique opportunity to investigate a wide range of risk factors, outcomes, and genetic contributions to gain insight into suicide attempts and the healthcare system. Our work should be considered in light of several limitations. First, we leveraged register data to capture clinically-recorded suicide attempts, which would miss individuals who attempted suicide but did not present to healthcare. Consequently, our findings may have underestimated the true incidence of suicide attempts. Second, we only had primary care data from Stockholm County (∼20% of Sweden) as primary care registers in Sweden are managed regionally and not nationally. Third, the generalizability of our findings may be constrained by the selection of individuals born in Sweden and relatively young; this was intentional in order to ensure a complete follow-up and accurate identification of relatives, minimizing potential bias in estimating incidence and genetic contributions. In future work, we will investigate minoritized segments of the Swedish population particularly given the psychiatric risks carried by immigrants.^29^ Finally, in prior work^30^, we noted the empirical comparability of Sweden to advantaged countries in Scandinavia, western Europe, a few Pacific nations (Australia, Japan, South Korea), North America (Canada, United States), and Israel with respect to health care and psychiatric morbidity and mortality.

In conclusion, this study provides crucial insights into suicidal behavior. Suicide attempts are a major marker of poor health and risks for subsequent morbidity and mortality; indeed, they may carry the greatest mortal risk seen in clinical psychiatry. Our data suggest that suicide attempt presenting to any healthcare unit is relevant to trigger intervention. These results underscore the need for systematic prevention efforts for individuals who have recently attempted to die by suicide.

## Supporting information

Manuscript supplement

## Data Availability

The raw data are protected and are not available for sharing due to data privacy laws. For the Swedish register data, researchers can apply for data access at Statistics Sweden (SCB, https://www.scb.se/en/) and The National Board of Health and Welfare (Socialstyrelsen, https://www.socialstyrelsen.se/)

