## Supplementary material for "Suicide attempts: genetic and environmental risk factors, impact, and healthcare utilization—an analysis of nationwide data": Manuscript supplement

### **STUDY PROTOCOL**

**Study Name:** Suicide attempts: genetic and environmental risk factors, impact, and healthcare utilization—an analysis of nationwide data.

**Principal Investigator:**

Yi Lu, Ph.D.

Department of Medical Epidemiology and Biostatistics, Karolinska Institutet, Stockholm, Sweden

**Co-Investigators:**

Fang Fang, Ph.D.

Institute of Environmental Medicine, Karolinska Institutet, Stockholm, Sweden

Patrick F. Sullivan, M.D. FRANZCP

Departments of Genetics and Psychiatry, University of North Carolina at Chapel Hill, Chapel Hill, USA

**Background**

Suicide is a major public health challenge, with attempted suicide being a key predictor of subsequent suicide death. To date, few studies use suicide attempt as a main outcome of interest especially at a population level. The purpose of this study is to better understand individuals who have made suicide attempts and so provide novel insights to guide future suicide prevention initiatives.

**Aim**

To use national registry data to produce a comprehensive epidemiological review of suicide attempts, including incidence rates, risk factors, outcomes, genetic influences, and healthcare utilization.

**Methods**

***Data***

We will use data from multiple Swedish national registries including:

National Patient Register (NPR) to identify suicide attempts, psychiatric disorders, and general medical diseases, using the International Classification of Diseases (ICD) codes.

The Causes of Death Register to identify deaths by suicide and other causes

Total Population Register and Longitudinal Integrated Database for Health Insurance and Labor Market Studies (LISA) to identify information on sociodemographic factors and risk factors

National Prescribed Drug Register to identify dispensed medications

Multi-Generation Register and Medical Birth Register to identify family relationship

Primary care data in the Stockholm region to describe healthcare seeking for a subset of the population

***Study Design***

To study incidence and genetic epidemiology of suicide attempt, we will apply a cohort study design based on the Swedish population. Individuals will be followed from age 10 through to the either death, first emigration or end of study follow up on 31^st^ December 2019. Death by suicide is uncommon under the age of 10.98% of self-harm events in individuals under the age of 10 in our dataset were classified as ‘unintentional self-harm’. Follow up is stopped at the end of 2019 to avoid pandemic precautions impacting findings.

To study risk factors associated suicide attempts (odd ratios) we will use nested case-control study design (matching ratio 1 exposed:5 unexposed).

To study outcomes (cumulative incidence, hazard ratios) we will use a matched cohort (matching ratio 1 case:5 controls).

***Study Population***

| **Analyses** | **Inclusion criteria** | **Exclusion criteria** |
| --- | --- | --- |
| All Analyses | Born in Sweden  Date of birth between 1963-1998 | Death before age 10  First emigration record before age 10 |
| Genetic Epidemiology  Analyses | Above criteria AND  Relatives who were born outside of the cohort | Above criteria AND  Individuals who were adopted or missed information on one or both biological parents  Pairs where relative did not reach age 10 by end of follow up |

***Definition of Suicide Attempt***

The primary target population of this study are individuals who have made suicide attempts. We will define suicide attempts as an incident of self-harm with suicidal or undetermined intent that led to an individual seeking care in an emergency department, inpatient, or outpatient setting. We will identify suicide attempt ‘events’ using established methodologies involving use of ICD codes as well as indirect markers to identify events reliably within registry data.

ICD codes to define suicidal behavior from previous studies includes: Self-harm of determined intent (ICD 10 X60-X84, ICD 8/9 E95* & uncertain intent (ICD 10 Y10-Y34, ICD 8/9 E98*). We will consult clinicians about how those codes are used.

***Research questions and analysis plan***

Each outcome of interest will be analyzed using appropriate statistical methodology.

| **Questions** | **Analysis** |
| --- | --- |
| **Healthcare utilization** |  |
| Where do individuals present at first attempt of suicide? | Take first event of every ID, count % presenting to each location (Primary care, Outpatient, Inpatient) |
| What treatment is received after first attempt? | % Receiving Inpatient/Outpatient care, primary care |
| What medication is started within 1 month after the first attempt? | Extract all medication within 1 months, identify medications that patients dispensed within the month prior to event |
| What happened to the individual 1 month before the first attempt? | Number of healthcare visits (I/O/Pharmacy) |
|  | Number of visits to GP/clinical psychology |
|  | Medication use (ATC code to group by class) |
| **Epidemiology** |  |
| Population measures for burden of diseases | Cumulative incidence (incidence proportion) |
|  | Incidence rate |
|  | Cumulative incidence by sex |
|  | Incidence by calendar year of birth (stratified by sex) |
|  | Incidence by calendar year of occurrence |
|  | Incidence by age of occurrence (stratified by sex) |
|  | incidence by county of birth |
|  | Incidence by country of birth |
| What are the demographic characteristics of the suicide attempt group? | Describe the above characteristics for individuals with and without suicide attempts |
| What are the methods used in suicide attempts? | % of methods among total events (main diagnosis for method), using the first 3 characters of ICD codes provided |
|  | Split by sex |
| **Risk factor** | *Using nested case-control design. Controls are matched with cases by birth year and sex. Rationale: More efficient when doing analyses with time-varying variables* |
| What are the risk factors of suicide attempt? (Stratify by sex) | Using conditional logistic regression to estimate OR of suicide attempt in relation to risk factors |
| Individual | Socioeconomic status (Education, income, marital status) |
|  | Comorbid conditions  Psychiatric disorders (MDD, BIP, SCZ, eating disorder, substance use disorders, psychotic disorders)  General medical disorders (CVD, diabetes, obesity) |
|  | Adverse life events (family history of suicide, parental/spouse/child death, divorce) |
| Family-related | Parental socioeconomic status (Education, income, marital status) |
|  | Parental comorbidities |
| **Outcomes** | *Using matched cohort design* |
| What are the all-cause/suicide/non-suicide mortality rate at 3 months, 1y, 5y among those who have previously attempted suicide? | Cumulative incidence at 3 time points  Hazard curve or K-M curve |
| What are the relative risk for suicide/suicide/non-suicide mortality comparing those with and without a suicide attempt? | HR of outcomes |
| Describe the number of attempts? | % of individuals with single/multiple attempts |
| Describe those who died at the initial suicide attempt | % died at the initial attempt  Characteristics of those who died at the initial attempt |
| **Genetic epidemiology** |  |
| What is the proportion of Swedish families that are impacted by suicide attempt? | % of families with suicide attempt  (using both ‘genetic extended families’ and nuclear family)  Extended family includes everyone that are connect to one another via at least 1 spouse  Nuclear family= parents and offspring (1 individual can be part of >1 families) |
| Is suicide attempt aggregated within family (extended family)? | Familial aggregation of suicide attempt  OR/HR of suicide attempt among relatives comparing proband with vs without a suicide attempt  Relatedness: Parent-offspring, full sibling-half sibling |
| Is suicide attempt coaggregated with other psychiatric disorders within family (extended family)? | Familial co-aggregation with other psychiatric disorders (MDD, SCZ, BIP, SUD)  OR/HR of suicide attempt in probands whose relatives had other psychiatric disorders compared with probands whose relatives did not have psychiatric disorders |
| What is the heritability estimate? | Estimate pedigree heritability using SEM |
| To what extent suicide attempt genetically overlaps with other diseases? | Genetic correlation with other psychiatric disorders (MDD, SCZ, BIP, SUD) using SEM |

**Funding**

The Suicide Prevention Institute at the University of North Carolina, Chapel Hill, NC, USA, the Swedish Research Council, US National Institutes of Mental Health and the European Research Council.

**Expected start date**

March 2023

**Expected end date**

September 2023

**Expected submission date**

June 2024

### **SUPPLEMENTARY METHODS**

#### **Methods S1: Cohort definition and follow-up**

S1.1. **Inclusion criteria**

- Born in Sweden
- Born within period 1963-1998
- Alive and not emigrated at age 10

We included only Sweden-born individuals to form a more homogenous population. In general, immigrants are a unique group within the population with differential risk of suicide, different environmental exposures, and different patterns of mental healthcare seeking behavior.^1^ Additionally, including only those who were born in Sweden minimizes the risk of missing parental information and left-censoring of disease trajectory prior to immigration.

The birth cohort was chosen to 1) capture the peak age of suicide incident (~20-30 years old^2^); 2) identify sufficient numbers of suicide attempt cases; and 3) include adequate follow-up time to avoid misclassification of individuals with versus without suicide attempts. Since the coverage of the patient register was 1973-2019, individuals in the selected cohort were followed up between the age 0 to 56, with a minimum follow-up of 21 years (age 0-21) for the youngest birth year, and the maximum follow-up of 46 years (age 10-56) for the oldest cohort.

***For the genetic epidemiology analyses*** that included information on pairs of proband and relative, we allowed relatives to be born outside of the defined cohort period. This afforded sufficient sample size for analyses. However, relative pairs where the relative did not reach 10 years old by 31-12-2019 were not included because we started to follow individuals for suicide attempts at the age of 10.

S1.2. **Exclusion criteria**

**For the genetic epidemiology analyses,** we excluded:

- Individuals who were adopted because we cannot identify their biological relatives, to avoid biases in genetic analyses
- Individuals who were missing one or both parental personal identification (ID) numbers to ensure complete identification of siblings or had same ID for father and mother, which reflects data error
- Relative pairs where the relative had not reached age 10 by 31 December 2019

S1.3. **Follow-up period**

Start: Age 10 birthday for suicide attempt incidents; and date of birth for other variables

End: earliest date among death, first emigration date, or 2019-12-31

The follow-up period differed for different variables and analyses. In general, we started to follow the included population at the date of birth to capture medical conditions and risk factors. For suicide attempt incident, we started the follow-up at the date of 10^th^ birthday because suicide occurring before the age of 10 is uncommon in Sweden^3^, and suicide attempts captured before this age are likely to be self-harm of undetermined intent (categorized as such in 98% of the records before age 10 in our data).

We followed everyone from the start date until the earliest date among death, first emigration, or 2019-12-31. For analyses that involved incidence of suicide attempts, the follow-up period ended on the date of initial suicide attempt. Because an individual can have multiple immigration and emigration records and can utilize the Swedish healthcare system even during the period of emigration, it was difficult to capture their exact period of residence in Sweden. To avoid an overcomplicated study design, we censored at the initial emigration, i.e., assuming that after the initial emigration, individuals were not followed-up in the Swedish register.

#### **Methods S2: Family definitions**

We defined two types of family—inclusive and nuclear—using the Multi-Generation Register. In both cases, we included only families with all members born in Sweden.

***Inclusive family***: For all genetic analyses including, familial (co)aggregation and structural equation models (SEM), we defined inclusive families where parents, and full/half siblings who are connected via at least a parent or spouse. This method resulted in large and exhaustive families that are mutually exclusive (i.e., a person can only be in a single family within one generation). The families were used as clusters and were considered in our analyses to minimize biases in precision of estimates.

***Nuclear family***: For estimating the proportion of families with member(s) who attempted suicide, we additionally defined nuclear families that included only parents and full-sibling offspring (if there is). This method resulted in smaller family size and an individual can appear in multiple families.

#### **Methods S3: Statistical methods used in epidemiology section**

S3.1. **Cumulative incidence, incidence rate and hazard, competing risks**

We used the Kaplan-Meier method (survfit() function of the R package survival^4^) to estimate cumulative incidence at each age, and life-time risk (i.e., maximum cumulative incidence when the age of the population is max). Cumulative incidence was calculated as number of people having the event of interest before a specific time point divided by number of people who did not have the event or being censored (death, emigrated, or end of follow up) before the time point. When presenting cumulative incidence for different subgroup (e.g., by sex, county), we presented the life-time risk.

We further considered competing risks in the estimate of cumulative incidence by conducting some sensitivity analyses where use used Aalen-Johansen estimator instead of Kaplan-Meier method.

We used the function survRate() of the R package biostat3^5^ to obtain point estimates of incidence rate. The confidence intervals were estimated using the function stats::poisson.test(), which uses the Poisson rate confidence interval method.

We visualized the incidence rate (i.e., hazard) with confidence interval using the Nonparametric Smoothing of the Hazard Function (bshazard() function of the R package bshazard^6^).

We conducted sensitivity analyses for the estimation of cumulative incidence where the competing risk was considered (all-cause death as a competing risk of suicide attempt, and non-suicide death as a competing risk for death by suicide). We used the Aalen-Johansen estimator to estimate cumulative incidence of initial and subsequent suicide attempt, and death by suicide, taking competing risk into account and plot the estimates by age or time since the initial attempt.

S3.2. **Conditional logistic regression in nested case-control design**

We applied conditional logistic regression (function clogit() of the R package survival^4^) to estimate the odds ratio (OR) associated with the risk factors from the nested case-control design, which is mathematically approximate to the hazard ratio from a cohort design.^7^ All models were conditioned on matching clusters, hence by designed, adjusted for sex and birth year. In models 1, we did not adjust for further covariates, and in models 2 we further adjusted for socioeconomic status including education and income.

S3.3. **Stratified Cox proportional hazards regression in matched cohort design**

In the analysis of outcomes, we applied the stratified Cox proportional hazards regression (coxph() function of the R package survival^5^) to estimate hazard ratios of four outcomes - subsequent suicide attempt, suicide mortality, non-suicide mortality and all-cause mortality – in relation to having a suicide attempt. Attained age was the underlying time scale for all models. All models were stratified by matching clusters, hence by designed, adjusted for sex and birth year. In models 1, we did not further adjust for covariates, and in models 2 we further adjusted for socioeconomic status including education and income.

For the models where having subsequent suicide attempts was the outcome, if no subsequent attempts were recorded, data were censored at the end of follow-up 31/12/2019, date of death due to all causes or date of emigration.

For the models where mortality was the outcome, if death was not recorded, data was censored at the end of follow-up 31/12/2019 or date of emigration. Because we studied different causes of death, when the cause of death was not the outcome of interest, it was considered a censoring event in the analysis.

We plotted Schoenfeld’s residuals against time to detect any major violation against the proportional hazard assumption.

#### **Methods S4: Familial (co)aggregation**

We investigated the familial aggregation of suicide attempt and its co-aggregation with four other psychiatric disorders including major depressive disorder (MDD), schizophrenia (SCZ), bipolar disorder (BIP), and substance use disorder (SUD). Analyses were conducted for five types of relatedness, mother-offspring, father-offspring, full-siblings, maternal half-siblings, and paternal half-siblings. All possible pairs in families were included.

For aggregation, we fitted regression models where suicide attempts among probands was the outcome and suicide attempts among relatives was the exposure. For coaggregation, suicide attempts among probands was the outcome and psychiatric disorders among relatives were the exposures.

Probands were identified within the selected birth cohort 1963-1998, but the relatives could be born outside the selected birth cohort period. Practically, for each type of relatedness, we constructed a dataset where each pair appeared twice if both proband and relative were born within 1963-1998, with the role of proband and relative switched between the two individuals. If the relative was born outside the 1963-1998 period, the pair only appeared once.

We conducted two sets of analyses considering suicide attempt as a binary (with/without suicide attempt) and a time-to-event variable. All models included sex and birth year of the probands and relatives as covariates.

For the **binary outcome** variables, we fitted the Generalized Estimating Equations models with logit link function using the gee() function in the R package drgee^8^ to estimate the OR of having suicide attempts comparing probands whose relatives had suicide attempts or psychiatric disorders and probands without affected relatives. To avoid underestimation of standard errors due to including multiple pairs from families, we estimated robust standard errors by including family as clusters in the models.

For **time-to-event outcomes**, we applied the Cox proportional hazards regression models using the Survival R package with attained age as time scale. We estimated the HR of having suicide attempts among probands associated with suicide attempts or psychiatric disorders among relatives. Probands were considered exposed from the date when their relatives have a recorded event of interest. If there was no registered outcome in probands, data were censored at the end of follow-up 31/12/2019, date of death due to all causes, or date of emigration. We also estimated robust standard errors by including family as clusters in the models to avoid underestimation of standard errors due to including multiple pairs from families.

#### **Methods S5: Heritability and genetic correlation estimation**

We applied Structural Equation Models (SEM) to estimate heritability (*h^2^*) of suicide attempt (univariate model) and genetic correlations (*r_g_*) between suicide attempt and other psychiatric disorders (bivariate models). We assumed the liabilities (*h^2^*) and the variance shared between 2 traits (*r_g_*) come from three sources: additive genetic (A), shared environment (C), and unique environment (E), which also includes measurement error.

We used information from full-siblings and maternal half-siblings and assumed that on average, individuals of the full-sibling pairs share 50% of their additive genetic component (A) whereas individuals in the maternal half-sibling pairs shared 25%. Individuals in the full-sibling pairs and maternal half-sibling pairs were assumed to share 100% of their common environmental contribution (C) on average. A previous study using similar data showed that in Sweden, the majority of both full-siblings and maternal half-siblings grew up in the same household.^9^

The models were fitted using Weighted Least Square applied in the LISREL models in the OpenMx software version 2.19.8. We adjusted for sex (0 as male, 1 as female) and birth year (mapped into range 0-1 to increase the model-fitting optimization efficiency).

Because we included all pairs of siblings in all families to maximize sample size, there was dependency between the pairs. We used Bootstrap resampling (1000 replicates) to estimate confidence intervals (CI). 95% CIs were estimated as the range between the 26th and the 975th of the ascending-sorted 1000 Bootstrap estimates. We used inclusive families as a unit for simple random sampling with replacement (srswr). For each Bootstrap sample, full-siblings and maternal-half sibling data were drawn separately. The number of families being drawn was equal to the number of inclusive families.

#### **Methods S6: Sankey diagram of healthcare and medication utilization**

We described healthcare and medication utilization in relation to the presentation of the initial suicide attempt. To achieve this, we used a Sankey diagram to show the proportions of individuals with a suicide attempt in different categories before and after their initial suicide attempt. The primary care data were only available for the Stockholm region until 31/12/2013, and the Prescribed Drug Register started in 1/7/2005. Therefore, for these analyses, we only included individuals whose initial suicide attempt records occurred from 1/8/2005 until 30/11/2013. This period was chosen so that we could examine medication use within one month prior to and after the initial suicide attempt presentation. The analyses involved 3646 individuals with total 18,968 records of suicide attempt.

The results presented in *Tables S13, S14* were obtained from the same population used for the Sankey diagram.

We note that our data do not include all diagnoses, but are limited to those approved in the project’s ethical approval. “Clinical contacts” referred to all primary care contacts and specialist care contacts for the conditions listed below. Primary care included all contacts to the public and private providers operating with subsidies in the Stockholm region. Clinical contacts in specialist care (i.e., inpatient and outpatient) were limited to the following conditions (respective codes in ICD-10):

F00-F99, G00-G47, G60-G73, G91, A00-B99, D80-D89, H10, J00-J46, P27, R47-R49, R05-R06, K25-K37, K50-K51, K70-K85, K90, L00-L54, L90-L95, M00-M68, N00-N08, N10-N12, N30, N20-N21, N41- N49, N97, I00-I79, I98.3, O10-O16, O20-26, O85-86, P00-P96, E00-E35, E65-E68, Q00-Q99, S00-T98, V01-Y98.

### **SUPPLEMENTARY TABLES**

#### **Table S1: Literature review**

| **Author, year** | **Title** | **Population (Sample size)** | **Phenotype** | **Results summary** |
| --- | --- | --- | --- | --- |
| **Burden of diseases** |  |  |  |  |
| Kendler, 2023^10^ | Genetic liability to suicide attempt, suicide death, and psychiatric and substance use disorders on the risk for suicide attempt and suicide death: a Swedish national study | Individuals born in Sweden during 1932–1995, to Swedish-born parents  (N=7,661,519) | Suicide attempt (ICD-10: X60-X84, Y10-Y34 and equivalent codes in ICD-8/9) registered in inpatient, outpatient, and primary care | Prevalence of suicide attempt without death 3.27% |
| National Centre for Suicide Research and Prevention, 2021^11^ | Suicide attempts in Sweden | Swedish residents | Suicide attempt (ICD-10: X60-X84, Y10-Y34 and equivalent codes in ICD-8/9) registered in inpatient care | Rate ranged 80.4-128.2 patients per 100,000 inhabitants each year during 1987-2022  Highest rates among age group 15-24 in all years (1987-2022) |
| Nock, 2018^12^ | Cross-national prevalence and risk factors for suicidal ideation, plans and attempts | 17 countries from Africa, America, Asia and the Pacific, Europe, and the Middle East (N=84,850) | Suicide attempt (self-reported using the WHO Composite International Diagnostic Interview) | Lifetime prevalence 2.7% (SE 0.1) |
| **Risk factors** |  |  |  |  |
| Wiktorsson, 2022^13^ | Clinical Characteristics in Older, Middle-Aged and Young Adults Who Present With Suicide Attempts at Psychiatric Emergency Departments: A Multisite Study | Individuals who presented with self-harm at psychiatric emergency departments and had suicidal intent (N=683; age 18-44 years: N=423, age 45-64: N=164, age 65+: N= 96) | Self-harm with suicidal intent assessed by Columbia Suicide Severity Rating Scale (C-SSRS) | Comparing characteristics of 3 age groups, older adults scored higher than the younger groups on some suicidal intent scales.  Age group 65+ was more likely to have major depression and serious physical illness, and less likely to have anxiety, alcohol and substance use disorders. |
| Chen, 2020^14^ | Predicting suicide attempt or suicide death following a visit to psychiatric specialty care: A machine learning study using Swedish national registry data | Individuals aged 18-39 with inpatient and outpatient diagnosis of psychiatric disorders from Swedish registers  (N=126,205 individuals, 541,300 visits) | Suicide attempt from patient register: Intentional self-harm (ICD-10: X60–X84) or self-harm of undetermined intent (ICD-10: Y10-Y34) recorded as unplanned inpatient or outpatient visits  Suicide death from Cause of Death register: (ICD-10: X60–X84) or self-harm of undetermined intent (ICD-10: Y10-Y34) | Predict suicide attempts and deaths using information at visits to psychiatric care.  Proportions of visits followed by a suicide attempt/death at 30 days=1.7%, 90 days=3.5%.  Important predictors: Important predictors: Intentional self-harm during the past 1 year; unplanned visit to psychiatric specialty care service during the past 1 to 3 months; diagnosis of borderline personality disorder during the past 3 months; diagnosis of depressive disorder during the past month; recent dispensation of antidepressants within 6 months; anxiolytics within 12 months; benzodiazepines within 12 months; and antipsychotics within 3 years; prior intentional self-harm by poisoning or sharp object; family history of suicide attempt, substance use disorder, and borderline personality disorder. |
| Yates, 2019^15^ | Association of psychotic experiences with subsequent risk of suicidal ideation, suicide attempts, and suicide deaths: A systematic review and meta-analysis of longitudinal population studies | Systematic review included 8 studies from multiple countries (N=66,967) | Self-reported and clinician diagnosis of suicide attempt | Psychotic experiences associated with future suicide attempt: OR 3.15 (95% CI 2.23-4.45) |
| Givion, 2018^16^ | Serious suicide attempts: Systematic review of psychological risk factors | Systematic review included 39 studies investigating psychological factors associated with serious suicide attempts | Serious suicide attempts measured by various indicators including lethality, level of serious intent to die, medical consequences | Major risk factors: mental pains (depression, anxiety, hopelessness, distress), communication difficulties, decision-making impulsivity, aggression, adverse life events, childhood and family adversities |
| Nock, 2018^12^ | Cross-national prevalence and risk factors for suicidal ideation, plans, and attempts | 17 countries from Africa, America, Asia and the Pacific, Europe, and the Middle East (N=84,850) | Self-reported suicide attempts | Risk factors: being female, younger age, low education, unmarried, prior mental disorders |
| Brezo, 2006^17^ | Personality traits as correlates of suicidal ideation, suicide attempts, and suicide completions: a systematic review | Systematic review included 90 studies investigating the association between personality traits and suicide attempts, suicidal ideation, and suicide completions | Suicide attempts with diverse definitions | Risk factors: Neuroticism, impulsivity, hopelessness, hostility, anger, self-criticism, perceptual aberration.  Protective factor: Extroversion |
| **Outcome** |  |  |  |  |
| Demesmaeker, 2022^18^ | Suicide mortality after a nonfatal suicide attempt: A systematic review and meta-analysis | Systematic review included 41 cohort studies and randomized controlled trials from Europe and the US  (N range 58-39,685 individuals) | Non-fatal suicide attempts from clinical settings | Suicide rate after a nonfatal suicide attempt at:  1-year 2.8% (95% CI 2.2–3.5)  5-year 5.6% (95% CI 3.9–7.9)  10-year 7.4% (95% CI 5.2–10.4) |
| Soullane, 2022^19^ | Relationship between suicide attempt and medical morbidity in adolescent girls | Girls <20 years old during 1989-2019 in Quebec, Canada  (N=169,806; 8086 with suicide attempts) | Suicide attempts (ICD-9: E950-E959; ICD-10: X60-X84, Y87.0) in patient records of a hospital | Individuals with suicide attempts had higher risk of mortality (HR 3.11; 95% CI 1.69–5.70), hospitalization due to infection (HR 1.55; 95% CI 1.44–1.68), allergy (HR 1.72; 95% CI 1.45–2.05), and cardiovascular (HR 1.31; 95% CI 1.12–1.52).  Individuals with repeated suicide attempts had greater risks compared to individuals with only one recorded attempt.  Risk of hospitalization was more pronounced the first few years after an attempt. |
| Carr, 2017^20^ | Premature death among primary care patients with a history of self-harm | Individuals aged 15 to 64 years with a recorded episode of self-harm  (N=630,275 individuals; 30,017 with suicide attempts) | Self-harm with and without suicidal intent ascertained by list of Read codes to identify all cases of self-harm across the spectrum from milder forms of non-suicidal behavior to near-fatal suicide attempts | Compared to individuals without self-harm, those with self-harms had higher risk of death by all causes and by suicide. The risks were highest after 1 year then attenuated.  Hazard ratio of all-cause death:  ≤1 year: 3.59 (3.08–4.19)  >1 year: 1.70 (1.54–1.88)  Hazard ratio of suicide death:  ≤1 year: 54.43 (34.32–86.32)  >1 year: 7.62 (5.67–10.25 |
| Goldman-Mellor,  2014^21^ | Suicide attempt in young people: A signal for long-term health care and social needs | Individuals born between 4/1972 and 3/1973 in Dunedin, New Zealand  (N=1037; 91 with suicide attempts) | Self-reported self-injury with suicidal intent at age 24 or younger via interview | Compared to individuals without suicide attempt, those with a suicide attempt had:  **Poorer mental health:**  2 times risk of persistent episodes of major depression and persistent substance dependence; 3 times risk of subsequent suicide attempts; 3 times risk of subsequent non-suicidal self-injury; Required more mental health–related services (help-seeking, medication, hospitalization) **Poor physical health:**  More daily functional limitations, non-suicidal injuries  Higher levels of inflammatory markers; 2 times risk of metabolic syndrome; 4 years older heart age than actual age  **More harm towards others:**  2 times risk of being abusive in intimate relationships and convicted for a violent crime **Need more support:**  Longer unemployment; Longer dependence on welfare benefits  **Worse quality of life:**  Being physically victimized by romantic partners; Suffering from loneliness; Less satisfied with lives |
| Runeson, 2010^22^ | Method of attempted suicide as predictor of subsequent successful suicide: national long term cohort study | Individuals admitted to hospital in after attempted suicide during 1973-1982  (N=48,649) | Attempted suicide (ICD-8: E950-E959, E980-E989) predicting suicide death | 12% died by suicide after an attempt.  Individuals who attempted suicide by hanging, strangulation, or suffocation had the highest risk of death.  Among those who died, 87% happened within a year after the first attempt.  Most of those who died by suicide used the same method as they did at the first attempt. |
| Tidemalm, 2008^23^ | Risk of suicide after suicide attempt according to coexisting psychiatric disorder: Swedish cohort study with long term follow-up | Individuals admitted to hospital for attempted suicide during 1973-1982  (N=39,685) | Attempted suicide (ICD-8: E950-E959, E980-E989) predicting suicide death | Psychiatric disorder coexistent with a suicide attempt influences overall risk and temporality for suicide death.  Highest short-term risk associated with depression and bipolar disorder.  Strongest predictors for overall risk were schizophrenia, depression and bipolar disorder. |
| **Genetics** |  |  |  |  |
| Kendler, 2023^10^ | Genetic liability to suicide attempt, suicide death, and psychiatric and substance use disorders on the risk for suicide attempt and suicide death: a Swedish national study | Individuals born in Sweden during 1932–1995, to Swedish-born parents  (N=7,661,519; 3.27% suicide attempts; 0.57% suicide death) | Suicide attempt (ICD-10: X60-X84, Y10-Y34 and equivalent codes in ICD-8/9) registered in inpatient, outpatient, and primary care | Genetic susceptibilities to suicide attempt and suicide death are related but not identical;  Family genetic risk score (FGRS) for suicide attempt and alcohol use disorder were higher in predicting suicide attempts, while the FGRS for suicide death, bipolar disorder, and schizophrenia were higher in predicting suicide deaths. |
| Edwards, 2021^24^ | On the genetic and environmental relationship between suicide attempt and death by suicide | Twins, full siblings, and half siblings born in Sweden during 1960-1990 (N=1,314,990; 21,664 females and 21,854 males attempted suicide; 1,048 females and 3,109 males died by suicide) | Suicide attempt (ICD-10: X60-X84, Y10-Y34 and equivalent codes in ICD-8/9) registered in inpatient and outpatient | Heritability of suicide attempt: Females 0.52 (95% CI=0.44-0.56)  Males 0.41 (95% CI=0.38-0.49) Heritability of suicide death: Females 0.45 (95% CI=0.39-0.59)  Males 0.44 (95% CI=0.43-0.44)  Genetic correlation between suicide attempt and suicide death: Female 0.67 (95% CI=0.55-0.67)  Male 0.74 (95% CI=0.63-0.87) |
| Kendler, 2020^25^ | The sources of parent-child transmission of risk for suicide attempt and deaths by suicide in Swedish National Samples | Individuals born in Sweden during 1960-1990  (N offspring: 2,175,259 in intact families; 152,436 not lived with father; 73,785 lived with stepfather; 15,624 in adoptive families.  Suicide attempt among offspring: 2.8% in intact families; 6.3% in those not lived with father, 5.5% in those lived with stepfather, 6.7% among those lived in adoptive families) | Suicide attempt (ICD-8/9: E95 and E98; ICD-10: X60–X89, Y10–Y34, Y87, Y90, Y91) registered in inpatient and outpatient | Suicide attempt transmitted through generation Genes only (correlation 0.13, 95% CI 0.11-0.15) Genes plus rearing (correlation 0.23, 95% CI 0.23-0.24) Rearing only (correlation 0.14 (95% CI 0.11-0.16); Suicide attempt was more strongly transmitted to male offspring compared with female offspring; Parental psychiatric disorders accounted for 40% of the genetic transmission but had no impact on rearing effects |
| Cho, 2006^26^ | Genetic contribution to suicidal behaviors and associated risk factors among adolescents in the U.S. | Adolescents in grades 7–12 in the United States from the National Longitudinal Study of Adolescent Health (Add Health)  (N=1448 individuals, 724 twin pairs) | 1 self-reported item in questionnaire | Heritability 24% (calculated from concordance rate using Holzinger’s formula: 3 of 13 MZ pairs and 2 of 21 DZ pairs were concordant; 37.5% versus 17.4%; H_C_ = 24%) |
| Baldessarini, 2004^27^ | Genetics of suicide: an overview | Systematic review included 22 studies (N~25,000 individuals with suicidal behavior and family members) | Compare risks of suicides or of serious attempts among close relatives of index individuals with suicidal behavior, with such risks among relatives of non-suicidal or healthy controls. | Familial risk of suicidal behavior: Combined risk ratio 2.86 (95% CI 2.32–3.53) |
| Fu, 2002^28^ | A twin study of genetic and environmental influences on suicidality in men | Twin pairs from the Vietnam Era Twin Registry who were assessed in 1987 and 1992  (N=3372 twin pairs) | Suicide ideation and suicide attempt from self-reported | Twins whose monozygotic (MZ) co-twin had a record of suicidal ideation showed a higher tendency to report suicidal ideation (OR 2.96) or suicide attempt (OR 5.34).  Twins whose MZ co-twin had a history of suicide attempts had an increased likelihood of reporting suicidal ideation (OR 4.30) or suicide attempt (OR 12.06), compared to twins whose co-twin hadn't reported any suicidal behaviors.  Heritability of suicide attempts: 17.4% (95% CI 13.8–43.7)  Heritability of suicide ideation: 36.0% (95% CI 15.5–41.3) |
| Glowinski, 2001^29^ | Suicide attempts in an adolescent female twin sample | Missouri female adolescent twins, mean age 15.5 years  (N=3,416 individuals; 4.2% with suicide attempts) | Self-reported suicide attempt | Odds ratio for twin/co-twin suicide attempt: 11.6 (95% CI 4.7–28.6) for MZ twins; 4.2 (95% CI 1.2–15.3) for DZ twins  Heritability: 48% (95% CI 0–73.2) |
| Statham, 1998^30^ | Suicidal behaviour: an epidemiological and genetic study | Twins from an Australian twin panel  (N=5995 individuals; 3.0% female and 2% male with suicide attempts) | Self-reported suicide attempt | Heritability: 43% for any suicidal thoughts; 44% for persistent thoughts, plans or minor attempt; and 55% for serious suicide attempt (No CI provided) |
| **Healthcare utilization** |  |  |  |  |
| Probert-Lindström, 2023^31^ | Utilization of psychiatric services prior to suicide- a retrospective comparison of users with and without previous suicide attempts | Individuals who died by suicide in Sweden  (N=484) | Death by suicide from Cause of Death register (ICD-10: X60–X84) | 51% had previous suicide attempts  Compared to those without previous suicide attempt, those with previous suicide attempts were more likely to: have psychiatric diagnosis; use psychotropic medications; be absent from appointments during the last three months before death; have higher suicide risk in the psychiatric assessment |

#### **Table S2: Register data**

| **Register Name** | **Coverage year** | **Recorded information** |
| --- | --- | --- |
| Total Population Register (TPR)^32^ | Started in 1968;  Covers ~100% population | Dates of birth, sex, marital status |
| Swedish Medical Birth Register^33^ | Started in 1973;  Covers ~98% of births in Sweden | Maternal data (health and lifestyle), birth and neonatal care data;  Used to identify individuals who were born in Sweden |
| National Patient Register (NPR)^34^ | The NPR includes information on hospital-based inpatient healthcare utilization since 1964 and has covered inpatient care of psychiatric disorders nationwide since 1973. From 2001 onward, the NPR also includes information on specialist outpatient care. Covers ~100% for both psychiatric and somatic diseases in 1978 in inpatient care, ~80% for outpatient care (missingness mainly due to not including data from private caregivers) | Inpatient discharges and outpatient visits (Not including primary care records);  Diagnoses pertaining to visit (ICD codes), date of discharge and location of discharge, length of admission, procedures completed. Each entry in the NPR lists a primary diagnosis and up to 30 secondary diagnoses |
| Multi-Generation Register^35^ | Started in 1947;  Covers 97% on mothers, 95% on fathers of index persons | ID of parents of index persons who were born from 1932 onwards and were alive on 1/1/1961 |
| Cause of Death Register^36^ | Started in 1952;  Covers ~100% death in Sweden | The register records the date of death and the underlying and contributing causes of each death, coded according to the ICD system. In Sweden, an apparent or suspected unnatural death typically necessitates a forensic autopsy, which includes toxicological analysis |
| Longitudinal Integrated Database for Health Insurance and Labor Market Studies (LISA)^37^ | Covers years 1990-2019 | Education, employment, sick leave, disability pension |
| National Prescribed Drug Register^38^ | 1/7/2005-31/12/2015 | Prescribed drugs dispensed at pharmacies in Sweden (ATC codes) |
| Stockholm Primary Care register^39^ | 2003-2013 (period that overlap with the available National Patient Register data) | Primary care visits at public and private providers operating with subsidies in Stockholm;  Date and diagnoses pertaining to visit (ICD codes) |

#### **Table S3: ICD codes of suicide attempts, psychiatric disorders and general medical diseases**

| **Phenotype** | **ICD 10 (1997-2019)** | **ICD 9 (1987-1996)^a^** | **ICD 8 (1968-1986)** |
| --- | --- | --- | --- |
| **Any self-harm including suicide attempt** | 1. Intentional self-harm/poisoning (ICD-10: X60-X84); 2. Suicide and self-inflicted poisoning/injury (ICD-8/9: E950-E959); 3. Self-harm with undetermined intent (ICD-10: Y10-Y34, ICD-8/9: E980-E989); 4. Sequelae (ICD-10: Y87.0, Y87.2) 5. Death with any X, Y, E codes from Cause of death register | | |
| **Suicide attempt** | 1. X60-X84 2. Any of the other codes (Y10-Y34, Y87.0, Y87.2, E950-E959, E980-E989)   **AND** meeting one of the two additional conditions:   a. Used a notably lethal method of self-harm (firearm, jumping from heights, motor vehicle crash, suffocation, or poisoning by cooking gas) b. Led to inpatient care 6. Death with any X, Y, E codes from Cause of death register | | |
| **Death by Suicide using Cause of death register** | X60-X84, Y10-Y34, Y87.0, Y87.2 | E950-E959 | E950-E959 |
| **Major depressive disorder (MDD)** | F32, F33 | 296.3 (296B), 311 | 300.4 |
| **Schizophrenia, Schizoaffective disorder (SCZ)** | F20.0, F20.1, F20.2, F20.3, F20.4, F20.5, F20.6, F20.8, F20.9, F25.0, F25.1, F25.2, F25.8, F25.9 | 295.0 (295A), 295.1 (295B), 295.2 (295C), 295.3 (295D), 295.4 (295E), 295.6 (295G), 295.8 (295X), 295.9 (295W), 295.7 (295H) | 295.0, 295.1, 295.2, 295.3, 295.4, 295.6, 295.7, 295.8, 295.9 |
| **Bipolar, manic disorder** | F30.1, F30.2, F30.8, F30.9, F31 | 296.1 (296D), 296.4 (296E), 296.5 (296F), 296.6 (296G), 296.7 (296H), 296.89 (296W), 296.99 (296X) | 296.1, 296.3, 296.8 |
| **Eating disorder** | F50.0, F50.1, F50.2, F50.3, F50.4, F50.5, F50.8, F50.9 | 307.1 (307B), 307.5 (307F) | N/A |
| **Mental and behavioral disorders due to psychoactive substance use (excluding tobacco) (SUD)** | F10, F11, F12, F13, F14, F15, F16, F18, F19 | 291, 292, 303, 304, 305 (due to truncation of the code, we couldn’t remove 305.1 (305B) which is tobacco abuse | 291, 303, 304 |
| **Psychotic disorders** | F20, F21, F22, F23, F24, F25.0, F25.1, F25.2, F25.8, F25.9, F28, F29, F30.2, F31.2, F31.5, F31.64, F32.3 F33.3 | 295.0 (295A), 295.1 (295B), 295.2 (295C), 295.3 (295D), 295.4 (295E), 295.6 (295G), 295.7 (295H), 295.8 (295X), 295.9 (295W), 297.0 (297A), 297.1 (297B), 297.2 (297C), 297.3 (297D), 297.8 (297X), 297.9 (297W), 298.0 (298A), 298.1 (298B), 298.3 (298D), 298.4 (298E), 298.8 (298W), 298.9 (298X) | 295.0, 295.1, 295.2, 295.3, 295.4, 295.6, 295.7, 295.8, 295.9, 297.0, 297.1, 297.9, 298.0, 298.3, 298.9, 296.1, 296.2, 296.3, 296.8, 296.9 |
| **Type 1 and 2 diabetes** | E10-E11 | 250 | 250 |
| **Cardiovascular disease** | I00-I70, I73.0, I74-I75 | 390-438, 440, 444, 445 | 390-438, 440, 444, 445, 450-453, 458 |
| **Obesity** | E65, E66 | 278.0 (278A), 278.1 (278B) | 277 |
| **Sleep disorder** | G47.0-G47.9 | 307.4 (307E), 780.5 (780F) | 306.40 |
| **Chronic pain** | F52.5, F52.6, G43, G44, G93.3, G50.0, G50.1, G54.6, H57.1, H92.0, K07.6, K21, K58,  M00-M19, M20-M25, M30-M36, M40-M54, M60-M99, M79.7, N20-N23, N30.1, N41.1, N94, N48.3, R07, R10, R12, R26, R30, R51, R52 | 346.0 (346A), 346.1 (346B), 346.8 (346X), 346.9 (346W) | 346.09 |
| **Malformation** | Q00-Q99 | 74, 75 | 74, 75, 31 |
| **Injury** | S00-S99, T00-T98 | N/A | N/A |
| **Traumatic brain injury** | S02.0, S02.1, S02.7, S02.8, S02.9, S04.0, S06, S07.1 | 800-804, 850-854 | 800-804, 850-854 |
| **Accident** | V01-V99, W00-W99, X00-X59 | E800-E949 | E807-E949 |
| **Assault/ victimization** | X85-X99, Y0 | E960-E969 | E960-E969 |

*If the last characters of the code were not specified, all codes starting with the specified characters were included*

*^a^Swedish version of the code, if different from the international codes, is specified in brackets*

#### **Table S4: Variable definitions**

| **Variable** | **Definition** | **Cut-off/categories** |
| --- | --- | --- |
| **Subgroups to describe burden of disease** |  |  |
| Sex | Registered sex retrieved from the Total Population Register^32^ | Binary (Male/Female) |
| Age | Age at the initial suicide attempt (year) | Continuous variable  To describe the incidence rate of suicide attempt by age at attempt and calendar year, we split the follow-up at every 5 years of age between 10 and 60, and split the calendar years at 1975, 2015 and every 5 years in between. |
| **Methods** |  |  |
| Methods of suicide attempt | Methods of attempting suicide based on 3 first characters of ICD-10 codes, and 4 first characters of ICD-8/9 codes | Drug/poison: X60, X61, X62, X63, X64, X65, X66, X68, X69, Y10, Y11, Y12, Y13, Y14, Y15, Y16, Y18, Y19, E950, E980  Firearm: X72, X73, X74, X75, Y22, Y23, Y24, Y25, E955, E985  Gas: X67, Y17, E951, E952, E981, E982  Heat: X76, X77, Y26, Y27  Jump/crash: X80, X81, X82, Y30, Y31, Y32, E987, E957  Object: X78, X79, Y28, Y29, E956, E986  Suffocation: X70, X71, Y20, Y21, E953, E954, E983, E984  Others (sequalae, unspecified means): X83, X84, Y870, Y33, Y34, Y872, E958, E959, E988, E989 |
| Lethal methods | Based on ICD codes for firearm (X72, X73, X74, X75, Y22, Y23, Y24, Y25, E955, E985), jumping from heights or motor vehicle crash (X80, X81, X82, Y30, Y31, Y32, E957, E987), suffocation (X70, X71, Y20, Y21, E953, E954, E983, E984), and poisoning by domestic gas (E951, E981) | Binary variable (yes/no) |
| **Other Risk factors^a^** |  |  |
| ***Socio-economic status*** |  |  |
| Education | Highest education level that was recorded in the year before the date of initial suicide attempt. If the value was missing in that year, we imputed using the most recent non-missing value.  Variables SUN2000NIVA_OLD (1990-2018) and SUN2020NIVA_OLD (2019-2020) from LISA (reported on a yearly basis). | Categorized the number of education years into 4 groups: 1) Primary school (<9 years); 2) Secondary school (9-12 years); 3) Post-secondary (>12 years); 4) Unknown (included class ‘unknown’ and missing value in the data) |
| Income | Disposable income per consumption unit within the family which is calculated by using family disposable income divided by consumption unit (calculated by number of family with weights assigned for adults/children).^37^ Disposable income was recorded in the year before the date of the initial suicide attempt. If the value was missing in that year, we imputed using the most recent non-missing value. Variables DispInkKE04 (2005-2020) and DispInkKE (1998-2020) from LISA (reported yearly) | Categorized based on rank in tertile by year among the included individuals:  1) Low; 2) Medium; 3) High; 4) Unknown |
| Marital status | Marital status recorded in the year before the date of initial suicide attempt.  *Note: Marital status is updated yearly for each individual and recorded in the variable CIVIL in the Total Population Register. | 4 categories: 1) Unmarried; 2) Married; 3) Divorced; 4) Widow(er) |
| ***Comorbidities*** |  |  |
| Psychiatric disorders: Major depressive disorder, schizophrenia, bipolar disorder, substance use disorders, eating disorder, psychotic disorders | Having any diagnosis of the listed disorders in the Patient Register before the date of the initial suicide attempt.  ICD codes to define the disorders in *Table S3* | Binary variable (yes/no) |
| General medical diseases: Type 1/2 diabetes, cardiovascular disease, obesity, sleep disorder, chronic pain, malformation | Having any diagnosis of the listed diseases in the Patient Register before the date of the initial suicide attempt.  ICD codes to define the diseases in *Table S3* | Binary variable (yes/no) |
| ***Adverse life events*** |  |  |
| Experience of any injury, Traumatic brain injury, accident, Assault/ victimization | Having any diagnosis of the listed conditions in the Patient Register before the date of the initial suicide attempt.  ICD codes to define the conditions in *Table S3* | Binary variable (yes/no) |
| Divorce | Ever had marital status as “Divorced” before the year of initial suicide attempt | Binary variable (yes/no) |
| Loss of spouse | Ever had marital status as “Widow(er)” before the year of initial suicide attempt | Binary variable (yes/no) |
| Loss of parent | Loss of any biological/adoptive parent before the initial suicide attempt | Binary variable (yes/no) |
| Parental suicide death | Any suicide death of any biological/adoptive parent before the index individual’s initial suicide attempt | Binary variable (yes/no) |
| Parental suicide attempt | Any suicide attempt of any biological/adoptive parent before the index individual’s initial suicide attempt | Binary variable (yes/no) |
| Parental psychiatric disorder | Diagnosis of any psychiatric disorder (listed above) among biological/adoptive parents, before the index individual’s initial suicide attempt | Binary variable (yes/no) |
| Parental substance use disorder | Diagnosis of any substance use disorder among biological/adoptive parents, before the index individual’s initial suicide attempt | Binary variable (yes/no) |
| Parental education | Highest level of education among biological/adoptive parents before the index individual’s initial suicide attempt. If the value was missing in that year, we imputed using the most recent non-missing value. | Categorized the number of education years into 4 groups: 1) Primary school (<9 years); 2) Secondary school (9-12 years); 3) Post-secondary (>12 years), 4) Unknown (included class ‘unknown’ and missing value in the data) |
| Parental income | Highest level of income rank among biological/adoptive parents before the index individual’s initial suicide attempt. Disposable income per consumption unit within the family which is calculated by using family disposable income divided by consumption unit (calculated by number of family with weights assigned for adults/children).^37^ If the value was missing in that year, we imputed using the most recent non-missing value. Variables DispInkKE04 (2005-2020) and DispInkKE (1998-2020) from LISA (reported yearly) | Categorized based on rank in tertile by year among the included cohort:  1) Low; 2) Medium; 3) High; 4) Unknown |
| Loss of any child | Loss of any biological/adopted child before the initial suicide attempt | Binary variable (yes/no) |
| **Outcomes** |  |  |
| All-cause mortality | Death identified from the Total population register using registered date of death | Time-to-event variable |
| Suicide mortality | Death by suicide identified from the Cause of Death register using ICD codes (*Table S3*) | Time-to-event variable |
| Non-suicide mortality | Death by suicide identified from the Cause of Death register using ICD codes, excluded those with suicide death | Time-to-event variable |
| Subsequent suicide attempt | Defined using patient record for suicide attempt. All records with overlapping admission and discharge date were considered belonging to the same attempt. All records were arranged chronologically based on admission date, a record was considered marking a new attempt if its admission date was after the discharge date of the previous record. | Time-to-event variable |

*^a^Risk factors were selected based on the literature^40^*

#### **Table S5: Characteristics of individuals with and without suicide attempt/self-harm**

| **Suicide attempt** | | | | | |
| --- | --- | --- | --- | --- | --- |
| **Characteristics** | **No suicide attempt**  **N=3,582,730** | | **Suicide attempt**  **N=113,890** | | **p-value^a^** |
| **Sex** |  | |  | | <0.001 |
| Female | 1,735,126 (48.4%) | | 62,893 (55.2%) | |  |
| Male | 1,847,604 (51.6%) | | 50,997 (44.8%) | |  |
| **Age at end of follow-up** |  | |  | |  |
| Mean (SD) | 39.3 (10.5) | | 40.2 (10.3) | | <0.001 |
| Range | 21.0 - 57.0 | | 21.0 - 57.0 | |  |
| Median | 39.4 | | 40.0 | |  |
| Interquartile range | 29.8 - 48.7 | | 31.2 - 49.6 | |  |
| **Self-harm** | | | | | |
| **Characteristics** | **No self-harm**  **N=3,538,841** | **Self-harm**  **N=157,779** | | **p-value^a^** | |
| **Sex^a^** |  |  | | <0.001 | |
| Female | 17,19,130 (48.6%) | 78,889 (50.0%) | |  | |
| Male | 1,819,711 (51.4%) | 78,890 (50.0%) | |  | |
| **Age at end of follow-up** |  |  | |  | |
| Mean (SD) | 39.3 (10.6) | 39.5 (10.3) | | <0.001 | |
| Range | 21.0 - 57.0 | 21.0 - 57.0 | |  | |
| Median | 39.5 | 38.8 | |  | |
| Interquartile range | 29.9 - 48.7 | 30.5 - 48.8 | |  | |

*Proportion was calculated among the group of suicide attempt/self-harm status (i.e., by column)*

*^a^Test for statistical difference in the characteristic between 2 groups with and without suicide attempt/self-harm using chi-squared test for categorical variables and t-test for continuous variables.*

#### **Table S6: Characteristics of individuals who died/survived at the initial suicide attempt**

| **Characteristics^a^** | **Died initial attempt**  **N=10,033** | **Survived initial attempt**  **N=103,857** |
| --- | --- | --- |
| **Sex** | | |
| Female | 2309 (23.0%) | 60584 (58.3%) |
| Male | 7724 (77.0%) | 43273 (41.7%) |
| **Age at the index date** | | |
| Mean (SD) | 30 (10) | 25 (9) |
| Range | 10 - 57 | 10 - 57 |
| Median | 28 | 22 |
| Interquartile range | 22 - 38 | 18 - 30 |
| **Year of birth** |  |  |
| 1963-1969 | 3921 (39.1%) | 23279 (22.4%) |
| 1970-1979 | 3027 (30.2%) | 26783 (25.8%) |
| 1980-1989 | 2039 (20.3%) | 30707 (29.6%) |
| 1990-1998 | 1046 (10.4%) | 23088 (22.2%) |
| **Educational attainment before the year of the index date** | | |
| Primary school (<9y) | 2491 (24.8%) | 34631 (33.3%) |
| Secondary school (9-12y) | 4638 (46.2%) | 35893 (34.6%) |
| Post-secondary (>12y) | 1545 (15.4%) | 8405 (8.1%) |
| Unknown | 1359 (13.5%) | 24928 (24.0%) |
| **Disposable income before the year of the index date (rank in tertile by year)** | | |
| Low | 3159 (31.5%) | 35079 (33.8%) |
| Medium | 2051 (20.4%) | 20069 (19.3%) |
| High | 1989 (19.8%) | 14459 (13.9%) |
| Unknown | 2834 (28.2%) | 34250 (33.0%) |
| **Marital status in the year before index date** |  |  |
| Unmarried | 8408 (83.8%) | 91000 (87.6%) |
| Married | 993 (9.9%) | 7808 (7.5%) |
| Divorced | 619 (6.2%) | 4951 (4.8%) |
| Widow(er) | 13 (0.1%) | 98 (0.1%) |
| Unknown | 0 (0.0%) | 0 (0.0%) |
| **Highest education level of parents** |  |  |
| Primary school (<9y) | 1609 (16.0%) | 13102 (12.6%) |
| Secondary school (9-12y) | 4412 (44.0%) | 50043 (48.2%) |
| Post-secondary (>12y) | 2797 (27.9%) | 28249 (27.2%) |
| Unknown | 1215 (12.1%) | 12463 (12.0%) |
| **Highest income level of parents (tertile rank by year)** |  |  |
| Low | 1967 (19.6%) | 22542 (21.7%) |
| Medium | 2489 (24.8%) | 28258 (27.2%) |
| High | 2704 (27.0%) | 25989 (25.0%) |
| Unknown | 2873 (28.6%) | 27068 (26.1%) |
| **Any of the following mental disorders (any diagnosis by a specialist before the index date)** | | |
| No | 6082 (60.6%) | 65402 (63.0%) |
| Yes | 3951 (39.4%) | 38455 (37.0%) |
| **Major depressive disorder (any diagnosis by a specialist before the index date)** | | |
| No | 8154 (81.3%) | 82731 (79.7%) |
| Yes | 1879 (18.7%) | 21126 (20.3%) |
| **Schizophrenia (any diagnosis by a specialist before the index date)** | | |
| No | 9638 (96.1%) | 102360 (98.6%) |
| Yes | 395 (3.9%) | 1497 (1.4%) |
| **Bipolar disorder (any diagnosis by a specialist before the index date)** | | |
| No | 9598 (95.7%) | 100181 (96.5%) |
| Yes | 435 (4.3%) | 3676 (3.5%) |
| **Substance use disorder (any diagnosis by a specialist before the index date)** | | |
| No | 7695 (76.7%) | 82142 (79.1%) |
| Yes | 2338 (23.3%) | 21715 (20.9%) |
| **Eating disorder (any diagnosis by a specialist before the index date)** | | |
| No | 9906 (98.7%) | 100345 (96.6%) |
| Yes | 127 (1.3%) | 3512 (3.4%) |
| **Psychotic disorder (any diagnosis by a specialist before the index date)** | | |
| No | 9009 (89.8%) | 98805 (95.1%) |
| Yes | 1024 (10.2%) | 5052 (4.9%) |
| **Any of the following genera medical diseases (any diagnosis by a specialist before the index date)** | | |
| No | 6448 (64.3%) | 62345 (60.0%) |
| Yes | 3585 (35.7%) | 41512 (40.0%) |
| **Type 1/2 diabetes (any diagnosis by a specialist before the index date)** | | |
| No | 9860 (98.3%) | 102075 (98.3%) |
| Yes | 173 (1.7%) | 1782 (1.7%) |
| **Any cardiovascular disease (any diagnosis by a specialist before the index date)** | | |
| No | 9473 (94.4%) | 100149 (96.4%) |
| Yes | 560 (5.6%) | 3708 (3.6%) |
| **Obesity (any diagnosis by a specialist before the index date)** | | |
| No | 9798 (97.7%) | 100919 (97.2%) |
| Yes | 235 (2.3%) | 2938 (2.8%) |
| **Sleep disorder (any diagnosis by a specialist before the index date)** | | |
| No | 9721 (96.9%) | 101494 (97.7%) |
| Yes | 312 (3.1%) | 2363 (2.3%) |
| **Chronic pain (any diagnosis by a specialist before the index date)** | | |
| No | 7169 (71.5%) | 68759 (66.2%) |
| Yes | 2864 (28.5%) | 35098 (33.8%) |
| **Malformation (any diagnosis by a specialist before the index date)** | | |
| No | 9441 (94.1%) | 97395 (93.8%) |
| Yes | 592 (5.9%) | 6462 (6.2%) |
| **Injury (any diagnosis by a specialist before the index date)** | | |
| No | 6340 (63.2%) | 61023 (58.8%) |
| Yes | 3693 (36.8%) | 42834 (41.2%) |
| **Traumatic brain injury (any diagnosis by a specialist before the index date)** | | |
| No | 8641 (86.1%) | 89164 (85.9%) |
| Yes | 1392 (13.9%) | 14693 (14.1%) |
| **Accident (any diagnosis by a specialist before the index date)** | | |
| No | 5403 (53.9%) | 55546 (53.5%) |
| Yes | 4630 (46.1%) | 48311 (46.5%) |
| **Assault/victimization (any diagnosis by a specialist before the index date)** | | |
| No | 9379 (93.5%) | 95985 (92.4%) |
| Yes | 654 (6.5%) | 7872 (7.6%) |
| **Ever divorced before the year of the index date** |  |  |
| No | 9323 (92.9%) | 97982 (94.3%) |
| Yes | 709 (7.1%) | 5874 (5.7%) |
| Unknown | 1 (0.0%) | 1 (0.0%) |
| **Ever experienced spousal death before the index date** |  |  |
| No | 10015 (99.8%) | 103741 (99.9%) |
| Yes | 17 (0.2%) | 115 (0.1%) |
| Unknown | 1 (0.0%) | 1 (0.0%) |
| **Death of any parent before the index date** |  |  |
| No | 7692 (76.7%) | 88366 (85.1%) |
| Yes | 2341 (23.3%) | 15491 (14.9%) |
| **Death of any parent by suicide before the index date** |  |  |
| No | 9667 (96.4%) | 101329 (97.6%) |
| Yes | 366 (3.6%) | 2528 (2.4%) |
| **Suicide attempt of any parent before the index date** |  |  |
| No | 8728 (87.0%) | 88933 (85.6%) |
| Yes | 1305 (13.0%) | 14924 (14.4%) |
| **Any mental disorder in any parent (any diagnosis by a specialist before the index date)** | | |
| No | 7670 (76.4%) | 77321 (74.4%) |
| Yes | 2363 (23.6%) | 26536 (25.6%) |
| **Any substance use disorder in any parent (any diagnosis by a specialist before the index date)** | | |
| No | 8536 (85.1%) | 85829 (82.6%) |
| Yes | 1497 (14.9%) | 18028 (17.4%) |
| **Death of any child before the index date** |  |  |
| No | 9976 (99.4%) | 103500 (99.7%) |
| Yes | 57 (0.6%) | 357 (0.3%) |
| **Method used in initial suicide attempt** |  |  |
| Drug/poison | 2594 (25.9%) | 79088 (76.2%) |
| Firearm | 943 (9.4%) | 592 (0.6%) |
| Gas | 444 (4.4%) | 488 (0.5%) |
| Heat | 109 (1.1%) | 237 (0.2%) |
| Jump/crash | 1405 (14.0%) | 3687 (3.6%) |
| Object | 175 (1.7%) | 11829 (11.4%) |
| Other | 422 (4.2%) | 5825 (5.6%) |
| Suffocation | 3941 (39.3%) | 2111 (2.0%) |
| **Lethal method of attempt** |  |  |
| No | 3739 (37.3%) | 97440 (93.8%) |
| Yes | 6294 (62.7%) | 6417 (6.2%) |

*Proportion was calculated among the those who died or survived the initial suicide attempt (i.e., by column)*

*^a^Definition in Table S4*

#### **Table S7: Odds ratios of suicide attempts associated with potential risk factors**

|  | **All sexes** | | | | | **Male** | | **Female** | |
| --- | --- | --- | --- | --- | --- | --- | --- | --- | --- |
| **Risk factors** | **Cases** (suicide attempt) | **Controls**  (No suicide attempt) | **Model 1**  OR (95% CI) | | **Model 2**  OR (95% CI) | **Model 1**  OR (95% CI) | **Model 2**  OR (95% CI) | **Model 1**  OR (95% CI) | **Model 2**  OR (95% CI) |
| **Socio-demographic factors** | | | | | | | | | |
| **Educational attainment before the year of the date of initial suicide attempt** | | | | | | | | | |
| Primary school (<9y) | 37122 (32.6%) | 114833 (20.2%) | | 1 | 1 | 1 | 1 | 1 | 1 |
| Secondary school (9-12y) | 40531 (35.6%) | 212877 (37.4%) | | 0.34  (0.33, 0.34) | 0.38*  (0.37, 0.39) | 0.33  (0.33, 0.34) | 0.38*  (0.37, 0.39) | 0.34  (0.33, 0.35) | 0.38*  (0.37, 0.39) |
| Post-secondary (>12y) | 9950 (8.7%) | 114252 (20.1%) | | 0.14  (0.13, 0.14) | 0.16*  (0.16, 0.17) | 0.13  (0.12, 0.13) | 0.15*  (0.14, 0.15) | 0.15  (0.14, 0.15) | 0.17*  (0.17, 0.18) |
| Unknown | 26287 (23.1%) | 127488 (22.4%) | | 0.93  (0.89, 0.97) | 0.94  (0.9, 0.99) | 0.92  (0.86, 0.98) | 0.9  (0.83, 0.96) | 0.93  (0.88, 0.99) | 0.98  (0.92, 1.04) |
| **Disposable income before the year of the date of initial suicide attempt, rank in tertile by year^a^** | | | | | | | | | |
| Low | 38238 (33.6%) | 117326 (20.6%) | 1 | | 1 | 1 | 1 | 1 | 1 |
| Medium | 22120 (19.4%) | 125155 (22.0%) | 0.52  (0.51, 0.53) | | 0.56*  (0.55, 0.58) | 0.51  (0.49, 0.52) | 0.54*  (0.53, 0.56) | 0.54  (0.52, 0.55) | 0.58*  (0.57, 0.6) |
| High | 16448 (14.4%) | 140672 (24.7%) | 0.34  (0.33, 0.34) | | 0.41*  (0.4, 0.42) | 0.32  (0.31, 0.33) | 0.38*  (0.37, 0.39) | 0.36  (0.35, 0.37) | 0.43*  (0.42, 0.45) |
| Unknown | 37084 (32.6%) | 186297 (32.7%) | 0.53  (0.5, 0.57) | | 0.55*  (0.51, 0.59) | 0.55  (0.48, 0.62) | 0.56*  (0.5, 0.64) | 0.53  (0.49, 0.58) | 0.54*  (0.49, 0.59) |
| **Marital status in the year before date of initial suicide attempt** | | | | | | | | | |
| Unmarried | 99408 (87.3%) | 485848 (85.3%) | 1 | | 1 | 1 | 1 | 1 | 1 |
| Married | 8801 (7.7%) | 71592 (12.6%) | 0.57  (0.55, 0.58) | | 0.63*  (0.61, 0.65) | 0.48  (0.46, 0.5) | 0.53*  (0.51, 0.56) | 0.67  (0.64, 0.69) | 0.73*  (0.71, 0.76) |
| Divorced | 5570 (4.9%) | 11327 (2.0%) | 2.29  (2.21, 2.38) | | 1.94*  (1.86, 2.01) | 1.92  (1.82, 2.03) | 1.79*  (1.69, 1.9) | 2.68  (2.55, 2.81) | 2.11*  (2, 2.22) |
| Widow(er) | 111 (0.1%) | 249 (0.0%) | 2.1  (1.68, 2.63) | | 1.94*  (1.52, 2.47) | 1.44  (0.92, 2.26) | 1.36  (0.84, 2.19) | 2.56  (1.97, 3.34) | 2.31*  (1.75, 3.06) |
| **Parental socio-demographic factors** | | | | | | | | | |
| **Highest education level of parents** | | | | | | | | | |
| Primary school (<9y) | 14711 (12.9%) | 58400 (10.3%) | | 1 | 1 | 1 | 1 | 1 | 1 |
| Secondary school (9-12y) | 54455 (47.8%) | 243370 (42.7%) | | 0.86  (0.84, 0.88) | 0.99  (0.97, 1.01) | 0.85  (0.82, 0.87) | 1  (0.97, 1.03) | 0.87  (0.84, 0.9) | 0.98  (0.96, 1.02) |
| Post-secondary (>12y) | 31046 (27.3%) | 201858 (35.4%) | | 0.58  (0.57, 0.6) | 0.85*  (0.83, 0.87) | 0.55  (0.53, 0.57) | 0.85*  (0.82, 0.88) | 0.62  (0.6, 0.64) | 0.85*  (0.83, 0.88) |
| Unknown | 13678 (12.0%) | 65822 (11.6%) | | 1.15  (1.08, 1.22) | 1.07  (1, 1.13) | 1.13  (1.04, 1.23) | 1.05  (0.96, 1.15) | 1.16  (1.07, 1.26) | 1.08  (0.99, 1.17) |
| **Highest income level of parents (tertile rank by year)** | | | | | | | | | |
| Low | 24509 (21.5%) | 92466 (16.2%) | 1 | | 1 | 1 | 1 | 1 | 1 |
| Medium | 30747 (27.0%) | 150701 (26.5%) | 0.75  (0.74, 0.76) | | 0.96*  (0.94, 0.98) | 0.74  (0.72, 0.76) | 0.99  (0.97, 1.03) | 0.76  (0.74, 0.78) | 0.94*  (0.91, 0.96) |
| High | 28693 (25.2%) | 178802 (31.4%) | 0.58  (0.56, 0.59) | | 0.87*  (0.86, 0.89) | 0.54  (0.52, 0.55) | 0.88*  (0.85, 0.9) | 0.61  (0.6, 0.63) | 0.87*  (0.85, 0.9) |
| Unknown | 29941 (26.3%) | 147481 (25.9%) | 1.02  (0.96, 1.08) | | 1.17*  (1.1, 1.24) | 1.05  (0.97, 1.15) | 1.23*  (1.13, 1.35) | 0.98  (0.91, 1.07) | 1.11  (1.02, 1.21) |
| **Psychiatric disorders** | | | | | | | | | |
| **Any of the studied (below) psychiatric disorders (any diagnosis by a specialist before the date of the initial suicide attempt)** | | | | | | | | | |
| No | 71484 (62.8%) | 544085 (95.5%) | 1 | | 1 | 1 | 1 | 1 | 1 |
| Yes | 42406 (37.2%) | 25365 (4.5%) | 16.68  (16.33, 17.03) | | 13.82*  (13.52, 14.13) | 17.49  (16.95, 18.04) | 13.66*  (13.23, 14.11) | 16  (15.54, 16.47) | 13.96*  (13.55, 14.38) |
| **Major depressive disorder (any diagnosis by a specialist before the date of the initial suicide attempt)** | | | | | | | | | |
| No | 90885 (79.8%) | 557535 (97.9%) | 1 | | 1 | 1 | 1 | 1 | 1 |
| Yes | 23005 (20.2%) | 11915 (2.1%) | 14.56  (14.18, 14.95) | | 12.85*  (12.49, 13.21) | 13.55  (13, 14.11) | 11.36*  (10.88, 11.87) | 15.32  (14.79, 15.86) | 13.9*  (13.41, 14.42) |
| **Schizophrenia (any diagnosis by a specialist before the date of the initial suicide attempt)** | | | | | | | | | |
| No | 111998 (98.3%) | 568666 (99.9%) | 1 | | 1 | 1 | 1 | 1 | 1 |
| Yes | 1892 (1.7%) | 784 (0.1%) | 12.41  (11.4, 13.5) | | 9.24*  (8.46, 10.1) | 11.18  (10.09, 12.39) | 8.16*  (7.31, 9.1) | 15.18  (13.09, 17.61) | 11.66*  (9.99, 13.61) |
| **Bipolar disorder (any diagnosis by a specialist before the date of initial suicide attempt)** | | | | | | | | | |
| No | 109779 (96.4%) | 567731 (99.7%) | 1 | | 1 | 1 | 1 | 1 | 1 |
| Yes | 4111 (3.6%) | 1719 (0.3%) | 12.84  (12.11, 13.61) | | 11.52* (10.83, 12.25) | 11.8  (10.79, 12.91) | 10.59*  (9.62, 11.65) | 13.62  (12.62, 14.7) | 12.18* (11.24, 13.2) |
| **Substance use disorder (any diagnosis by a specialist before the date of the initial suicide attempt)** | | | | | | | | | |
| No | 89837 (78.9%) | 558218 (98.0%) | 1 | | 1 | 1 | 1 | 1 | 1 |
| Yes | 24053 (21.1%) | 11232 (2.0%) | 15.26 (14.86, 15.66) | | 11.49* (11.18, 11.81) | 16.62  (16.04, 17.22) | 12.05*  (11.62, 12.5) | 13.7  (13.18, 14.24) | 10.86* (10.43, 11.3) |
| **Eating disorder (any diagnosis by a specialist before the date of the initial suicide attempt)** | | | | | | | | | |
| No | 110251 (96.8%) | 566083 (99.4%) | 1 | | 1 | 1 | 1 | 1 | 1 |
| Yes | 3639 (3.2%) | 3367 (0.6%) | 5.77  (5.5, 6.06) | | 5.83*  (5.54, 6.13) | 4.06 (3.43, 4.8) | 3.54* (2.96, 4.23) | 5.96 (5.66, 6.27) | 6.04* (5.73, 6.37) |
| **Psychotic disorder (any diagnosis by a specialist before the date of the initial suicide attempt)** | | | | | | | | | |
| No | 107814 (94.7%) | 567188 (99.6%) | 1 | | 1 | 1 | 1 | 1 | 1 |
| Yes | 6076 (5.3%) | 2262 (0.4%) | 14.62  (13.91, 15.37) | | 11.76*  (11.16, 12.4) | 13.84  (12.97, 14.77) | 10.75*  (10.03, 11.51) | 15.83  (14.62, 17.13) | 13.31* (12.26, 14.46) |
| **General medical diseases** | | | | | | | | | |
| **Any of the studied (below) general medical diseases (any diagnosis by a specialist before the date of the initial suicide attempt)** | | | | | | | | | |
| No | 68793 (60.4%) | 414631 (72.8%) | 1 | | 1 | 1 | 1 | 1 | 1 |
| Yes | 45097 (39.6%) | 154819 (27.2%) | 2.04  (2.01, 2.07) | | 1.86*  (1.83, 1.89) | 1.87  (1.83, 1.91) | 1.71*  (1.67, 1.75) | 2.2  (2.16, 2.25) | 2.01*  (1.96, 2.05) |
| **Type 1/2 diabetes (any diagnosis by a specialist before the date of the initial suicide attempt)** | | | | | | | | | |
| No | 111935 (98.3%) | 564706 (99.2%) | 1 | | 1 | 1 | 1 | 1 | 1 |
| Yes | 1955 (1.7%) | 4744 (0.8%) | 2.08  (1.98, 2.2) | | 1.88*  (1.78, 1.99) | 2.21  (2.06, 2.38) | 1.98*  (1.83, 2.14) | 1.95  (1.8, 2.11) | 1.78*  (1.64, 1.93) |
| **Cardiovascular disease (any diagnosis by a specialist before the date of the initial suicide attempt)** | | | | | | | | | |
| No | 109622 (96.3%) | 558440 (98.1%) | 1 | | 1 | 1 | 1 | 1 | 1 |
| Yes | 4268 (3.7%) | 11010 (1.9%) | 2.03  (1.96, 2.11) | | 1.84*  (1.77, 1.91) | 2.11  (2.01, 2.22) | 1.93*  (1.83, 2.03) | 1.93  (1.82, 2.04) | 1.74*  (1.64, 1.84) |
| **Obesity (any diagnosis by a specialist before the date of the initial suicide attempt)** | | | | | | | | | |
| No | 110717 (97.2%) | 563231 (98.9%) | 1 | | 1 | 1 | 1 | 1 | 1 |
| Yes | 3173 (2.8%) | 6219 (1.1%) | 2.66  (2.55, 2.78) | | 2.03*  (1.94, 2.13) | 2.3  (2.13, 2.49) | 1.79*  (1.65, 1.95) | 2.84  (2.7, 3) | 2.18*  (2.06, 2.3) |
| **Sleep disorder (any diagnosis by a specialist before the date of the initial suicide attempt)** | | | | | | | | | |
| No | 111215 (97.7%) | 565796 (99.4%) | 1 | | 1 | 1 | 1 | 1 | 1 |
| Yes | 2675 (2.3%) | 3654 (0.6%) | 3.82  (3.63, 4.02) | | 3.22*  (3.05, 3.4) | 3.34  (3.12, 3.57) | 2.8*  (2.6, 3.01) | 4.62  (4.27, 5.01) | 3.92*  (3.6, 4.26) |
| **Chronic pain^b^ (any diagnosis by a specialist before the date of the initial suicide attempt)** | | | | | | | | | |
| No | 75928 (66.7%) | 444052 (78.0%) | 1 | | 1 | 1 | 1 | 1 | 1 |
| Yes | 37962 (33.3%) | 125398 (22.0%) | 2.11  (2.08, 2.15) | | 1.94*  (1.9, 1.97) | 1.93  (1.89, 1.98) | 1.78*  (1.74, 1.83) | 2.27  (2.22, 2.32) | 2.07*  (2.03, 2.12) |
| **Malformation (any diagnosis by a specialist before the date of the initial suicide attempt)** | | | | | | | | | |
| No | 106836 (93.8%) | 540677 (94.9%) | 1 | | 1 | 1 | 1 | 1 | 1 |
| Yes | 7054 (6.2%) | 28773 (5.1%) | 1.24  (1.21, 1.28) | | 1.13*  (1.1, 1.17) | 1.19  (1.15, 1.24) | 1.08*  (1.04, 1.12) | 1.3  (1.25, 1.35) | 1.19*  (1.14, 1.24) |
| **Adverse life events** | | | | | | | | | |
| **Injury (any diagnosis by a specialist before the date of the initial suicide attempt)** | | | | | | | | | |
| No | 67363 (59.1%) | 421164 (74.0%) | 1 | | 1 | 1 | 1 | 1 | 1 |
| Yes | 46527 (40.9%) | 148286 (26.0%) | 2.61  (2.57, 2.65) | | 2.39*  (2.35, 2.43) | 2.61  (2.55, 2.67) | 2.34*  (2.28, 2.4) | 2.61  (2.55, 2.66) | 2.43*  (2.37, 2.49) |
| **Traumatic brain injury (any diagnosis by a specialist before the date of the initial suicide attempt)** | | | | | | | | | |
| No | 97805 (85.9%) | 531009 (93.2%) | 1 | | 1 | 1 | 1 | 1 | 1 |
| Yes | 16085 (14.1%) | 38441 (6.8%) | 2.29  (2.25, 2.34) | | 2.1*  (2.06, 2.15) | 2.34  (2.28, 2.4) | 2.09*  (2.03, 2.15) | 2.24  (2.17, 2.31) | 2.11*  (2.05, 2.18) |
| **Accident (any diagnosis by a specialist before the date of the initial suicide attempt)** | | | | | | | | | |
| No | 60949 (53.5%) | 395439 (69.4%) | 1 | | 1 | 1 | 1 | 1 | 1 |
| Yes | 52941 (46.5%) | 174011 (30.6%) | 2.2  (2.17, 2.23) | | 2.05*  (2.02, 2.08) | 2.23  (2.18, 2.27) | 2.03*  (1.99, 2.07) | 2.18  (2.14, 2.22) | 2.06*  (2.02, 2.1) |
| **Assault/victimization (any diagnosis by a specialist before the date of the initial suicide attempt)** | | | | | | | | | |
| No | 105364 (92.5%) | 561240 (98.6%) | 1 | | 1 | 1 | 1 | 1 | 1 |
| Yes | 8526 (7.5%) | 8210 (1.4%) | 5.83  (5.65, 6.02) | | 4.38*  (4.23, 4.53) | 5.09  (4.9, 5.3) | 3.89*  (3.73, 4.05) | 7.7  (7.27, 8.15) | 5.55*  (5.22, 5.89) |
| **Ever divorced before the year of the initial suicide attempt** | | | | | | | | | |
| No | 107305 (94.2%) | 554387 (97.4%) | 1 | | 1 | 1 | 1 | 1 | 1 |
| Yes | 6583 (5.8%) | 14622 (2.6%) | 2.68 (2.6, 2.77) | | 2.18* (2.11, 2.26) | 2.28 (2.17, 2.4) | 2.03* (1.93, 2.14) | 3.05 (2.92, 3.18) | 2.33* (2.23, 2.44) |
| **Ever experienced spousal death before the date of the initial suicide attempt** | | | | | | | | | |
| No | 113756 (99.9%) | 568716 (99.9%) | 1 | | 1 | 1 | 1 | 1 | 1 |
| Yes | 132 (0.1%) | 293 (0.1%) | 2.26  (1.84, 2.77) | | 2.02*  (1.62, 2.52) | 1.72  (1.15, 2.57) | 1.5  (0.97, 2.3) | 2.51  (1.97, 3.2) | 2.28*  (1.76, 2.94) |
| **Death of any parent before the date of the initial suicide attempt** | | | | | | | | | |
| No | 96058 (84.3%) | 508425 (89.3%) | 1 | | 1 | 1 | 1 | 1 | 1 |
| Yes | 17832 (15.7%) | 61025 (10.7%) | 1.68  (1.64, 1.71) | | 1.47*  (1.44, 1.5) | 1.7  (1.66, 1.75) | 1.47*  (1.43, 1.52) | 1.65  (1.6, 1.7) | 1.47*  (1.43, 1.51) |
| **Death of any parent by suicide before the date of the initial suicide attempt** | | | | | | | | | |
| No | 110996 (97.5%) | 563938 (99.0%) | 1 | | 1 | 1 | 1 | 1 | 1 |
| Yes | 2894 (2.5%) | 5512 (1.0%) | 2.67  (2.55, 2.8) | | 2.33*  (2.22, 2.45) | 2.76  (2.58, 2.94) | 2.32*  (2.17, 2.49) | 2.6  (2.43, 2.77) | 2.34*  (2.18, 2.5) |
| **Self-harm in any parent before the date of the initial suicide attempt** | | | | | | | | | |
| No | 97661 (85.8%) | 538443 (94.6%) | 1 | | 1 | 1 | 1 | 1 | 1 |
| Yes | 16229 (14.2%) | 31007 (5.4%) | 2.91  (2.85, 2.97) | | 2.43*  (2.38, 2.48) | 2.91  (2.83, 3) | 2.35*  (2.27, 2.42) | 2.91  (2.83, 2.99) | 2.5*  (2.43, 2.58) |
| **Any psychiatric disorder in any parent (any diagnosis by a specialist before the date of the initial suicide attempt)** | | | | | | | | | |
| No | 84991 (74.6%) | 501042 (88.0%) | 1 | | 1 | 1 | 1 | 1 | 1 |
| Yes | 28899 (25.4%) | 68408 (12.0%) | 2.55  (2.51, 2.59) | | 2.14*  (2.1, 2.17) | 2.56  (2.5, 2.62) | 2.07*  (2.02, 2.13) | 2.54  (2.49, 2.6) | 2.19*  (2.14, 2.24) |
| **Any substance use disorder in any parent (any diagnosis by a specialist before the date of the initial suicide attempt)** | | | | | | | | | |
| No | 94365 (82.9%) | 530625 (93.2%) | 1 | | 1 | 1 | 1 | 1 | 1 |
| Yes | 19525 (17.1%) | 38825 (6.8%) | 2.85  (2.8, 2.91) | | 2.29*  (2.25, 2.34) | 2.95  (2.87, 3.04) | 2.28*  (2.22, 2.35) | 2.77  (2.7, 2.84) | 2.3*  (2.24, 2.36) |
| **Death of any child before the date of the initial suicide attempt** | | | | | | | | | |
| No | 113476 (99.6%) | 568371 (99.8%) | 1 | | 1 | 1 | 1 | 1 | 1 |
| Yes | 414 (0.4%) | 1079 (0.2%) | 1.93  (1.72, 2.16) | | 1.45*  (1.28, 1.64) | 1.74  (1.45, 2.08) | 1.41  (1.17, 1.72) | 2.08  (1.79, 2.41) | 1.48*  (1.27, 1.73) |

*Odds ratios were estimated using the conditional logistic regression, conditioning on matching factors (i.e., sex and birthyear)*

*Model 1: no covariate*

*Model 2: adjusted for education and income*

*Asterisks (*) indicate OR was statistically significantly different from 1 (p-value ≤ 4.17*10^-4^, Bonferroni-corrected p-value for 120 hypothesis tests). We only tested ORs of model 2 for all population, male and female*

*^a^Disposable income per consumption unit within family, rank in tertile by year among the included individuals*

*^b^Chronic pain included migraine, sexual pain, headache, temporomandibular joint disorders, gastro-esophageal reflux disease, irritable bowel disease (IBS), fibromyalgia, interstitial cystitis, chronic fatigue syndrome, back/neck pain, arthritis/osteoarthritis/joint pain, other musculoskeletal/connective tissue pain, and other painful conditions.*

#### **Table S8: Odds ratios of any self-harm associated with potential risk factors**

|  | **All sexes** | | | | | | | **Male** | | | | **Female** | | | |
| --- | --- | --- | --- | --- | --- | --- | --- | --- | --- | --- | --- | --- | --- | --- | --- |
| **Risk factors** | **Cases** (Self-harm) | **Controls**  (No self-harm) | | **Model 1**  OR (95% CI) | | **Model 2**  OR (95% CI) | | **Model 1**  OR (95% CI) | | **Model 2**  OR (95% CI) | | **Model 1**  OR (95% CI) | | **Model 2**  OR (95% CI) | |
| **Socio-demographic factors** | | | | | | | | | | | | | | | |
| **Educational attainment before the year of the initial self-harm** | | | | | | | | | | | | | | | |
| Primary school (<9y) | 45850 (29.1%) | 154231 (19.6%) | | 1 | | 1 | | 1 | | 1 | | 1 | | 1 | |
| Secondary school (9-12y) | 59494 (37.7%) | 300779 (38.1%) | | 0.42  (0.41, 0.43) | | 0.46  (0.45, 0.47) | | 0.44  (0.43, 0.45) | | 0.48 (0.47, 0.5) | | 0.39  (0.38, 0.4) | | 0.43  (0.41, 0.44) | |
| Post-secondary (>12y) | 18263 (11.6%) | 167363 (21.2%) | | 0.21  (0.21, 0.22) | | 0.24  (0.23, 0.24) | | 0.21  (0.2, 0.22) | | 0.23  (0.23, 0.24) | | 0.21  (0.2, 0.22) | | 0.24  (0.23, 0.25) | |
| Unknown | 34172 (21.7%) | 166522 (21.1%) | | 0.96  (0.92, 0.99) | | 0.97  (0.93, 1.02) | | 0.93  (0.88, 0.99) | | 0.93  (0.87, 0.98) | | 0.97  (0.92, 1.03) | | 1.02  (0.96, 1.08) | |
| **Disposable income before the year of the initial self-harm, rank in tertile by year^a^** | | | | | | | | | | | | | | | |
| Low | 49651 (31.5%) | | 172289 (21.8%) | | 1 | | 1 | | 1 | | 1 | | 1 | | 1 |
| Medium | 34167 (21.7%) | | 185198 (23.5%) | | 0.62  (0.61, 0.63) | | 0.66  (0.65, 0.68) | | 0.64  (0.63, 0.66) | | 0.68  (0.66, 0.69) | | 0.6  (0.59, 0.62) | | 0.65  (0.64, 0.66) |
| High | 29666 (18.8%) | | 209045 (26.5%) | | 0.47  (0.46, 0.48) | | 0.55  (0.54, 0.56) | | 0.49  (0.48, 0.5) | | 0.56  (0.55, 0.57) | | 0.45  (0.44, 0.46) | | 0.53  (0.52, 0.54) |
| Unknown | 44295 (28.1%) | | 222363 (28.2%) | | 0.62  (0.58, 0.66) | | 0.62  (0.58, 0.67) | | 0.63  (0.57, 0.7) | | 0.64  (0.58, 0.71) | | 0.6  (0.56, 0.66) | | 0.6  (0.55, 0.66) |
| **Marital status in the year before date of the initial self-harm** | | | | | | | | | | | | | | | |
| Unmarried | 135214 (85.7%) | 663298 (84.1%) | | 1 | | 1 | | 1 | | 1 | | 1 | | 1 | |
| Married | 15299 (9.7%) | 107266 (13.6%) | | 0.67  (0.65, 0.68) | | 0.73  (0.72, 0.75) | | 0.63  (0.61, 0.64) | | 0.69  (0.67, 0.71) | | 0.72  (0.7, 0.74) | | 0.78  (0.76, 0.8) | |
| Divorced | 7110 (4.5%) | 17310 (2.2%) | | 1.93  (1.87, 1.99) | | 1.68  (1.63, 1.74) | | 1.65  (1.58, 1.73) | | 1.54  (1.47, 1.62) | | 2.24  (2.14, 2.33) | | 1.83  (1.75, 1.91) | |
| Widow(er) | 155 (0.1%) | 430 (0.1%) | | 1.7  (1.41, 2.04) | | 1.56  (1.29, 1.9) | | 1.17  (0.84, 1.64) | | 1.16  (0.82, 1.65) | | 2.11  (1.69, 2.63) | | 1.85  (1.47, 2.33) | |
| **Parental socio-demographic factors** | | | | | | | | | | | | | | | |
| **Highest education level of parents** | | | | | | | | | | | | | | | |
| Primary school (<9y) | 19561 (12.4%) | 81583 (10.3%) | | 1 | | 1 | | 1 | | 1 | | 1 | | 1 | |
| Secondary school (9-12y) | 77086 (48.9%) | 348479 (44.2%) | | 0.9  (0.88, 0.91) | | 1.01  (1, 1.03) | | 0.88  (0.86, 0.91) | | 1.01  (0.99, 1.04) | | 0.91  (0.89, 0.93) | | 1.02  (0.99, 1.04) | |
| Post-secondary (>12y) | 47065 (29.8%) | 291132 (36.9%) | | 0.65  (0.64, 0.66) | | 0.88  (0.86, 0.9) | | 0.62  (0.6, 0.64) | | 0.86  (0.84, 0.89) | | 0.68  (0.66, 0.7) | | 0.9  (0.87, 0.92) | |
| Unknown | 14067 (8.9%) | 67701 (8.6%) | | 1.12  (1.06, 1.17) | | 1.06  (1.01, 1.12) | | 1.08  (1, 1.16) | | 1.04  (0.97, 1.13) | | 1.16  (1.08, 1.25) | | 1.08  (1, 1.17) | |
| **Highest income level of parents (tertile rank by year) ^a^** | | | | | | | | | | | | | | | |
| Low | 35206 (22.3%) | 144733 (18.3%) | | 1 | | 1 | | 1 | | 1 | | 1 | | 1 | |
| Medium | 46292 (29.3%) | 226210 (28.7%) | | 0.83  (0.81, 0.84) | | 0.98  (0.97, 1) | | 0.84  (0.82, 0.86) | | 1.01  (0.99, 1.03) | | 0.81  (0.79, 0.83) | | 0.96  (0.94, 0.98) | |
| High | 45803 (29.0%) | 267872 (34.0%) | | 0.68  (0.67, 0.69) | | 0.91  (0.9, 0.93) | | 0.66  (0.65, 0.68) | | 0.92  (0.9, 0.94) | | 0.69  (0.67, 0.71) | | 0.91  (0.89, 0.94) | |
| Unknown | 30478 (19.3%) | 150080 (19.0%) | | 1.03  (0.98, 1.09) | | 1.1  (1.05, 1.16) | | 1.06  (0.99, 1.14) | | 1.14  (1.06, 1.22) | | 1  (0.94, 1.08) | | 1.07  (0.99, 1.15) | |
| **Psychiatric disorders** | | | | | | | | | | | | | | | |
| **Any of the following psychiatric disorders (any diagnosis by a specialist before the date of the initial self-harm)** | | | | | | | | | | | | | | | |
| No | 111647 (70.8%) | 752094 (95.3%) | | 1 | | 1 | | 1 | | 1 | | 1 | | 1 | |
| Yes | 46132 (29.2%) | 36801 (4.7%) | | 9.83  (9.66, 9.99) | | 8.32  (8.18, 8.47) | | 9.27  (9.06, 9.5) | | 7.63  (7.45, 7.82) | | 10.42  (10.17, 10.67) | | 9.1  (8.88, 9.33) | |
| **Major depressive disorder (any diagnosis by a specialist before the date of the initial self-harm)** | | | | | | | | | | | | | | | |
| No | 133211 (84.4%) | 771472 (97.8%) | | 1 | | 1 | | 1 | | 1 | | 1 | | 1 | |
| Yes | 24568 (15.6%) | 17423 (2.2%) | | 9.29  (9.09, 9.5) | | 8.04  (7.86, 8.22) | | 8.01  (7.75, 8.28) | | 6.7  (6.47, 6.93) | | 10.42  (10.12, 10.73) | | 9.22  (8.95, 9.51) | |
| **Schizophrenia (any diagnosis by a specialist before the date of the initial self-harm)** | | | | | | | | | | | | | | | |
| No | 155819 (98.8%) | 787686 (99.8%) | | 1 | | 1 | | 1 | | 1 | | 1 | | 1 | |
| Yes | 1960 (1.2%) | 1209 (0.2%) | | 8.25  (7.68, 8.87) | | 6.19  (5.74, 6.67) | | 7.37  (6.75, 8.05) | | 5.45  (4.97, 5.98) | | 10.3  (9.08, 11.7) | | 7.98  (7, 9.1) | |
| **Bipolar disorder (any diagnosis by a specialist before the date of the initial self-harm)** | | | | | | | | | | | | | | | |
| No | 153469 (97.3%) | 786402 (99.7%) | | 1 | | 1 | | 1 | | 1 | | 1 | | 1 | |
| Yes | 4310 (2.7%) | 2493 (0.3%) | | 9.06  (8.62, 9.53) | | 7.82  (7.42, 8.24) | | 7.98  (7.39, 8.61) | | 6.75  (6.23, 7.31) | | 9.96  (9.31, 10.65) | | 8.69  (8.11, 9.31) | |
| **Substance use disorder (any diagnosis by a specialist before the date of the initial self-harm)** | | | | | | | | | | | | | | | |
| No | 131568 (83.4%) | 772492 (97.9%) | | 1 | | 1 | | 1 | | 1 | | 1 | | 1 | |
| Yes | 26211 (16.6%) | 16403 (2.1%) | | 10.14  (9.92, 10.36) | | 7.94  (7.76, 8.12) | | 10.11  (9.83, 10.39) | | 7.84  (7.62, 8.07) | | 10.19  (9.85, 10.55) | | 8.2  (7.92, 8.49) | |
| **Eating disorder (any diagnosis by a specialist before the date of the initial self-harm)** | | | | | | | | | | | | | | | |
| No | 153833 (97.5%) | 784386 (99.4%) | | 1 | | 1 | | 1 | | 1 | | 1 | | 1 | |
| Yes | 3946 (2.5%) | 4509 (0.6%) | | 4.61  (4.41, 4.82) | | 4.49  (4.29, 4.7) | | 2.48  (2.16, 2.86) | | 2.22  (1.92, 2.57) | | 4.96  (4.74, 5.2) | | 4.88  (4.65, 5.12) | |
| **Psychotic disorder (any diagnosis by a specialist before the date of the initial self-harm)** | | | | | | | | | | | | | | | |
| No | 151451 (96.0%) | 785502 (99.6%) | | 1 | | 1 | | 1 | | 1 | | 1 | | 1 | |
| Yes | 6328 (4.0%) | 3393 (0.4%) | | 9.86  (9.45, 10.29) | | 7.91  (7.57, 8.27) | | 9  (8.52, 9.5) | | 7.04  (6.65, 7.45) | | 11.35  (10.59, 12.16) | | 9.49  (8.84, 10.19) | |
| **General medical diseases** | | | | | | | | | | | | | | | |
| **Any of the following general medical diseases (any diagnosis by a specialist before the date of the initial self-harm)** | | | | | | | | | | | | | | | |
| No | 93272 (59.1%) | 558917 (70.8%) | | 1 | | 1 | | 1 | | 1 | | 1 | | 1 | |
| Yes | 64507 (40.9%) | 229978 (29.2%) | | 1.88  (1.86, 1.91) | | 1.75  (1.73, 1.78) | | 1.72  (1.69, 1.75) | | 1.61  (1.58, 1.64) | | 2.08  (2.05, 2.12) | | 1.92  (1.89, 1.96) | |
| **Type 1/2 diabetes (any diagnosis by a specialist before the date of the initial self-harm)** | | | | | | | | | | | | | | | |
| No | 155303 (98.4%) | 782265 (99.2%) | | 1 | | 1 | | 1 | | 1 | | 1 | | 1 | |
| Yes | 2476 (1.6%) | 6630 (0.8%) | | 1.89  (1.8, 1.98) | | 1.75  (1.67, 1.84) | | 1.87  (1.76, 1.99) | | 1.75  (1.64, 1.87) | | 1.91  (1.78, 2.05) | | 1.75  (1.63, 1.88) | |
| **Cardiovascular disease (any diagnosis by a specialist before the date of the initial self-harm)** | | | | | | | | | | | | | | | |
| No | 151960 (96.3%) | 772339 (97.9%) | | 1 | | 1 | | 1 | | 1 | | 1 | | 1 | |
| Yes | 5819 (3.7%) | 16556 (2.1%) | | 1.83  (1.77, 1.88) | | 1.68  (1.62, 1.73) | | 1.79  (1.72, 1.86) | | 1.65  (1.58, 1.72) | | 1.88  (1.79, 1.97) | | 1.71  (1.63, 1.8) | |
| **Obesity (any diagnosis by a specialist before the date of the initial self-harm)** | | | | | | | | | | | | | | | |
| No | 153800 (97.5%) | 779517 (98.8%) | | 1 | | 1 | | 1 | | 1 | | 1 | | 1 | |
| Yes | 3979 (2.5%) | 9378 (1.2%) | | 2.19  (2.11, 2.27) | | 1.77  (1.7, 1.84) | | 1.99  (1.86, 2.12) | | 1.65  (1.54, 1.76) | | 2.3  (2.2, 2.41) | | 1.84  (1.75, 1.93) | |
| **Sleep disorder (any diagnosis by a specialist before the date of the initial self-harm)** | | | | | | | | | | | | | | | |
| No | 154451 (97.9%) | 783269 (99.3%) | | 1 | | 1 | | 1 | | 1 | | 1 | | 1 | |
| Yes | 3328 (2.1%) | 5626 (0.7%) | | 3.05  (2.92, 3.19) | | 2.66  (2.55, 2.79) | | 2.66  (2.52, 2.82) | | 2.34  (2.21, 2.48) | | 3.83  (3.57, 4.11) | | 3.28  (3.05, 3.54) | |
| **Chronic pain^b^ (any diagnosis by a specialist before the date of the initial self-harm)** | | | | | | | | | | | | | | | |
| No | 102789 (65.1%) | 601668 (76.3%) | | 1 | | 1 | | 1 | | 1 | | 1 | | 1 | |
| Yes | 54990 (34.9%) | 187227 (23.7%) | | 1.97  (1.94, 2) | | 1.84  (1.82, 1.87) | | 1.82  (1.78, 1.85) | | 1.71  (1.68, 1.74) | | 2.14  (2.1, 2.19) | | 1.99  (1.95, 2.03) | |
| **Malformation (any diagnosis by a specialist before the date of the initial self-harm)** | | | | | | | | | | | | | | | |
| No | 147919 (93.8%) | 746734 (94.7%) | | 1 | | 1 | | 1 | | 1 | | 1 | | 1 | |
| Yes | 9860 (6.2%) | 42161 (5.3%) | | 1.18 (1.16, 1.21) | | 1.1 (1.08, 1.13) | | 1.1 (1.07, 1.14) | | 1.03 (1, 1.07) | | 1.29 (1.25, 1.33) | | 1.2 (1.16, 1.24) | |
| **Adverse life events** | | | | | | | | | | | | | | | |
| **Injury (any diagnosis by a specialist before the date of the initial self-harm)** | | | | | | | | | | | | | | | |
| No | 90074 (57.1%) | 563524 (71.4%) | | 1 | | 1 | | 1 | | 1 | | 1 | | 1 | |
| Yes | 67705 (42.9%) | 225371 (28.6%) | | 2.32  (2.29, 2.35) | | 2.17  (2.14, 2.2) | | 2.24  (2.2, 2.28) | | 2.07  (2.04, 2.11) | | 2.41  (2.36, 2.46) | | 2.28  (2.24, 2.32) | |
| **Traumatic brain injury (any diagnosis by a specialist before the date of the initial self-harm)** | | | | | | | | | | | | | | | |
| No | 136504 (86.5%) | 733522 (93.0%) | | 1 | | 1 | | 1 | | 1 | | 1 | | 1 | |
| Yes | 21275 (13.5%) | 55373 (7.0%) | | 2.08  (2.04, 2.12) | | 1.94  (1.91, 1.98) | | 2.06  (2.02, 2.11) | | 1.9  (1.86, 1.95) | | 2.11  (2.05, 2.16) | | 2  (1.94, 2.05) | |
| **Accident (any diagnosis by a specialist before the date of the initial self-harm)** | | | | | | | | | | | | | | | |
| No | 82418 (52.2%) | 531826 (67.4%) | | 1 | | 1 | | 1 | | 1 | | 1 | | 1 | |
| Yes | 75361 (47.8%) | 257069 (32.6%) | | 2.07  (2.05, 2.1) | | 1.96  (1.94, 1.98) | | 2.04  (2.01, 2.07) | | 1.9  (1.87, 1.94) | | 2.11  (2.07, 2.15) | | 2.01  (1.98, 2.05) | |
| **Assault/victimization (any diagnosis by a specialist before the date of the initial self-harm)** | | | | | | | | | | | | | | | |
| No | 147569 (93.5%) | 776662 (98.4%) | | 1 | | 1 | | 1 | | 1 | | 1 | | 1 | |
| Yes | 10210 (6.5%) | 12233 (1.6%) | | 4.57  (4.45, 4.7) | | 3.59  (3.48, 3.69) | | 3.97  (3.84, 4.1) | | 3.18  (3.07, 3.29) | | 6.42  (6.1, 6.76) | | 4.79  (4.54, 5.05) | |
| **Ever divorced before the year of the date of the initial self-harm** | | | | | | | | | | | | | | | |
| No | 149232 (94.6%) | 765863 (97.1%) | | 1 | | 1 | | 1 | | 1 | | 1 | | 1 | |
| Yes | 8543 (5.4%) | 22430 (2.8%) | | 2.16  (2.1, 2.23) | | 1.83  (1.78, 1.89) | | 1.87  (1.79, 1.95) | | 1.68  (1.61, 1.76) | | 2.46  (2.37, 2.56) | | 1.97  (1.89, 2.05) | |
| **Ever experienced spousal death before the date of the initial self-harm** | | | | | | | | | | | | | | | |
| No | 157594 (99.9%) | 787787 (99.9%) | | 1 | | 1 | | 1 | | 1 | | 1 | | 1 | |
| Yes | 181 (0.1%) | 506 (0.1%) | | 1.79  (1.51, 2.12) | | 1.6  (1.34, 1.91) | | 1.39  (1.02, 1.89) | | 1.31  (0.95, 1.81) | | 2.03  (1.65, 2.49) | | 1.76  (1.42, 2.18) | |
| **Death of any parent before the date of the initial self-harm** | | | | | | | | | | | | | | | |
| No | 133967 (84.9%) | 698547 (88.5%) | | 1 | | 1 | | 1 | | 1 | | 1 | | 1 | |
| Yes | 23812 (15.1%) | 90348 (11.5%) | | 1.46  (1.44, 1.49) | | 1.32  (1.29, 1.34) | | 1.44  (1.41, 1.48) | | 1.29  (1.26, 1.32) | | 1.48  (1.44, 1.52) | | 1.35  (1.31, 1.38) | |
| **Death of any parent by suicide before the date of the initial self-harm** | | | | | | | | | | | | | | | |
| No | 154339 (97.8%) | 780820 (99.0%) | | 1 | | 1 | | 1 | | 1 | | 1 | | 1 | |
| Yes | 3440 (2.2%) | 8075 (1.0%) | | 2.16  (2.08, 2.25) | | 1.92  (1.84, 2) | | 2.09  (1.98, 2.21) | | 1.84  (1.74, 1.95) | | 2.25  (2.12, 2.39) | | 2.01  (1.9, 2.14) | |
| **Suicide attempt in any parent before the date of the initial self-harm** | | | | | | | | | | | | | | | |
| No | 136039 (86.2%) | 744150 (94.3%) | | 1 | | 1 | | 1 | | 1 | | 1 | | 1 | |
| Yes | 21740 (13.8%) | 44745 (5.7%) | | 2.67  (2.62, 2.72) | | 2.34  (2.29, 2.38) | | 2.61  (2.54, 2.67) | | 2.26  (2.2, 2.31) | | 2.74  (2.67, 2.81) | | 2.42  (2.36, 2.48) | |
| **Any psychiatric disorder in any parent (any diagnosis by a specialist before the date of the initial self-harm)** | | | | | | | | | | | | | | | |
| No | 122275 (77.5%) | 690810 (87.6%) | | 1 | | 1 | | 1 | | 1 | | 1 | | 1 | |
| Yes | 35504 (22.5%) | 98085 (12.4%) | | 2.07  (2.05, 2.1) | | 1.8  (1.78, 1.83) | | 1.97  (1.93, 2.01) | | 1.7  (1.67, 1.73) | | 2.19  (2.14, 2.23) | | 1.92  (1.88, 1.96) | |
| **Any substance use disorder in any parent (any diagnosis by a specialist before the date of the initial self-harm)** | | | | | | | | | | | | | | | |
| No | 134167 (85.0%) | 733127 (92.9%) | | 1 | | 1 | | 1 | | 1 | | 1 | | 1 | |
| Yes | 23612 (15.0%) | 55768 (7.1%) | | 2.33  (2.29, 2.36) | | 1.95  (1.92, 1.98) | | 2.26  (2.2, 2.31) | | 1.87  (1.82, 1.91) | | 2.4  (2.35, 2.46) | | 2.04  (1.99, 2.09) | |
| **Death of any child before the date of the initial self-harm** | | | | | | | | | | | | | | | |
| No | 157242 (99.7%) | 787241 (99.8%) | | 1 | | 1 | | 1 | | 1 | | 1 | | 1 | |
| Yes | 537 (0.3%) | 1654 (0.2%) | | 1.63  (1.48, 1.8) | | 1.37  (1.24, 1.52) | | 1.46  (1.26, 1.69) | | 1.28  (1.1, 1.49) | | 1.79  (1.57, 2.05) | | 1.45  (1.26, 1.66) | |

*Odds ratios were estimated using the conditional logistic regression, conditioned on matching factors (i.e., sex and birthyear)*

*Model 1: no covariate*

*Model 2: adjusted for education and income*

*^a^Disposable income per consumption unit within family, rank in tertile by year among the included individuals*

*^b^Chronic pain included migraine, sexual pain, headache, temporomandibular joint disorders, gastro-esophageal reflux disease, irritable bowel disease (IBS), fibromyalgia, interstitial cystitis, chronic fatigue syndrome, back/neck pain, arthritis/osteoarthritis/joint pain, other musculoskeletal/connective tissue pain, and other painful conditions.*

#### **Table S9: Hazard ratios of mortality**

a. **All population – Suicide attempt^a^**

| **Outcome** | **Exposure group** | **Events** | **Total PY (100,000)** | **Incidence rate per 100,000 PYs (95% CI)** | **Model 1**  **HR (95% CI)** | **Model 2**  **HR (95% CI)** |
| --- | --- | --- | --- | --- | --- | --- |
| Subsequent attempt^c^ | No suicide attempt | 8823 | 73.27 | 120.41  (117.91, 122.95) | 1 (ref) | 1 (ref) |
|  | Suicide attempt | 34608 | 10.88 | 3181.98  (3148.55, 3215.69) | 26.38  (25.69, 27.08) | 23.39  (22.78, 24.02) |
| Suicide mortality | No suicide attempt | 1098 | 74.11 | 14.82  (13.95, 15.72) | 1 (ref) | 1 (ref) |
|  | Suicide attempt | 3835 | 14.49 | 264.63  (256.32, 273.14) | 18.22  (17.00, 19.52) | 16.40  (15.29, 17.60) |
| Non-suicide mortality | No suicide attempt | 3902 | 73.90 | 52.8  (51.16, 54.49) | 1 (ref) | 1 (ref) |
|  | Suicide attempt | 3989 | 14.58 | 273.56  (265.13, 282.18) | 5.27  (5.04, 5.51) | 4.41  (4.21, 4.63) |
| All-cause mortality | No suicide attempt | 5000 | 73.79 | 67.76  (65.89, 69.66) | 1 (ref) | 1 (ref) |
|  | Suicide attempt | 7824 | 14.17 | 552.17  (540.00, 564.54) | 8.43  (8.12, 8.75) | 7.31  (7.04, 7.59) |

b. **Stratified by sex – Suicide attempt^b^**

| **Sex group** | **Exposure group** | **Events** | **Total PY (100,000)** | **Incidence rate per 100,000 PYs (95% CI)** | **HR (95% CI)** | **p-value for interaction** |
| --- | --- | --- | --- | --- | --- | --- |
| **Subsequent suicide attempt^c^** | | | | | | |
| Male | No suicide attempt | 3191 | 28.80 | 110.81  (107.00, 114.72) | 1 (ref) | 2.1*10^-3^ |
|  | Suicide attempt | 13741 | 4.33 | 3171.75  (3118.94, 3225.23) | 24.71  (23.64, 25.83) |  |
| Female | No suicide attempt | 5632 | 44.48 | 126.63  (123.34, 129.98) | 1 (ref) |  |
|  | Suicide attempt | 20867 | 6.54 | 3188.76  (3145.64, 3232.32) | 22.66  (21.92, 23.43) |  |
| **Suicide mortality** | | | | | | |
| Male | No suicide attempt | 679 | 29.04 | 23.38  (21.65, 25.21) | 1 (ref) | 5.7*10^-2^ |
|  | Suicide attempt | 2260 | 5.58 | 405.29  (388.75, 422.35) | 15.51  (14.17, 16.97) |  |
| Female | No suicide attempt | 419 | 45.07 | 9.3  (8.43, 10.23) | 1 (ref) |  |
|  | Suicide attempt | 1575 | 8.92 | 176.65  (168.04, 185.6) | 17.82  (15.94, 19.91) |  |
| **Non-suicide mortality** | | | | | | |
| Male | No suicide attempt | 2097 | 28.94 | 72.45  (69.39, 75.62) | 1 (ref) | 1.9*10^-28^ |
|  | Suicide attempt | 2669 | 5.61 | 475.66  (457.78, 494.05) | 5.48  (5.16, 5.83) |  |
| Female | No suicide attempt | 1805 | 44.96 | 40.15  (38.32, 42.05) | 1 (ref) |  |
|  | Suicide attempt | 1320 | 8.97 | 147.14  (139.31, 155.3) | 3.21  (2.98, 3.45) |  |
| **All-cause mortality** | | | | | | |
| Male | No suicide attempt | 2776 | 28.87 | 96.15  (92.61, 99.79) | 1 (ref) | 2.0*10^-17^ |
|  | Suicide attempt | 4929 | 5.36 | 919.47  (893.98, 945.5) | 8.39  (7.98, 8.83) |  |
| Female | No suicide attempt | 2224 | 44.92 | 49.51  (47.48, 51.61) | 1 (ref) |  |
|  | Suicide attempt | 2895 | 8.81 | 328.64  (316.78, 340.84) | 6.04  (5.7, 6.4) |  |

**c. All population – Self-harm^a^**

| **Outcome** | **Exposure group** | **Events** | **Total PY (100,000)** | **Incidence rate per 100,000 PYs (95% CI)** | **Model 1**  **HR (95% CI)** | **Model 2**  **HR (95% CI)** |
| --- | --- | --- | --- | --- | --- | --- |
| Subsequent self-harm^c^ | No self-harm | 16870 | 94.36 | 178.79  (176.10, 181.51) | 1 (ref) | 1 (ref) |
|  | Self-harm | 47700 | 13.92 | 3425.84  (3395.17, 3456.73) | 19.13  (18.76, 19.5) | 17.59  (17.25, 17.94) |
| Suicide mortality | No self-harm | 1406 | 95.79 | 14.68  (13.92, 15.47) | 1 (ref) | 1 (ref) |
|  | Self-harm | 3923 | 18.85 | 208.17  (201.71, 214.79) | 14.39  (13.52, 15.31) | 12.95  (12.15, 13.79) |
| Non-suicide mortality | No self-harm | 5032 | 95.53 | 52.68  (51.23, 54.15) | 1 (ref) | 1 (ref) |
|  | Self-harm | 4428 | 18.91 | 234.11  (227.26, 241.11) | 4.53  (4.34, 4.72) | 3.79  (3.63, 3.95) |
| All-cause mortality | No self-harm | 6438 | 95.40 | 67.48  (65.84, 69.15) | 1 (ref) | 1 (ref) |
|  | Self-harm | 8351 | 18.50 | 451.46  (441.83, 461.25) | 6.91  (6.68, 7.14) | 5.97  (5.77, 6.18) |

*Results from stratified Cox proportional regression estimated the association between mortality/subsequent attempt outcomes and suicide attempt status, stratified by matching clusters. By stratifying for matching clusters, all models were adjusted for matching factor (i.e., sex and birthyear). PY=person-year | HR=Hazard ratio | CI=Confidence interval*

*^a^Model 1 was not adjusted for additional covariates. Model 2 was further adjusted for education and income.*

*^b^A model was fitted for each outcome, beside matching factors, adjusting for education, and income, with the inclusion of an interaction term between sex and the outcome. Estimates were presented separately for each sex.*

*^c^Comparing the risk of making the 2^nd^ suicide attempt/self-harm among those who had had an initial attempt/self-harm and the risk of making an initial attempt/self-harm among those had never had an attempt/self-harm*

#### **Table S10: Proportion of families with individual attempted suicide/inflicted self-harm**

|  | **Nuclear family^a^** | | **Inclusive family^b^** | |
| --- | --- | --- | --- | --- |
|  | **N** | **%** | **N** | **%** |
| N families | 2,554,768 |  | 2,044,041 |  |
| Family with suicide attempt | 250,476 | 9.80 | 204,754 | 10.02 |
| Family with self-harm | 327,812 | 12.83 | 271,669 | 13.29 |

*^a^Includes parents and full-sibling offspring. ^b^Includes all parents and siblings that are related to each other via at least 1 spouse. Details for family definitions in Methods S2.*

#### **Table S11:** **Familial aggregation and coaggregation**

| a. **Suicide attempt** | | | | | | | | | | | | | | |
| --- | --- | --- | --- | --- | --- | --- | --- | --- | --- | --- | --- | --- | --- | --- |
|  | **Cox regression** | | | | | | **Generalised Estimating Equations** | | | | | | | |
| **Relatedness** | **Coefficient** | **SE** | **p-value** | **HR** | **95% CI of HR** | | **Coefficient** | **SE** | **p-value** | | **OR** | | **95% CI of OR** | |
| **Aggregation** | | | | | | | | | | | | | | |
| Mother-offspring | 1.0542 | 0.0111 | <2.2*10^-308^ | 2.8697 | 2.8076 | 2.9330 | 1.2130 | 0.0133 | <2.2*10^-308^ | 3.3635 | | 3.2772 | | 3.4522 |
| Father-offspring | 0.9750 | 0.0115 | <2.2*10^-308^ | 2.6513 | 2.5923 | 2.7116 | 1.0384 | 0.0140 | <2.2*10^-308^ | 2.8246 | | 2.7482 | | 2.9032 |
| Full sibling | 1.0253 | 0.0115 | <2.2*10^-308^ | 2.7878 | 2.7256 | 2.8515 | 1.1749 | 0.0153 | <2.2*10^-308^ | 3.2379 | | 3.1420 | | 3.3367 |
| Maternal half-sibling | 0.5195 | 0.0199 | 1.3*10^-150^ | 1.6812 | 1.6170 | 1.7480 | 0.5890 | 0.0244 | 3.5*10^-129^ | 1.8022 | | 1.7182 | | 1.8903 |
| Paternal half-sibling | 0.3638 | 0.0205 | 2.2*10^-70^ | 1.4388 | 1.3821 | 1.4978 | 0.4557 | 0.0249 | 1.2*10^-74^ | 1.5773 | | 1.5021 | | 1.6563 |
| Coaggregation with **major depressive disorder** | | | | | | | | | | | | | | |
| Mother-offspring | 0.6530 | 0.0108 | <2.2*10^-308^ | 1.9213 | 1.8810 | 1.9626 | 0.8488 | 0.0090 | <2.2*10^-308^ | 2.3369 | | 2.2959 | | 2.3786 |
| Father-offspring | 0.5694 | 0.0130 | <2.2*10^-308^ | 1.7672 | 1.7228 | 1.8127 | 0.7189 | 0.0098 | <2.2*10^-308^ | 2.0522 | | 2.0131 | | 2.0920 |
| Full sibling | 0.5202 | 0.0119 | <2.2*10^-308^ | 1.6824 | 1.6438 | 1.7220 | 0.7864 | 0.0091 | <2.2*10^-308^ | 2.1955 | | 2.1568 | | 2.2349 |
| Maternal half-sibling | 0.2395 | 0.0203 | 5.3*10^-32^ | 1.2706 | 1.2209 | 1.3222 | 0.3696 | 0.0146 | 5.4*10^-141^ | 1.4472 | | 1.4063 | | 1.4893 |
| Paternal half-sibling | 0.1868 | 0.0208 | 3.1*10^-19^ | 1.2053 | 1.1571 | 1.2556 | 0.3100 | 0.0149 | 1.2*10^-95^ | 1.3634 | | 1.3241 | | 1.4039 |
| Coaggregation with **schizophrenia** | | | | | | | | | | | | | | |
| Mother-offspring | 0.6323 | 0.0429 | 2.9*10^-49^ | 1.8819 | 1.7303 | 2.0468 | 0.9497 | 0.0343 | 4.8*10^-169^ | 2.5850 | | 2.4170 | | 2.7646 |
| Father-offspring | 0.7589 | 0.0499 | 2.7*10^-52^ | 2.1360 | 1.9371 | 2.3554 | 0.7275 | 0.0377 | 3.7*10^-83^ | 2.0699 | | 1.9226 | | 2.2284 |
| Full sibling | 0.5550 | 0.0396 | 1.1*10^-44^ | 1.7420 | 1.6119 | 1.8825 | 0.8249 | 0.0319 | 2.8*10^-147^ | 2.2816 | | 2.1432 | | 2.4289 |
| Maternal half-sibling | 0.3103 | 0.0691 | 7.0*10^-6^ | 1.3638 | 1.1911 | 1.5615 | 0.4498 | 0.0568 | 2.4*10^-15^ | 1.5679 | | 1.4028 | | 1.7525 |
| Paternal half-sibling | 0.1918 | 0.0702 | 6.3*10^-3^ | 1.2114 | 1.0556 | 1.3902 | 0.3282 | 0.0605 | 6.0*10^-8^ | 1.3884 | | 1.2330 | | 1.5633 |
| Coaggregation with **bipolar disorder** | | | | | | | | | | | | | | |
| Mother-offspring | 0.6124 | 0.0237 | 6.2*10^-147^ | 1.8449 | 1.7611 | 1.9327 | 0.9410 | 0.0179 | <2.2*10^-308^ | 2.5625 | | 2.4742 | | 2.6540 |
| Father-offspring | 0.5280 | 0.0300 | 1.7*10^-69^ | 1.6955 | 1.5988 | 1.7980 | 0.8015 | 0.0199 | <2.2*10^-308^ | 2.2289 | | 2.1438 | | 2.3174 |
| Full sibling | 0.5013 | 0.0278 | 1.3*10^-72^ | 1.6509 | 1.5633 | 1.7434 | 0.8620 | 0.0181 | <2.2*10^-308^ | 2.3680 | | 2.2854 | | 2.4535 |
| Maternal half-sibling | 0.2076 | 0.0490 | 2.3*10^-5^ | 1.2307 | 1.1180 | 1.3548 | 0.4039 | 0.0303 | 1.8*10^-40^ | 1.4977 | | 1.4113 | | 1.5894 |
| Paternal half-sibling | 0.2119 | 0.0479 | 9.8*10^-6^ | 1.2360 | 1.1252 | 1.3578 | 0.3397 | 0.0303 | 4.4*10^-29^ | 1.4045 | | 1.3234 | | 1.4906 |
| Coaggregation with **substance use disorder** | | | | | | | | | | | | | | |
| Mother-offspring | 0.9317 | 0.0125 | <2.2*10^-308^ | 2.5388 | 2.4773 | 2.6019 | 1.0690 | 0.0100 | <2.2*10^-308^ | 2.9125 | | 2.8558 | | 2.9703 |
| Father-offspring | 0.7982 | 0.0096 | <2.2*10^-308^ | 2.2215 | 2.1799 | 2.2638 | 0.9573 | 0.0102 | <2.2*10^-308^ | 2.6046 | | 2.5530 | | 2.6573 |
| Full sibling | 0.7840 | 0.0119 | <2.2*10^-308^ | 2.1902 | 2.1396 | 2.2420 | 1.0149 | 0.0103 | <2.2*10^-308^ | 2.7591 | | 2.7040 | | 2.8154 |
| Maternal half-sibling | 0.4036 | 0.0187 | 3.1*10^-103^ | 1.4971 | 1.4432 | 1.5530 | 0.5139 | 0.0157 | 8.2*10^-234^ | 1.6719 | | 1.6211 | | 1.7242 |
| Paternal half-sibling | 0.2847 | 0.0194 | 9.8*10^-49^ | 1.3293 | 1.2797 | 1.3809 | 0.3996 | 0.0164 | 2.2*10^-131^ | 1.4912 | | 1.4441 | | 1.5398 |

| b. **Self-harm** | | | | | | | | | | | | | | | |
| --- | --- | --- | --- | --- | --- | --- | --- | --- | --- | --- | --- | --- | --- | --- | --- |
|  | **Cox regression** | | | | | | **Generalised Estimating Equations** | | | | | | | | |
| **Relatedness** | **Coefficient** | **SE** | **p-value** | **HR** | **95% CI of HR** | | **Coefficient** | **SE** | **p-value** | | **OR** | | **95% CI of OR** | | |
| **Aggregation** |  |  |  |  |  |  |  |  |  |  | |  | |  | |
| Mother-offspring | 1.0542 | 0.0111 | <2.2*10^-308^ | 2.8697 | 2.8076 | 2.9330 | 1.1035 | 0.0111 | <2.2*10^-308^ | 3.0147 | | 2.9500 | | 3.0808 | |
| Father-offspring | 0.9750 | 0.0115 | <2.2*10^-308^ | 2.6513 | 2.5923 | 2.7116 | 1.0134 | 0.0112 | <2.2*10^-308^ | 2.7549 | | 2.6949 | | 2.8162 | |
| Full sibling | 1.0253 | 0.0115 | <2.2*10^-308^ | 2.7878 | 2.7256 | 2.8515 | 1.0957 | 0.0119 | <2.2*10^-308^ | 2.9914 | | 2.9223 | | 3.0622 | |
| Maternal half-sibling | 0.5195 | 0.0199 | 1.3*10^-150^ | 1.6812 | 1.6170 | 1.7480 | 0.5675 | 0.0205 | 2.8*10^-168^ | 1.7639 | | 1.6943 | | 1.8363 | |
| Paternal half-sibling | 0.3638 | 0.0205 | 2.2*10^-70^ | 1.4388 | 1.3821 | 1.4978 | 0.4148 | 0.0209 | 6.1*10^-88^ | 1.5141 | | 1.4534 | | 1.5773 | |
| Coaggregation with **major depressive disorder** | | | | | | | | | | | | | | | |
| Mother-offspring | 0.6530 | 0.0108 | <2.2*10^-308^ | 1.9213 | 1.8810 | 1.9626 | 0.7172 | 0.0084 | <2.2*10^-308^ | | 2.0486 | | 2.0153 | | 2.0825 |
| Father-offspring | 0.5694 | 0.0130 | <2.2*10^-308^ | 1.7672 | 1.7228 | 1.8127 | 0.5853 | 0.0090 | <2.2*10^-308^ | | 1.7956 | | 1.7640 | | 1.8277 |
| Full sibling | 0.5202 | 0.0119 | <2.2*10^-308^ | 1.6824 | 1.6438 | 1.7220 | 0.6250 | 0.0081 | <2.2*10^-308^ | | 1.8682 | | 1.8387 | | 1.8982 |
| Maternal half-sibling | 0.2395 | 0.0203 | 5.3*10^-32^ | 1.2706 | 1.2209 | 1.3222 | 0.3119 | 0.0135 | 1.4*10^-118^ | | 1.3660 | | 1.3304 | | 1.4026 |
| Paternal half-sibling | 0.1868 | 0.0208 | 3.1*10^-19^ | 1.2053 | 1.1571 | 1.2556 | 0.2558 | 0.0136 | 2.1*10^-78^ | | 1.2915 | | 1.2574 | | 1.3265 |
| Coaggregation with **schizophrenia** | | | | | | | | | | | | | | | |
| Mother-offspring | 0.6323 | 0.0429 | 2.9*10^-49^ | 1.8819 | 1.7303 | 2.0468 | 0.8002 | 0.0326 | 2.3*10^-133^ | | 2.2260 | | 2.0884 | | 2.3727 |
| Father-offspring | 0.7589 | 0.0499 | 2.7*10^-52^ | 2.1360 | 1.9371 | 2.3554 | 0.6288 | 0.0353 | 7,00*10^-71^ | | 1.8754 | | 1.7499 | | 2.0098 |
| Full sibling | 0.5550 | 0.0396 | 1.1*10^-44^ | 1.7420 | 1.6119 | 1.8825 | 0.6530 | 0.0299 | 1.1*10^-105^ | | 1.9214 | | 1.8120 | | 2.0374 |
| Maternal half-sibling | 0.3103 | 0.0691 | 7.0*10^-6^ | 1.3638 | 1.1911 | 1.5615 | 0.3603 | 0.0540 | 2.5*10^-11^ | | 1.4337 | | 1.2898 | | 1.5938 |
| Paternal half-sibling | 0.1918 | 0.0702 | 6.3*10^-3^ | 1.2114 | 1.0556 | 1.3902 | 0.2995 | 0.0555 | 6.7*10^-8^ | | 1.3492 | | 1.2102 | | 1.5042 |
| Coaggregation with **bipolar disorder** | | | | | | | | | | | | | | | |
| Mother-offspring | 0.6124 | 0.0237 | 6.2*10^-147^ | 1.8449 | 1.7611 | 1.9327 | 0.8174 | 0.0167 | <2.2*10^-308^ | | 2.2647 | | 2.1917 | | 2.3402 |
| Father-offspring | 0.5280 | 0.0300 | 1.7*10^-69^ | 1.6955 | 1.5988 | 1.7980 | 0.6832 | 0.0185 | 7.3*10^-299^ | | 1.9802 | | 1.9097 | | 2.0533 |
| Full sibling | 0.5013 | 0.0278 | 1.3*10^-72^ | 1.6509 | 1.5633 | 1.7434 | 0.7098 | 0.0165 | <2.2*10^-308^ | | 2.0336 | | 1.9689 | | 2.1005 |
| Maternal half-sibling | 0.2076 | 0.0490 | 2.3*10^-5^ | 1.2307 | 1.1180 | 1.3548 | 0.3504 | 0.0283 | 3.8*10^-35^ | | 1.4196 | | 1.3429 | | 1.5006 |
| Paternal half-sibling | 0.2119 | 0.0479 | 9.8*10^-6^ | 1.2360 | 1.1252 | 1.3578 | 0.2866 | 0.0279 | 9.1*10^-25^ | | 1.3319 | | 1.2611 | | 1.4068 |
| Coaggregation with **substance use disorder** | | | | | | | | | | | | | | | |
| Mother-offspring | 0.9317 | 0.0125 | <2.2*10^-308^ | 2.5388 | 2.4773 | 2.6019 | 0.9215 | 0.0094 | <2.2*10^-308^ | | 2.5131 | | 2.4671 | | 2.5600 |
| Father-offspring | 0.7982 | 0.0096 | <2.2*10^-308^ | 2.2215 | 2.1799 | 2.2638 | 0.8002 | 0.0095 | <2.2*10^-308^ | | 2.2261 | | 2.1850 | | 2.2679 |
| Full sibling | 0.7840 | 0.0119 | <2.2*10^-308^ | 2.1902 | 2.1396 | 2.2420 | 0.8336 | 0.0093 | <2.2*10^-308^ | | 2.3015 | | 2.2599 | | 2.3439 |
| Maternal half-sibling | 0.4036 | 0.0187 | 3.1*10^-103^ | 1.4971 | 1.4432 | 1.5530 | 0.4552 | 0.0144 | 2.9*10^-219^ | | 1.5764 | | 1.5325 | | 1.6215 |
| Paternal half-sibling | 0.2847 | 0.0194 | 9.8*10^-49^ | 1.3293 | 1.2797 | 1.3809 | 0.3469 | 0.0148 | 3.2*10^-121^ | | 1.4147 | | 1.3742 | | 1.4564 |

#### **Table S12: Heritability and genetic correlations**

| a. **Suicide attempt** | | | | | | | | | | | |
| --- | --- | --- | --- | --- | --- | --- | --- | --- | --- | --- | --- |
|  | **ACE** | | | | **AE** | | | | | |  |
| **Component** | **Estimate** | **SE** | **95% CI** |  | **Estimate** | **SE** | **95% CI** | | |  | **Model fit^a^** |
| A | **0.4189** | **0.0316** | **0.3592** | **0.4821** | 0.4953 | 0.0072 | 0.4820 | | | 0.5091 | ACE 𝝌^2^=521.9188  AE 𝝌^2^=529.6738  p-value=5.36*10^-3^ |
| C | **0.0357** | **0.0144** | **0.0082** | **0.0636** |  |  |  | | |  |  |
| E | **0.5453** | **0.0179** | **0.5106** | **0.5800** | 0.5047 | 0.0072 | 0.4909 | | | 0.5180 |  |
| ***r_g_* with** | ***r_g_*** | **SE** | **95% CI** |  | ***r_g_*** | **SE** | **95% CI** | | |  | **Model fit^b^** |
| Major depressive disorder | **0.7700** | **0.0333** | **0.7346** | **0.8630** | 0.7749 | 0.0087 | 0.7573 | | | 0.7924 | ACE 𝝌^2^=1301.6702  AE 𝝌^2^=1313.4154  p-value=3.81*10^-3^ |
| Schizophrenia | 0.5214 | 0.1702 | 0.2679 | 0.7615 | **0.4816** | **0.0186** | **0.4441** | | | **0.5182** | ACE 𝝌^2^=1438.8458  AE 𝝌^2^=1448.5235  p-value=2.15*10^-2^ |
| Bipolar disorder | 0.6982 | 0.0665 | 0.5778 | 0.8399 | **0.6180** | **0.0137** | **0.5878** | | | **0.6415** | ACE 𝝌^2^=661.2082  AE 𝝌^2^=670.496  p-value=2.57*10^-2^ |
| Substance use disorder | **0.8517** | **0.1181** | **0.8323** | **0.9568** | 0.8790 | 0.0073 | 0.8675 | | | 0.8954 | ACE 𝝌^2^=1344.4094  AE 𝝌^2^=1382.0187  p-value=3.42*10^-8^ |
| b. **Self-harm** | | | | | | | | | | | |
|  | **ACE** | | | | **AE** | | | | | |  |
| **Component** | **Estimate** | **SE** | **95% CI** | | **Estimate** | **SE** | | **95% CI** | | | **Model fit^a^** |
| A | **0.4226** | **0.0288** | **0.3707** | **0.4780** | 0.5043 | 0.0064 | | 0.4927 | 0.5170 | | ACE 𝝌^2^=498.3996  AE 𝝌^2^=510.1691  p-value=6.02*10^-4^ |
| C | **0.0385** | **0.0132** | **0.0119** | **0.0631** |  |  | |  |  | |  |
| E | **0.5388** | **0.0162** | **0.5067** | **0.5680** | 0.4957 | 0.0064 | | 0.4830 | 0.5073 | |  |
| ***r_g_* with** | ***r_g_*** | **SE** | **95% CI** | | ***r_g_*** | **SE** | | **95% CI** | | | **Model fit^b^** |
| Major depressive disorder | **0.6239** | **0.0309** | **0.5831** | **0.7022** | 0.6294 | 0.0097 | | 0.6141 | 0.6530 | | ACE 𝝌^2^=980.2867  AE 𝝌^2^=993.5244  p-value=4.15*10^-3^ |
| Schizophrenia | **0.3303** | **0.0895** | **0.1966** | **0.4971** | 0.3821 | 0.0189 | | 0.3391 | 0.4145 | | ACE 𝝌^2^=1474.9514  AE 𝝌^2^=1488.0219  p-value=4.49*10^-3^ |
| Bipolar disorder | **0.5304** | **0.0578** | **0.4363** | **0.6636** | 0.5057 | 0.0107 | | 0.4875 | 0.5296 | | ACE 𝝌^2^=641.904  AE 𝝌^2^=654.6228  p-value=5.29*10^-3^ |
| Substance use disorder | **0.7276** | **0.0368** | **0.6861** | **0.8254** | 0.7641 | 0.0093 | | 0.7465 | 0.7830 | | ACE 𝝌^2^=1618.8099  AE 𝝌^2^=1647.3276  p-value=2.83*10^-6^ |

**SE and 95% CI estimated from Bootstrap resampling (among successful optimizations, i.e., Mx exit status code 0). ^a^difference in degree of freedom=1. ^b^difference in degree of freedom=3. p-value for testing whether the fit of the AE model is equal to that of the ACE model. p<0.05 rejects the equal fit and indicates AE model fit is worse than ACE. Results from the best-fit model are in bold text and reported as main results***.**

#### **Table S13: Psychiatric contact before and after the initial suicide attempt**

|  | **1 month before^a^** | **1 month after^b^** | **Post-pre change (%)** |
| --- | --- | --- | --- |
| **Any psychiatric contact** | 721 (19.8%) | 1405 (38.7%) | 18.9 |
| **Any non-psychiatric contact** | 1275 (35.0%) | 1637 (45.1%) | 10.1 |

*Data from 3646 individuals with initial suicide attempt recorded in Stockholm region during 1/10/2005-30/9/2013. Groups are not mutually exclusive. Non-psychiatry contacts include missing type of clinic visit (~5%). ^a^Percentage was calculated among 3646 individuals. ^b^Percentage was calculated among 3629 individuals who survived the initial attempt.*

#### **Table S14: Number of individuals on psychotropic medication 1 month before and after the initial suicide attempt**

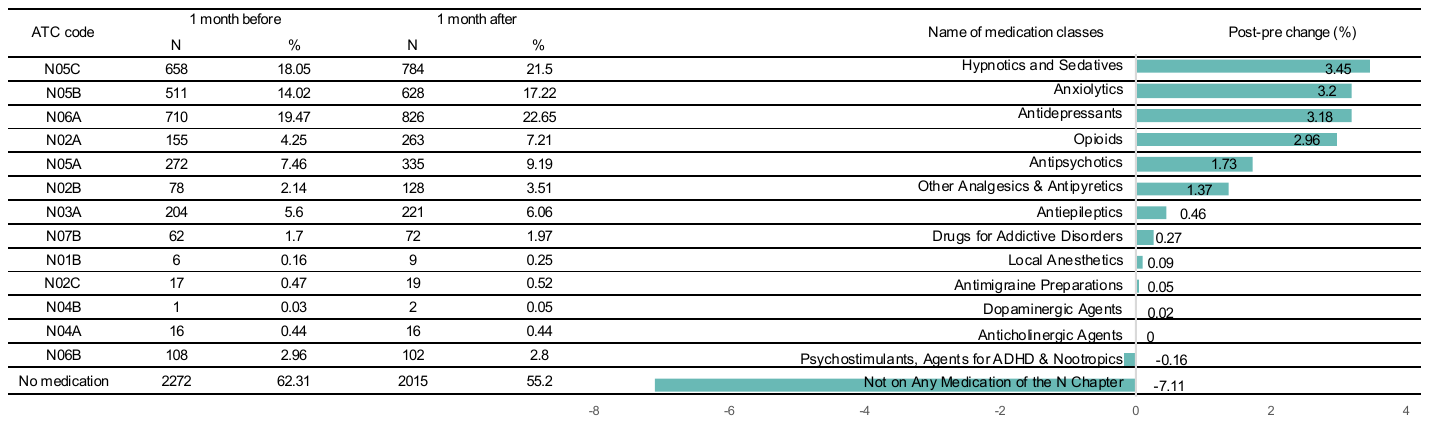

*N=number of people. Proportion counted among 3646 individuals with initial suicide attempt recorded in Stockholm region during 1/10/2005-30/9/2013. Medications dispensed on the day of initial suicide attempt were counted as 1 month after*

### **SUPPLEMENTARY FIGURES**

**a. Suicide attempt**

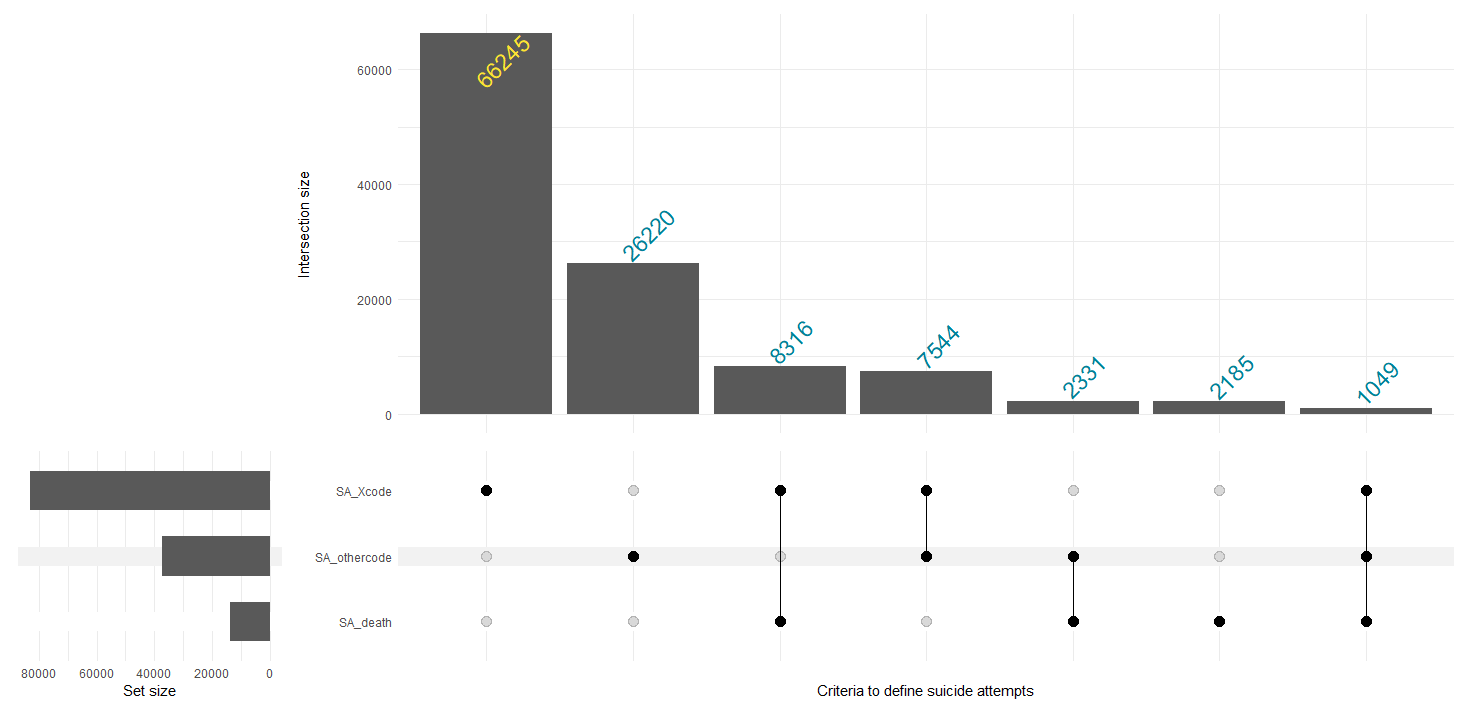

**b. Self-harm**

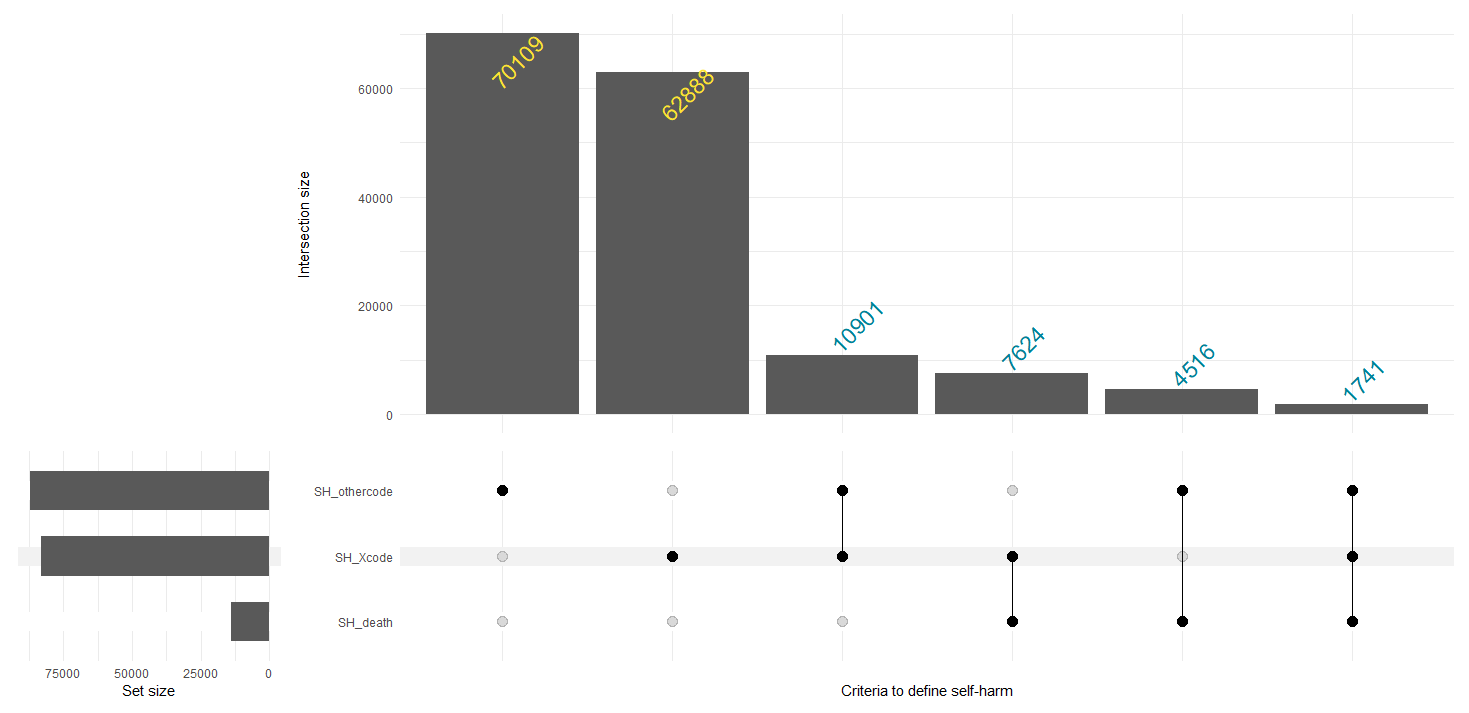

#### **Figure S1: Number of people meeting each criterion for definition of suicide attempt/self-harm**

a. ***Suicide attempt***

*SA_Xcode consists of individuals with at least 1 record with diagnosis ICD-10 codes X60-X84*

*SA_othercode consists of individuals with at least 1 record with diagnosis codes Y10-Y34, Y87.0, Y87.2, E980-E989, E950-E95* ***AND*** *meeting one of the two additional conditions: a. Used a notably lethal method of self-harm (firearm, jumping from heights, motor vehicle crash, suffocation, or poisoning by domestic gas)* ***OR*** *b. Led to inpatient care*

*SA_death consists of individuals with any record of death by suicide (X, Y, E codes) from Cause of Death Register*

b. ***Self-harm***

*SH_Xcode consists of individuals with at least 1 record with diagnosis ICD-10 codes X60-X84*

*SH_othercode consists of individuals with at least 1 record with diagnosis codes Y10-Y34, Y87.0, Y87.2, E980-E989, E950-E95*

*SH_death consists of individuals with any record of death by suicide (X, Y, E codes) from Cause of Death register*

Never attempted suicide at initial attempt of matched case (N=519,285)

match 1:5 on sex, birth year

Individuals born in Sweden

during 1963-1998

(N=3,859,963)

Excluded (N=163,343)

Death before age 10 (N=38,903)

First emigration before age 10 (N=124,440)

Broad birth cohort included

(N=3,696,620)

Final sample size

Analysis

Healthcare & medication utilization

Individuals with initial suicide attempt recorded in Stockholm region during 1/8/2005-30/11/2013

(N=3646)

Individuals w. Suicide attempt/Self-harm

(N=113,890 Suicide attempt)

(N=157,779 Self-harm)

Never attempted suicide/self-harm at initial attempt of matched case

(N=569,450 Suicide attempt)

(N=788,895 Self-harm)

Outcomes

(Matched cohort)

Risk factors

(Nested case-control)

Incidence of suicide attempt

match 1:5 on sex, birth year

Excluded (N=58,615)

1. Adopted children (N=29,271)

2. Missing parent ID (N=29,344)

Included probands (N=3,638,005)

Genetic epidemiology*

Parent-offspring (4,024,733 pairs)

Full-siblings (3,338,793 pairs)

Maternal half-siblings (592,797 pairs)

Paternal half-siblings (839,074 pairs)

Identified Sweden-born relatives

Individuals survived initial suicide attempt

(N=103,857)

*Footnote next page*

#### **Figure S2: Sample size and overview of analyses**

**For genetic epidemiology analyses*

*After removing adopted children and those without information of both parents, we included 3,638,005 probands. We used Multi-Generation Register data to identify relatives who were born in Sweden. Relatives could be born outside the identified birth cohort period (1963-1998). We removed pairs where the relative died or had first emigration record before 10 years old; or did not reach age 10 by 31/12/2019.*

*The number of relative pairs is presented in the figure.*

***For the aggregation/coaggregation analyses****, we constructed a dataset separately for each type of relative where each pair appeared twice if both proband and relative were born within 1963-1998, with the role of proband and relative switched between the two individuals; and appeared once if the relative was born outside the 1963-1998 period. This resulted in:*

- ***3,653,013 mother-offspring pairs***
- ***3,477,550 father-offspring pairs***
- ***4,992,249 full sibling pairs***
- ***908,740 maternal half-sibling pairs***
- ***1,164,125 paternal half-sibling pairs***

***For the estimation of heritability and genetic correlation****s using SEM, to ensure the quality of phenotype for both siblings, we included pairs where both siblings were born within the cohort 1963-1998. The final analyses included* ***2,143,654*** ***full sibling pairs*** *and* ***343,075 maternal half-sibling pairs****.*

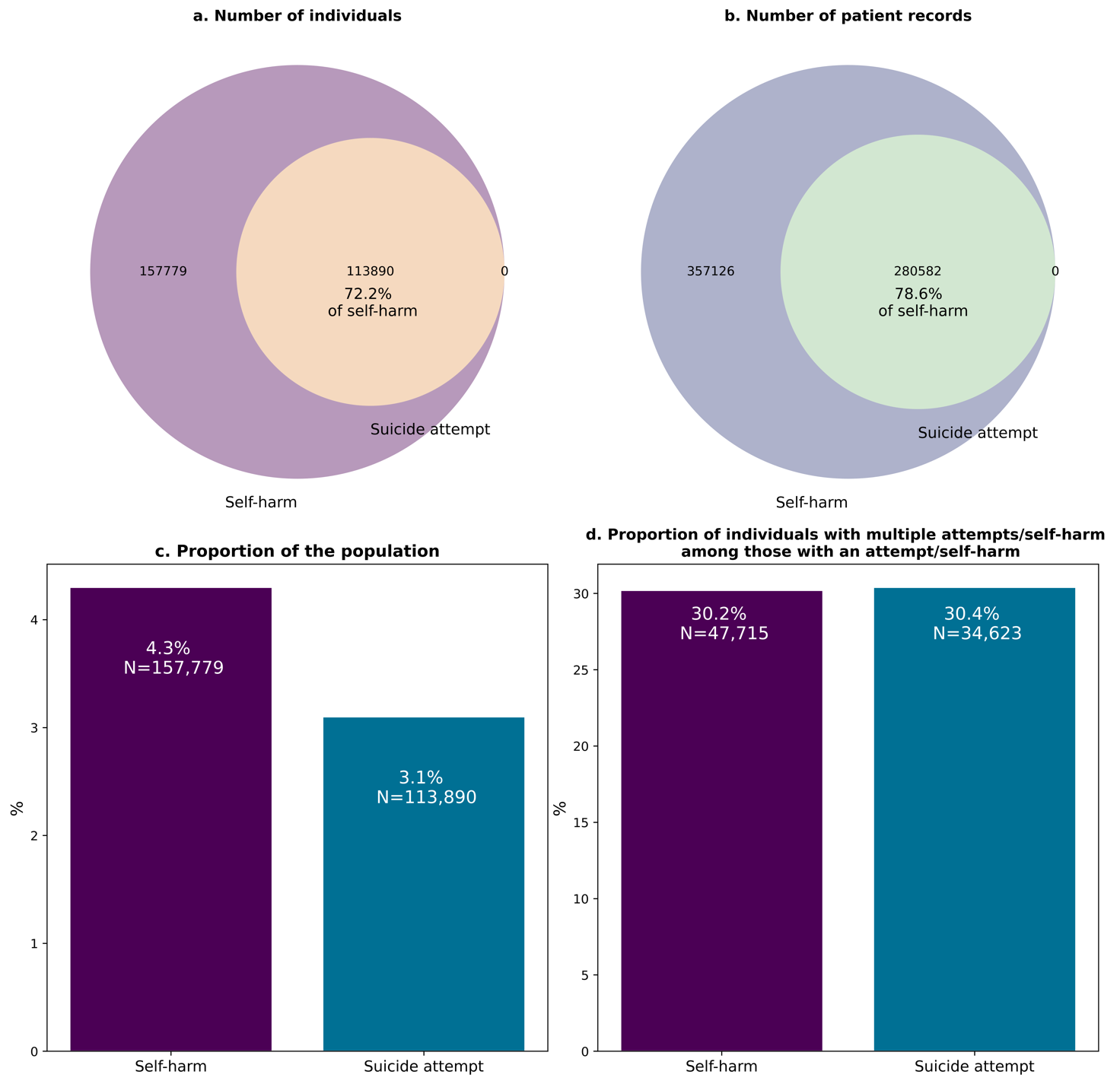

#### **Figure S3: Number of individuals and records of suicide attempt and self-harm**

***a.*** *Proportion of individuals with suicide attempts among those with any self-harm*

***b.*** *Proportion of suicide attempt patient records among all patient records of any self-harm*

***c.*** *Proportion of individuals with suicide attempts and self-harm among the included population*

***d.*** *Proportion of individuals with multiple attempts (>1) among all individuals with a suicide attempt/self-harm*

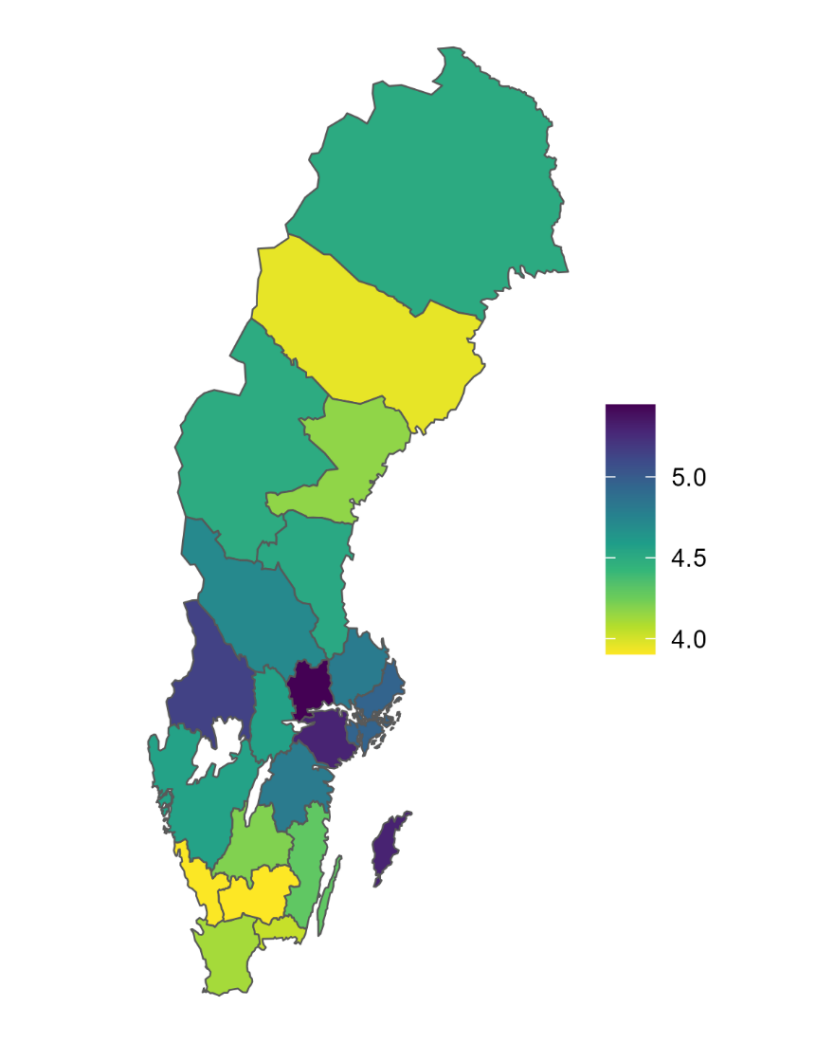

#### **Figure S4: Cumulative incidence of suicide attempts by county of birth**

*Colors represent lifetime risk (%) (i.e., cumulative incidence at the maximum age of 57)*

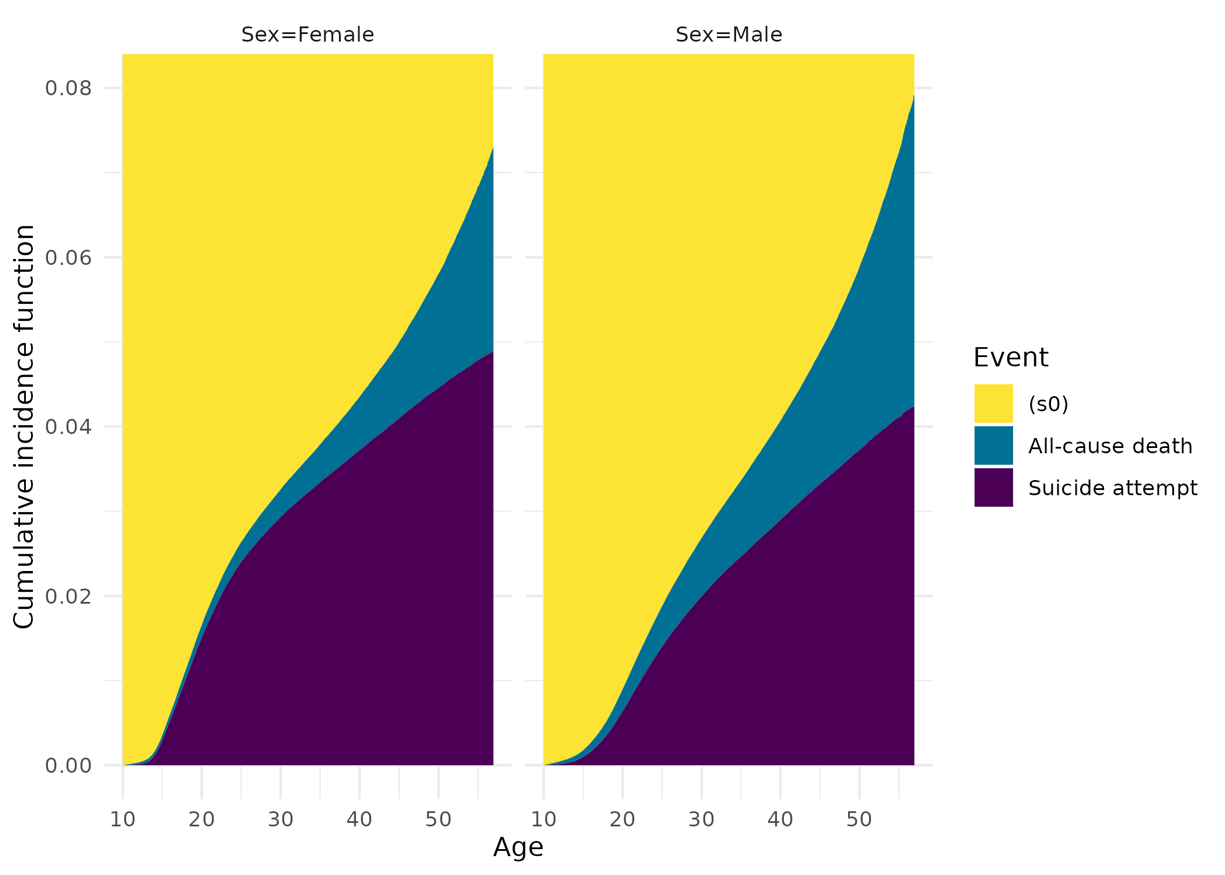

#### **Figure S5: Cumulative incidence of initial suicide attempt with all-cause mortality as competing risk**

*Estimated using Aalen-Johansen estimator*

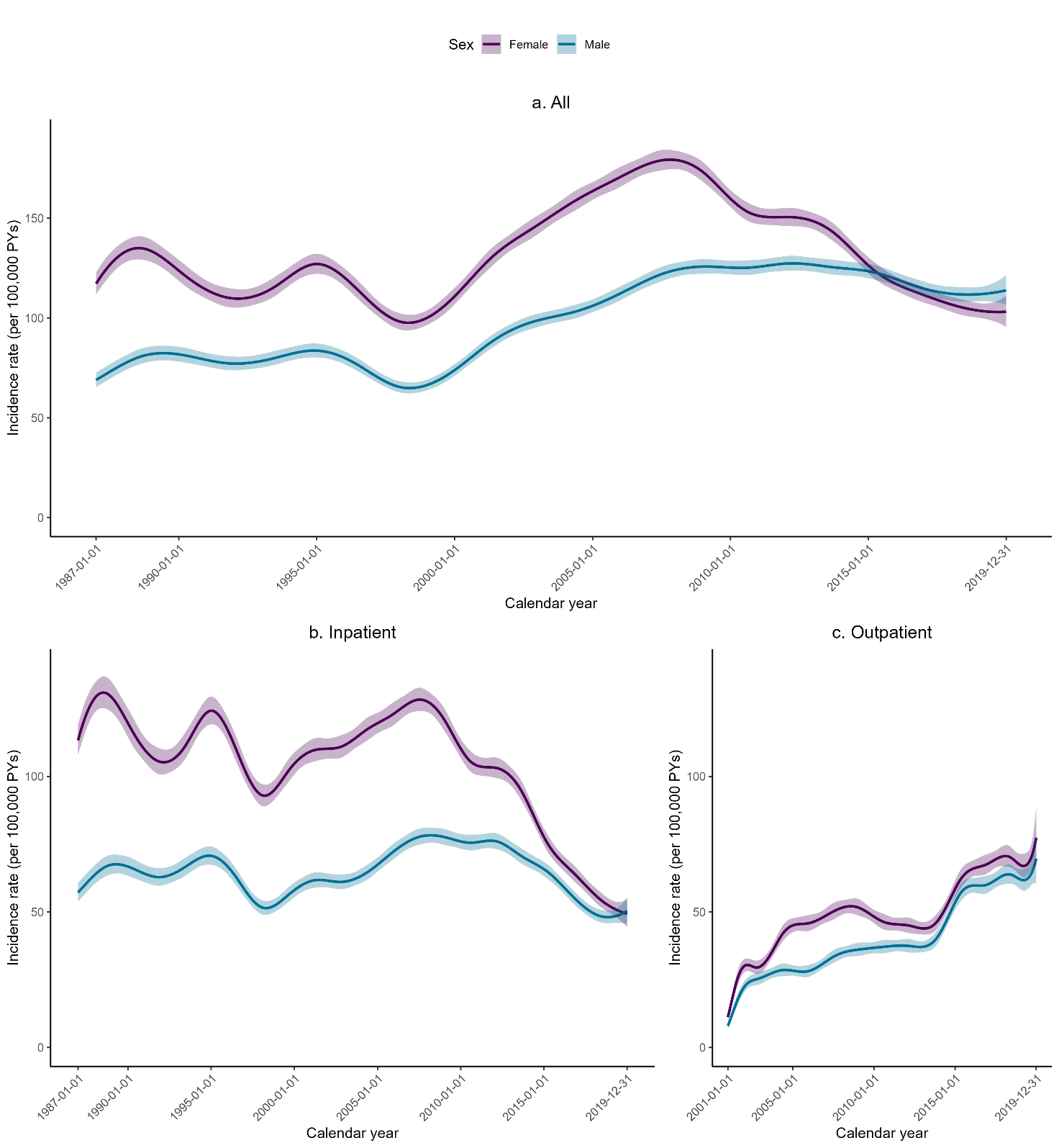

#### **Figure S6: Incidence rate of suicide attempt over calendar years**

*Colors represent data separately by sex. Shading regions show 95% confidence intervals of the estimates.*

***a)*** ***Incidence rate by calendar year at attempt****.* *Because the inpatient care register reached 100% coverage in 1987, data were presented from 1987 onward*

***b) Incidence rate by calendar year at attempt captured in inpatient care***

***c) Incidence rate by calendar year at attempt captured in outpatient care***

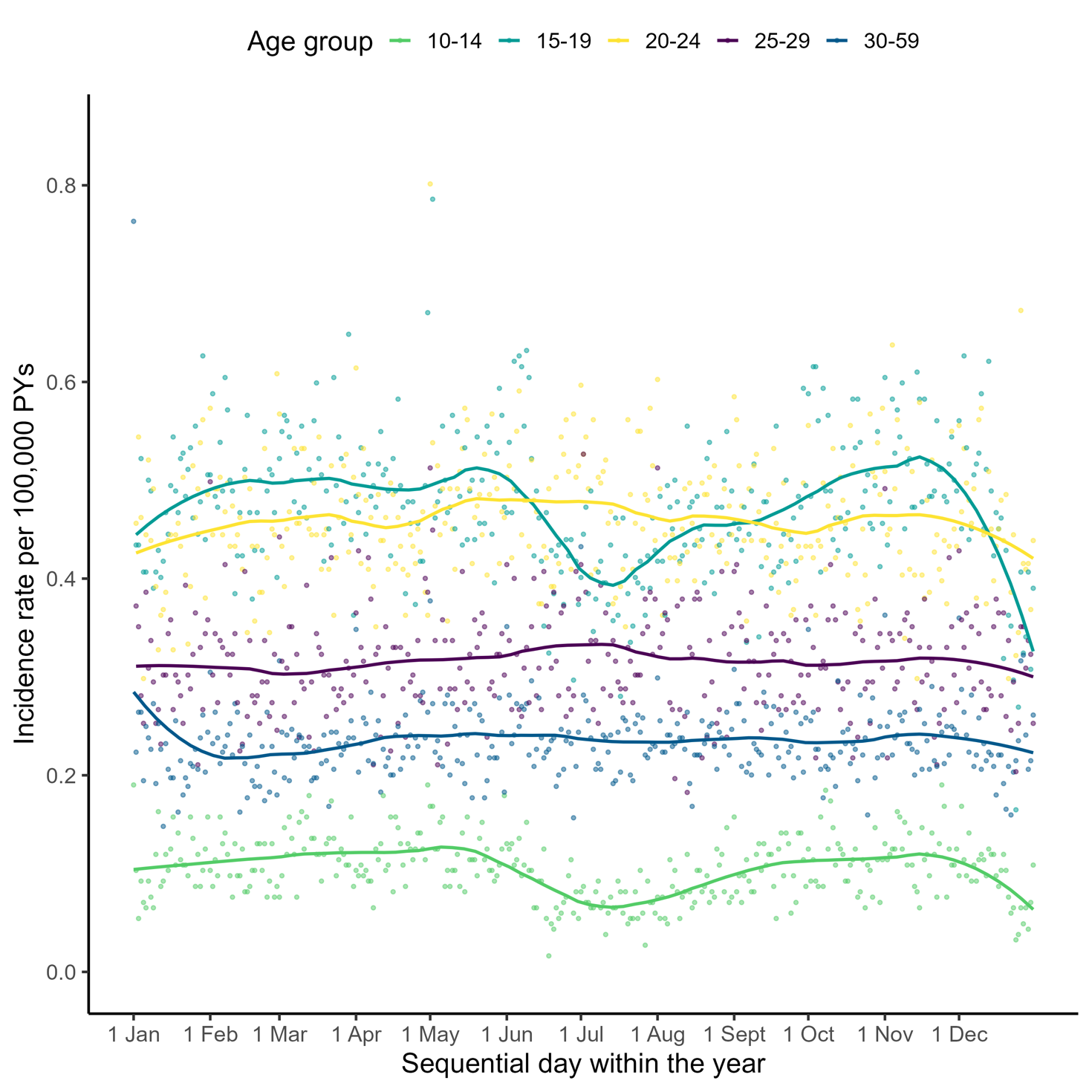

#### **Figure S7: Incidence rate of suicide attempt on each day of the year, by age group**

*The dots show number of events per 100,000 person-year on each sequential day within the year. The age categories were organized into 5-year intervals, with the exception of the 30-59 age group, which was collapsed into one group.* *Smooth splines are represented using penalized regression splines in generalized additive model.*

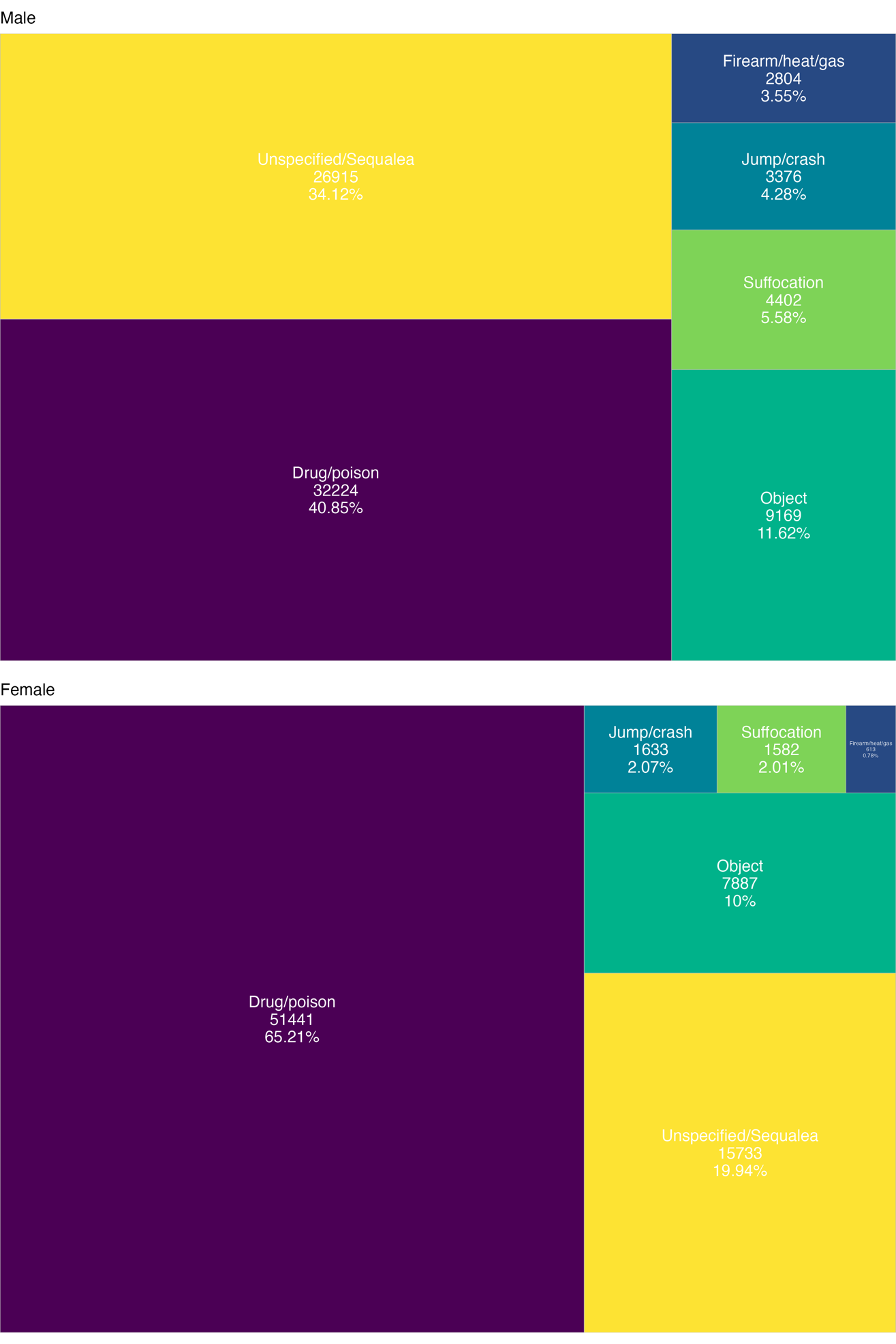

#### **Figure S8:** Methods used in the **initial self-harm** including suicide attempt

*Data from the initial patient record of each individual for males (top) and females (bottom).* *Methods that accounted for <1% were collapsed to a single category of Firearm/heat/gas. The areas are proportional to the number of individuals within each sex.*

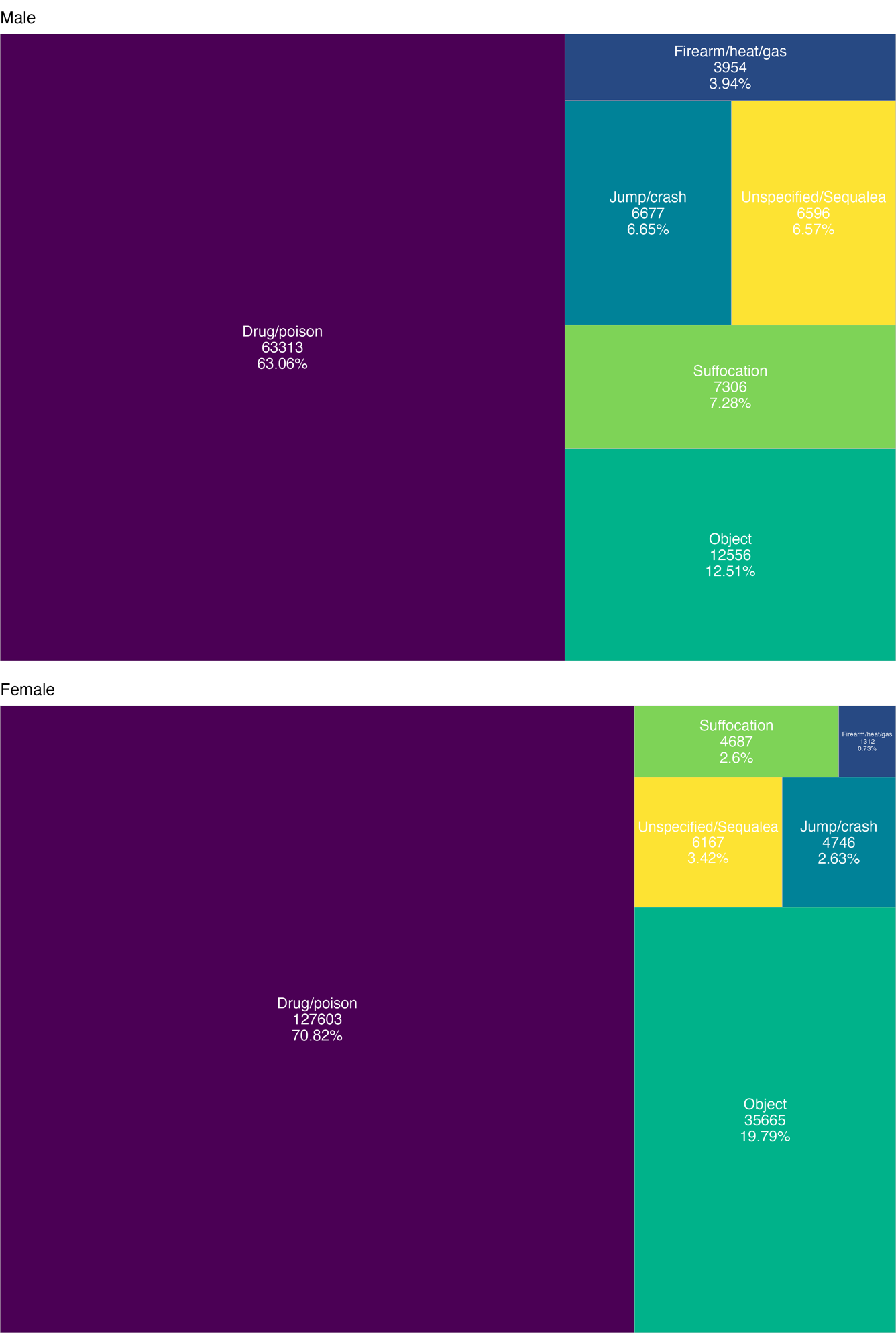

#### **Figure S9:** Methods used in **suicide attempts** from **all records**

*Data from all patient records of each individual for males (top) and females (bottom).* *Methods that accounted for <1% were collapsed to the same category of Firearm/heat/gas. The areas are proportional to the number of records within each sex.*

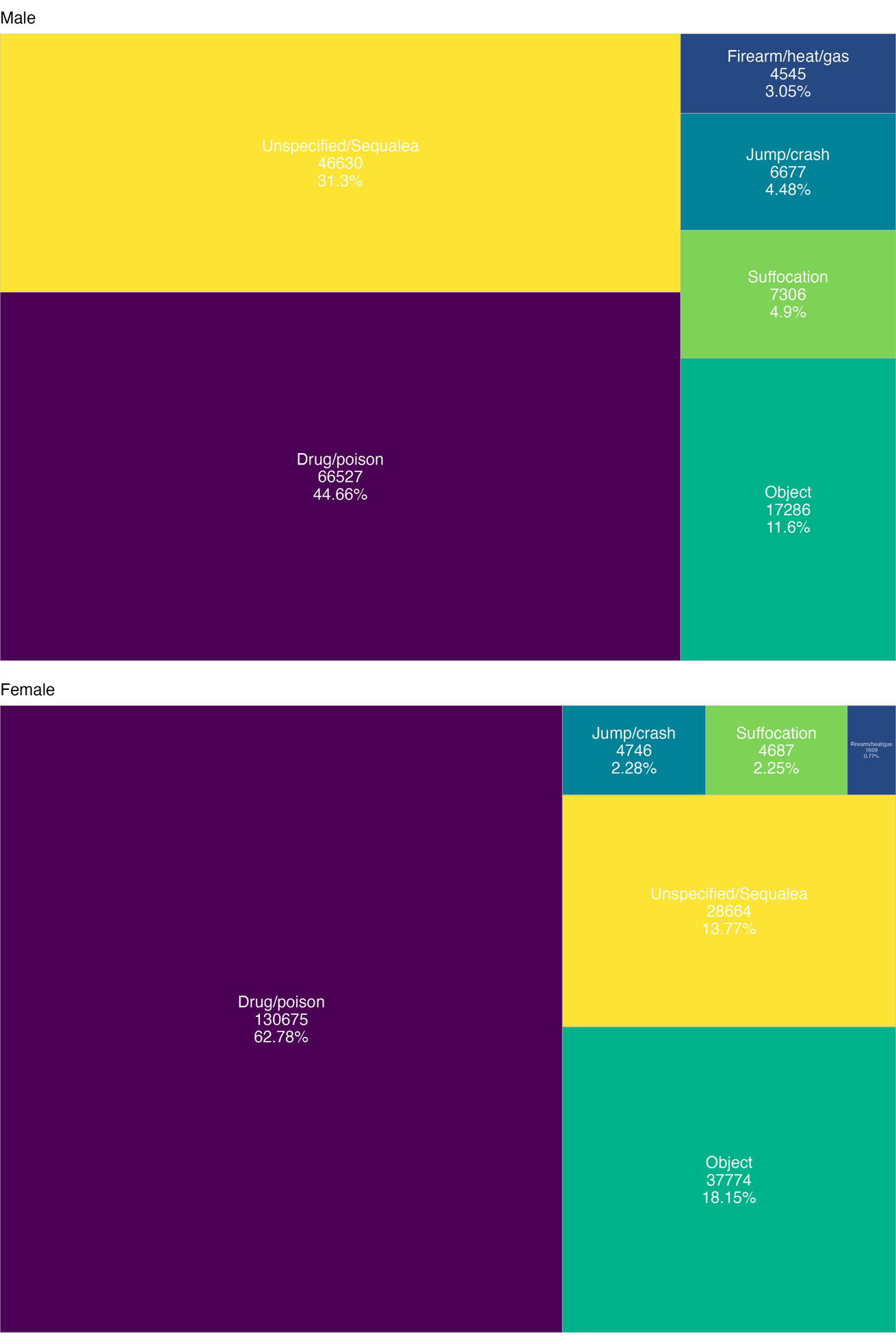

#### **Figure S10:** Methods used in **self-harm** from **all records**

*Data from all patient records of each individual for males (top) and females (bottom).* *Methods that accounted for <1% were collapsed to the same category of Firearm/heat/gas. The areas are proportional to the number of records within each sex.*

#### **Figure S11**: Distribution of risk factors among those with and without an suicide eattempt

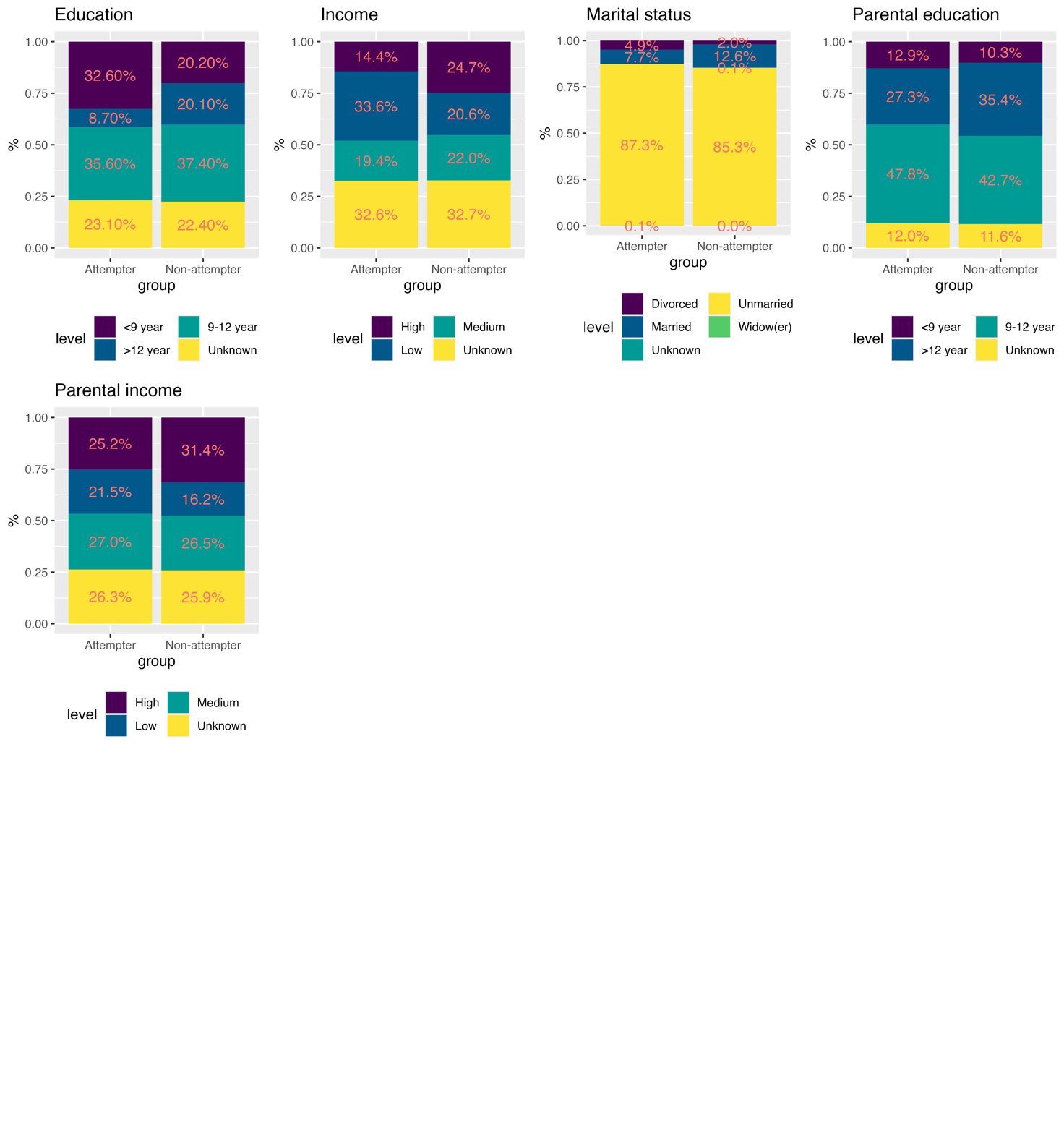

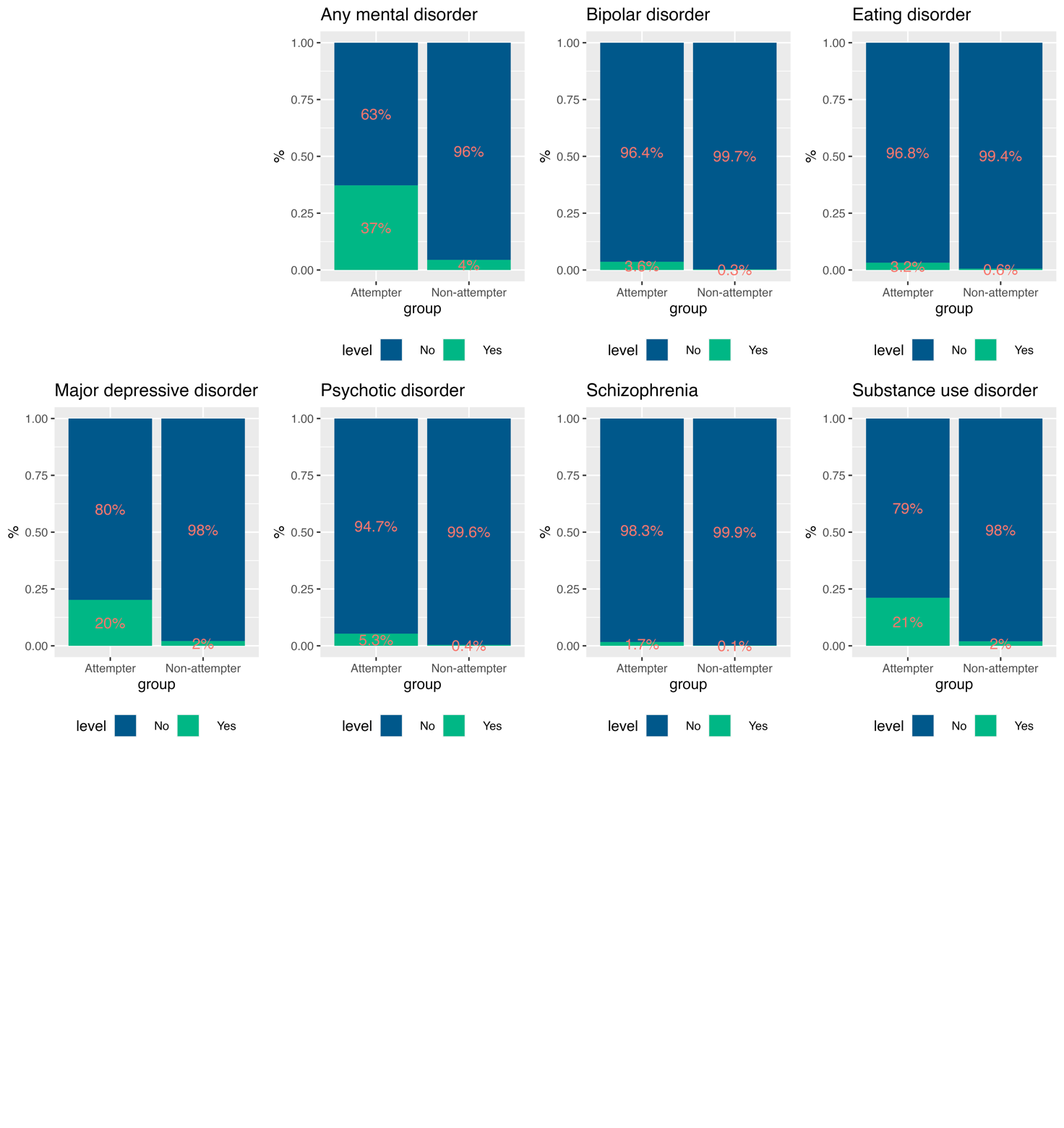

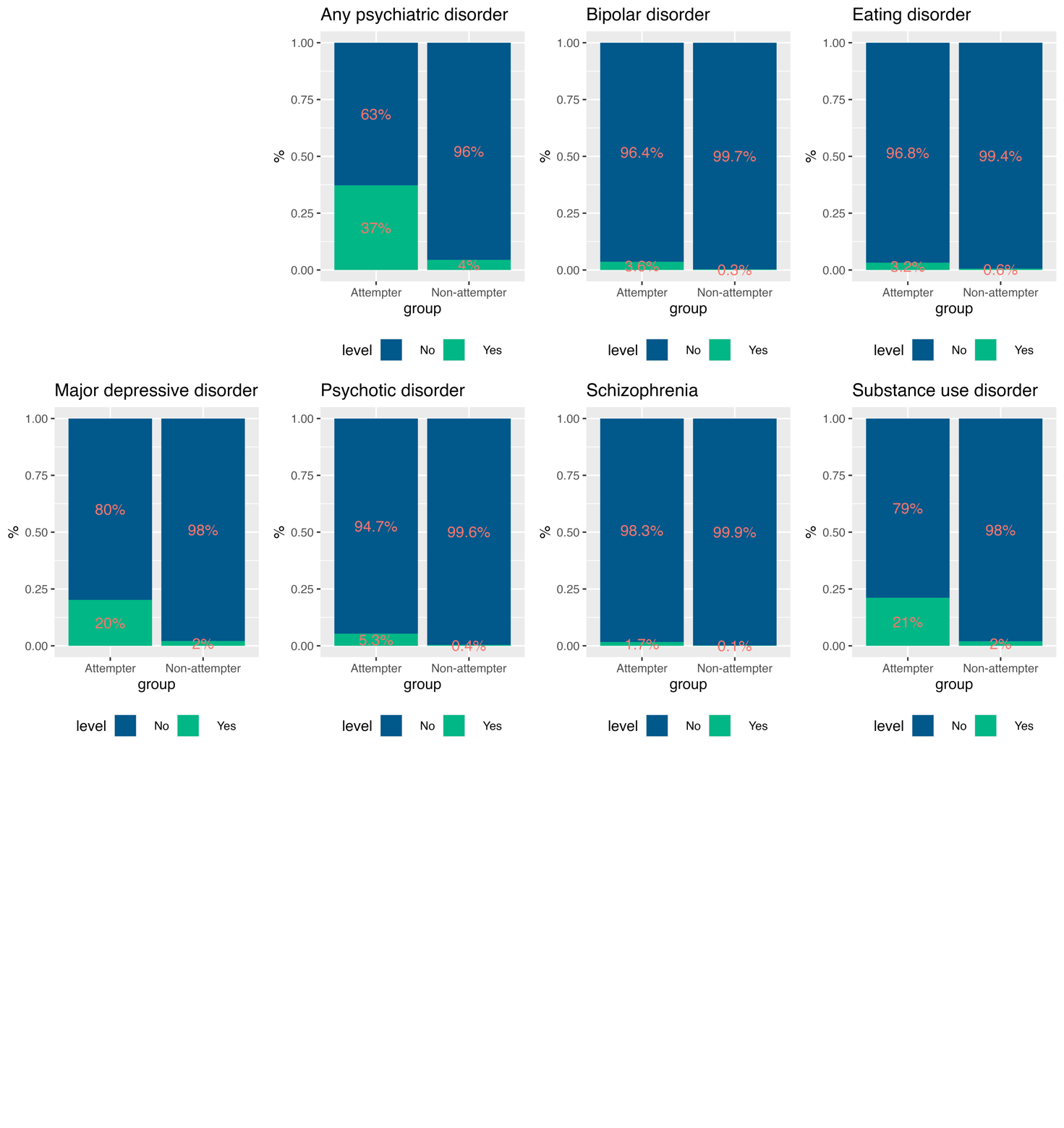

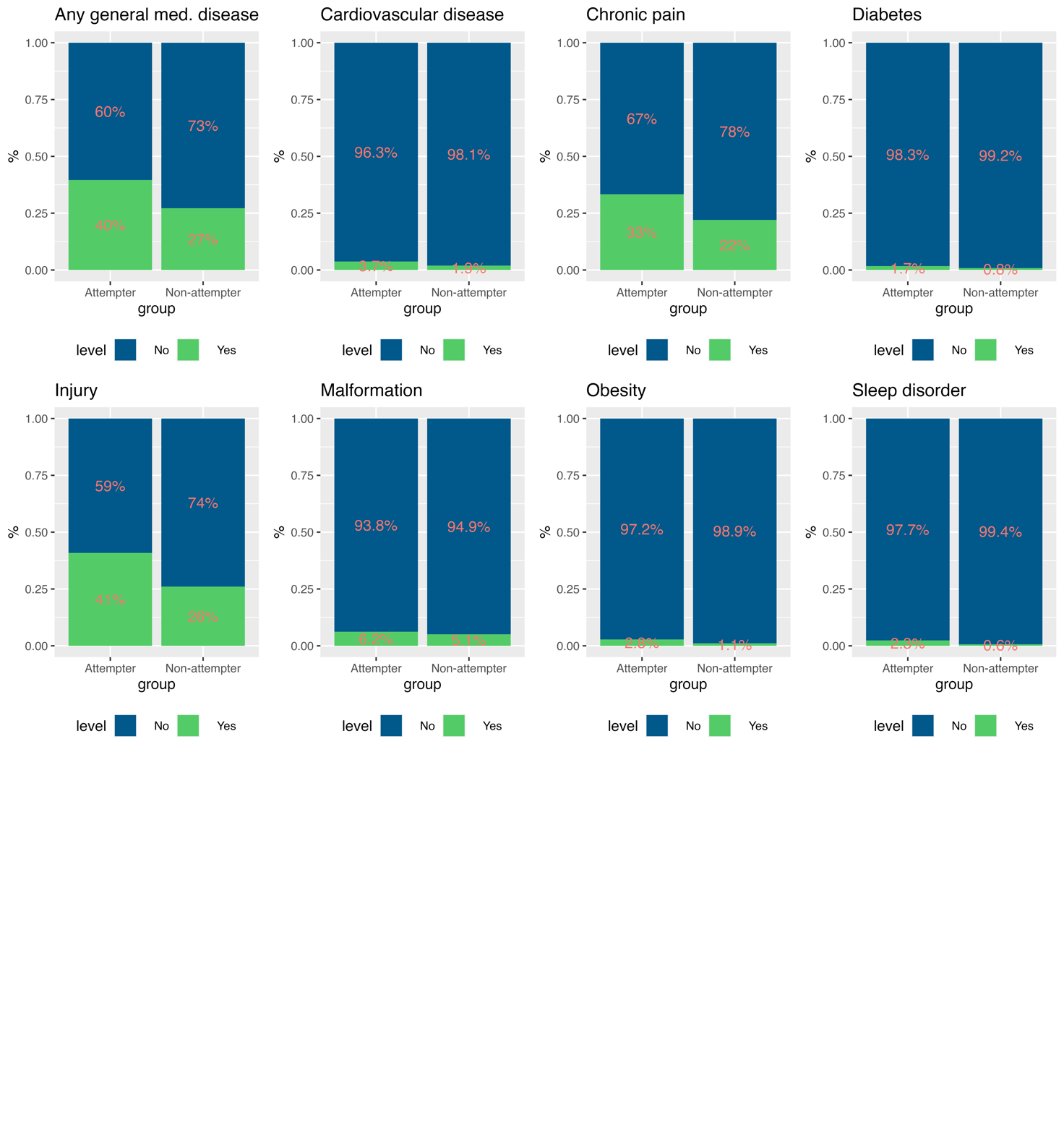

**Figure S11 (cont.)**: Distribution of risk factors among individuals with and without a suicide attempt ** SUD: Substance use disorder*

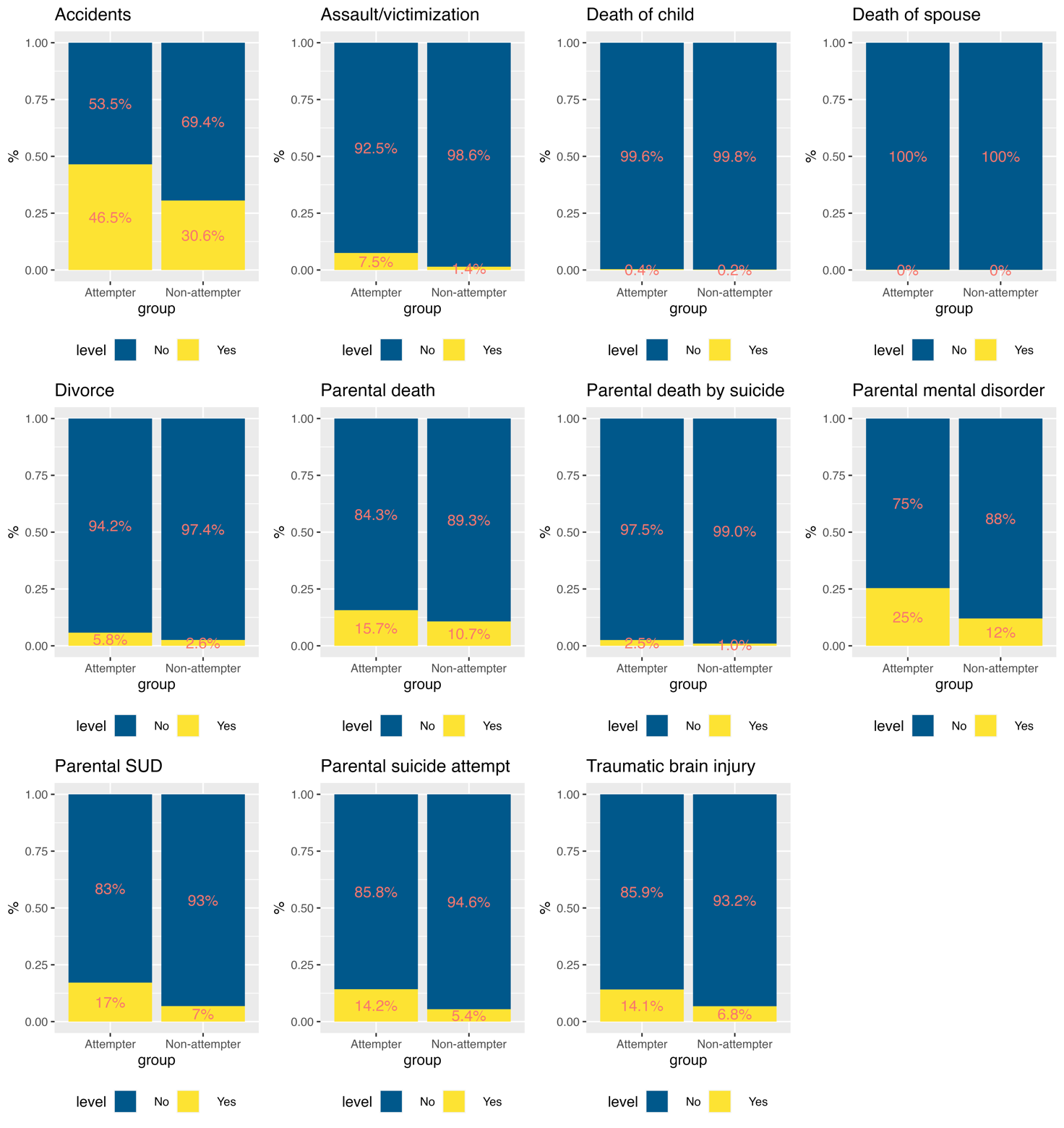

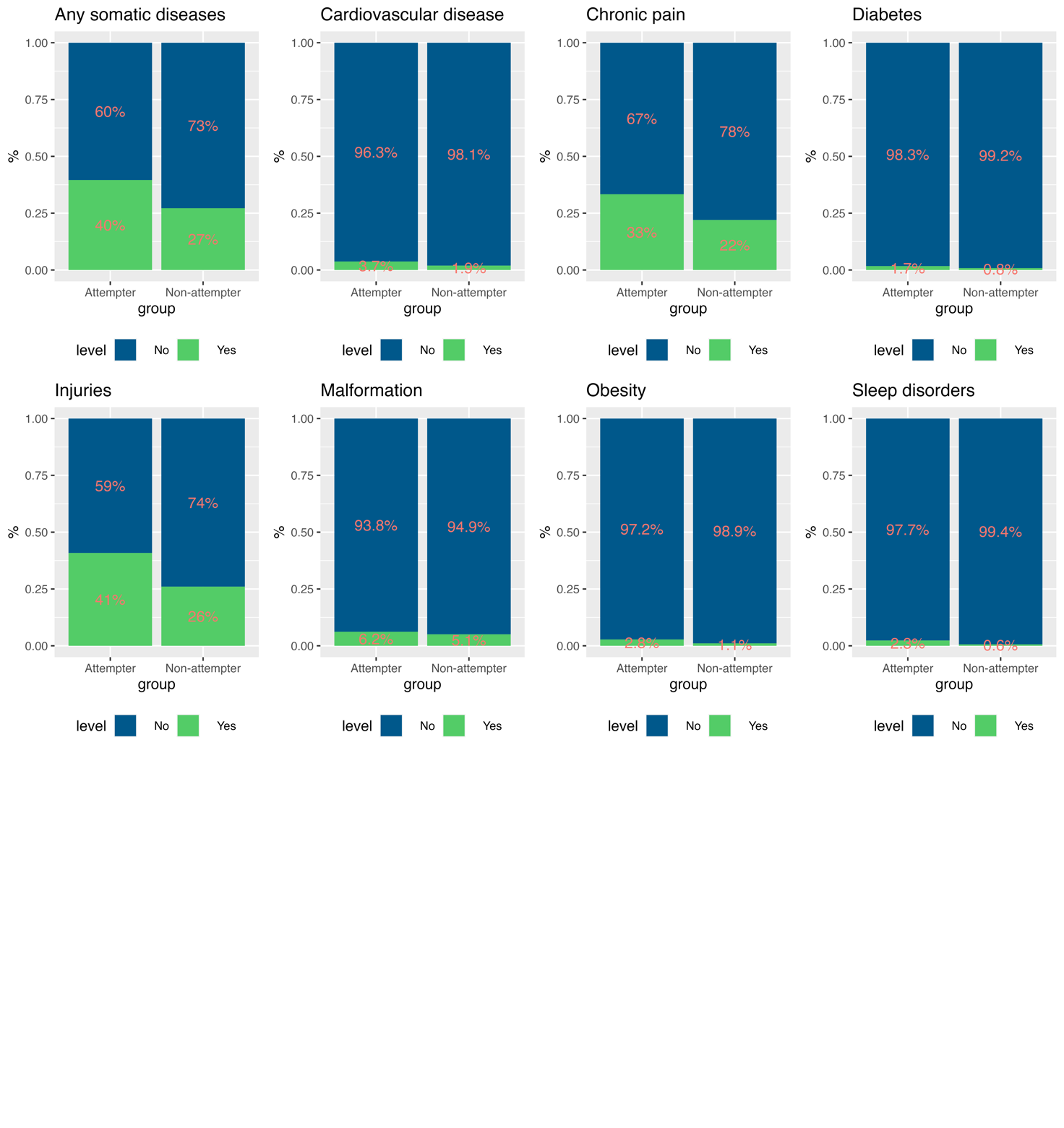

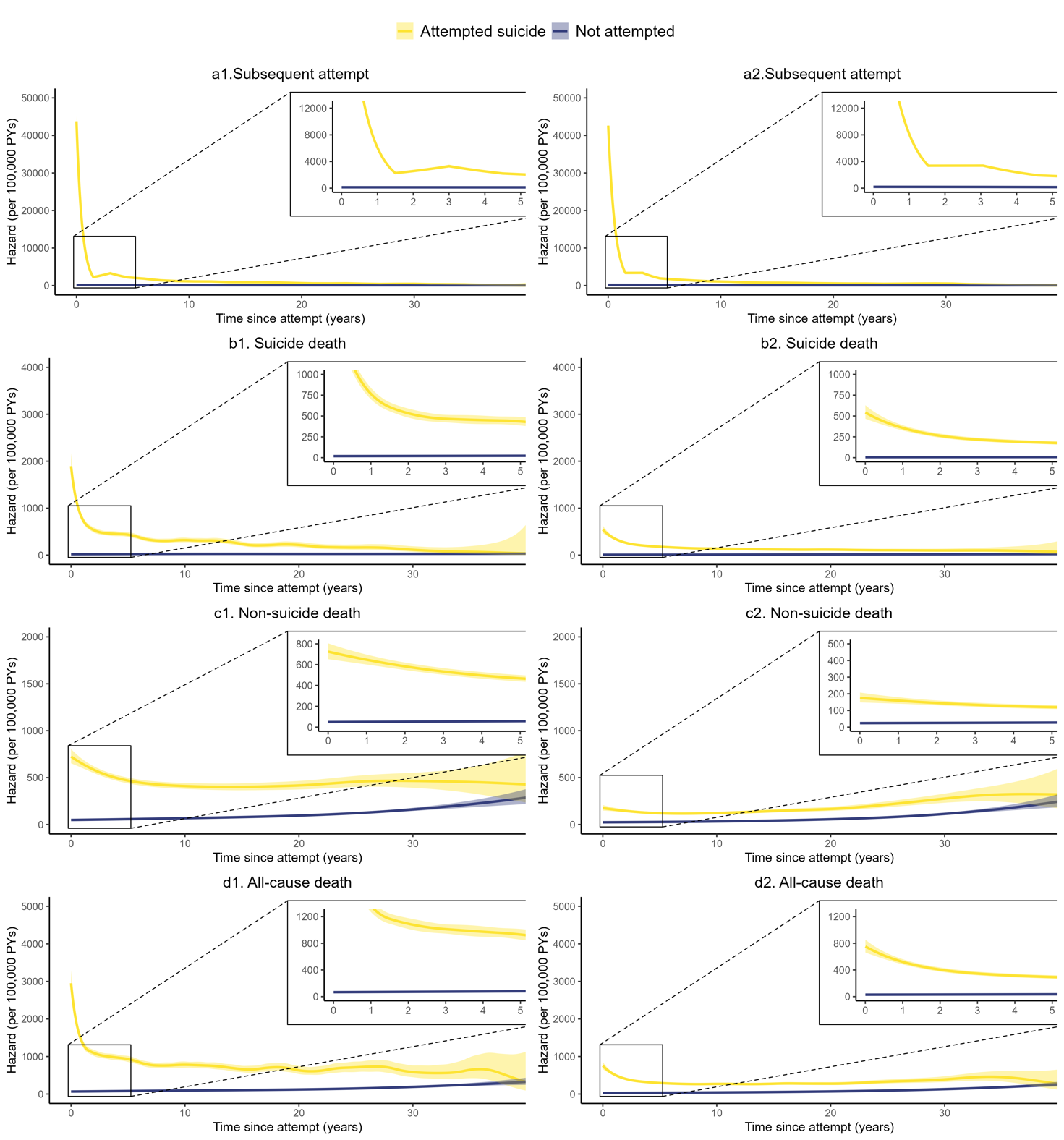

#### **Figure S12: Hazards of subsequent suicide attempt and mortality separately for males (a1-d1) and females (a2-d2)**

*The lines show hazards (number of events per 100,000 person-years) of four outcomes among individuals who had and had not an initial suicide attempt by sex from a matched cohort. Shaded areas represent 95% CI of the hazard.*

a. **Suicide attempt**

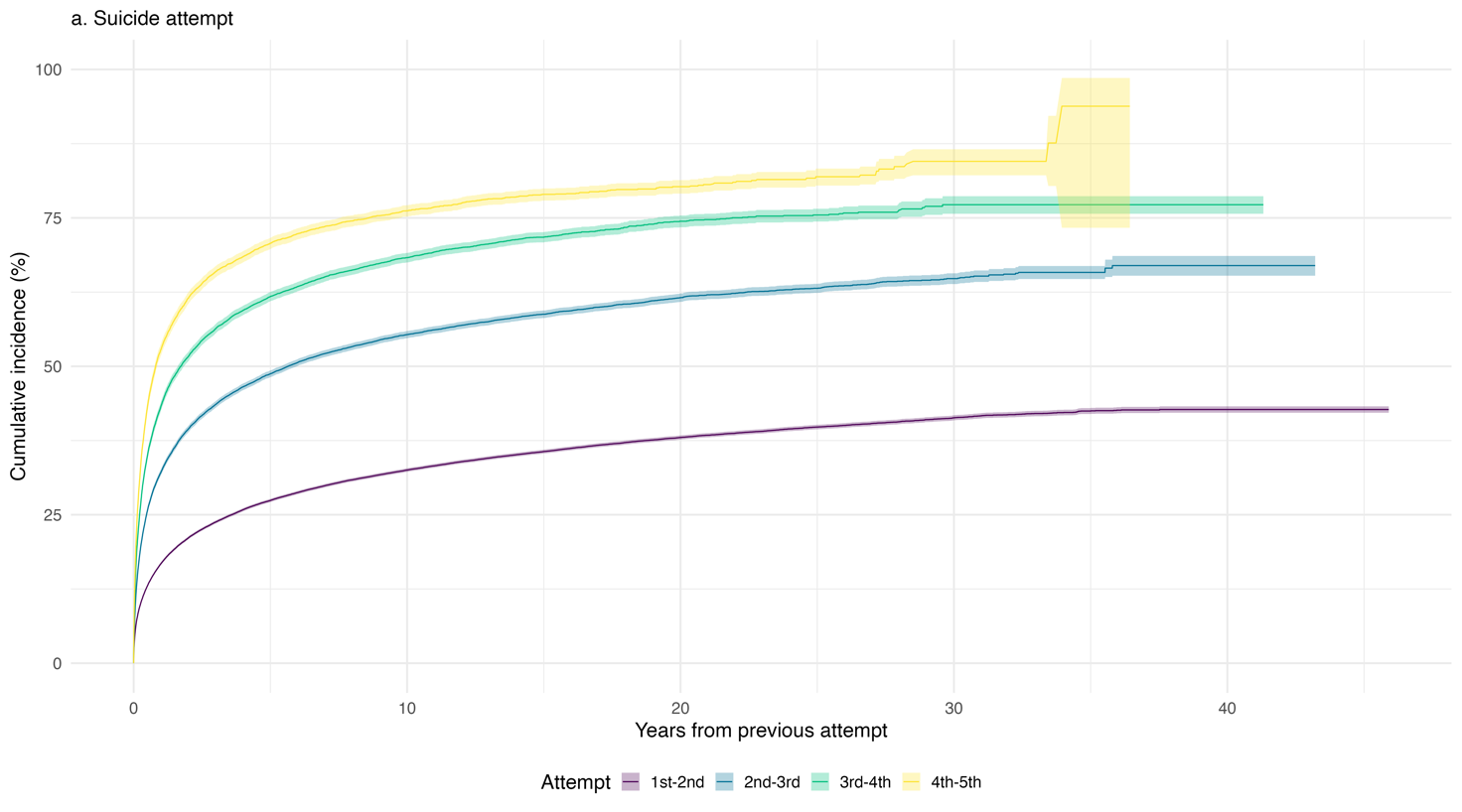

b. **Self-harm**

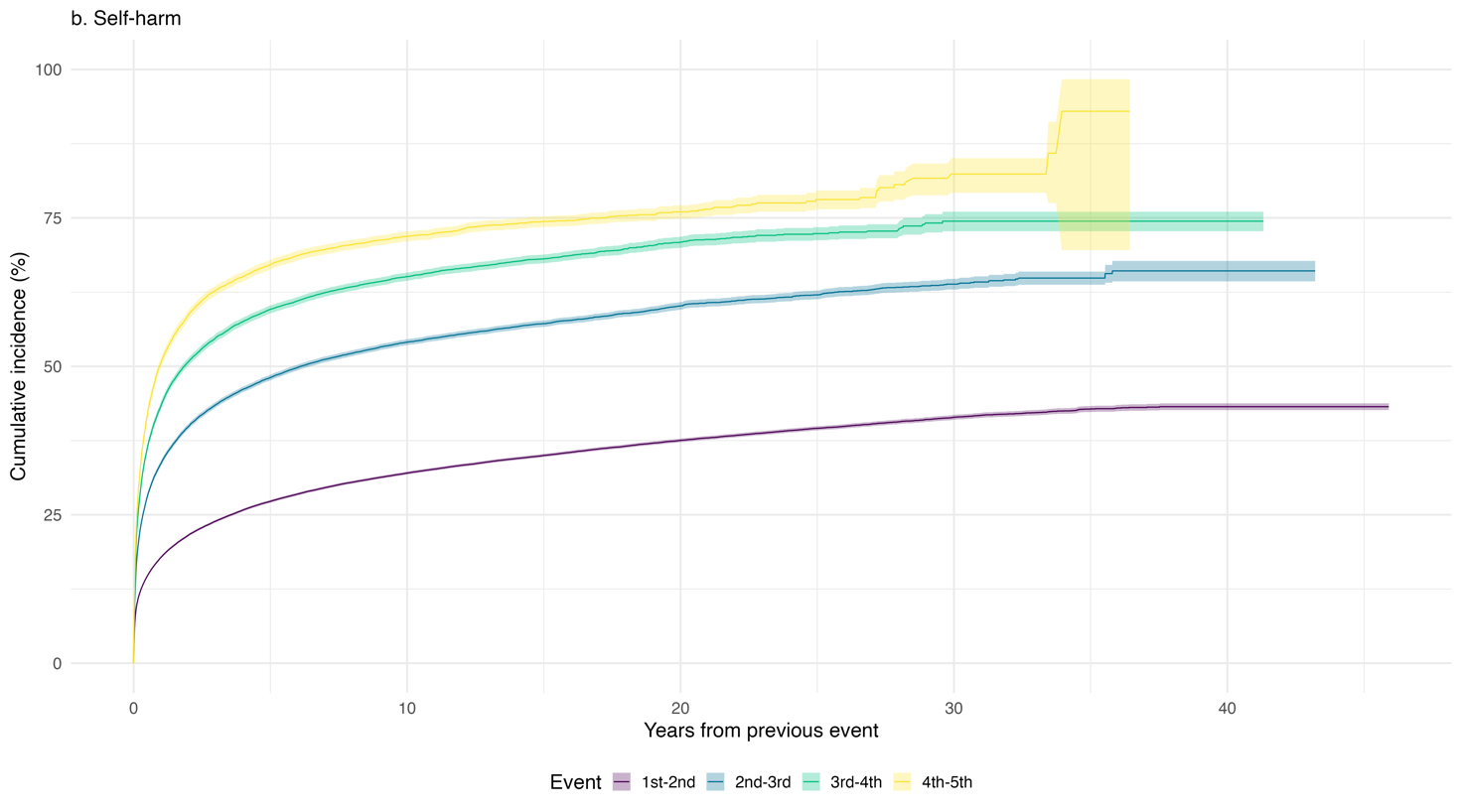

#### **Figure S13: Cumulative incidence of subsequent suicide attempts**

*Data for* ***a.*** ***Suicide attempt*** *and* ***b. Self-harm****. Cumulative incidence of having a subsequent suicide attempt/self-harm since the preceding attempt estimated using Kaplan-Meier method. Lines show the proportion of individuals who had a subsequent attempt/self-harm among those who had the preceding attempt (e.g., the lowest line represents the proportion of individuals attempted suicide/self-harm the 2^nd^ time among those with at least 1 attempt of suicide/self-harm). Shaded regions show 95% confidence interval of the cumulative incidence.*

a. **Suicide attempt**

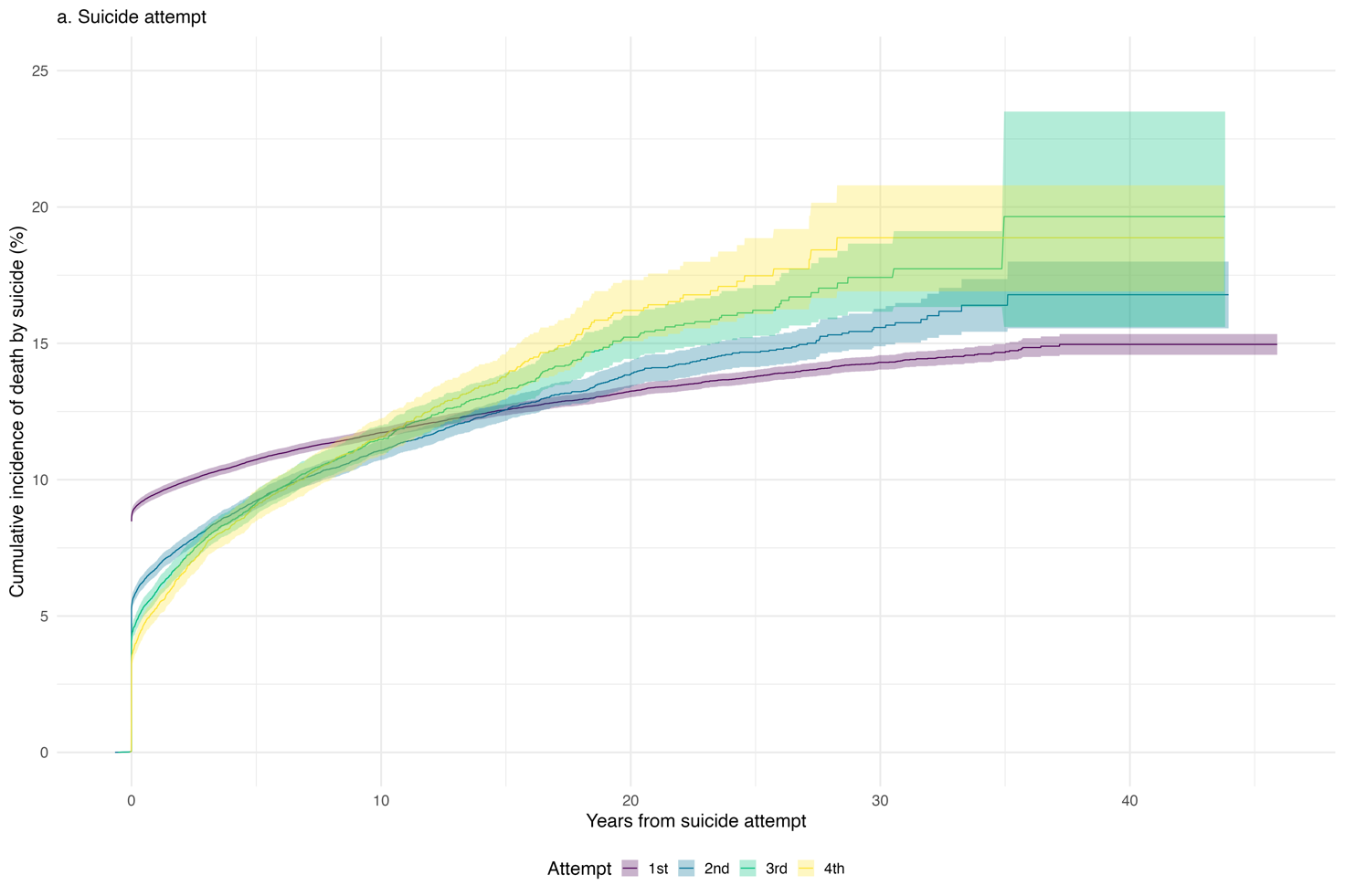

b. **Self-harm**

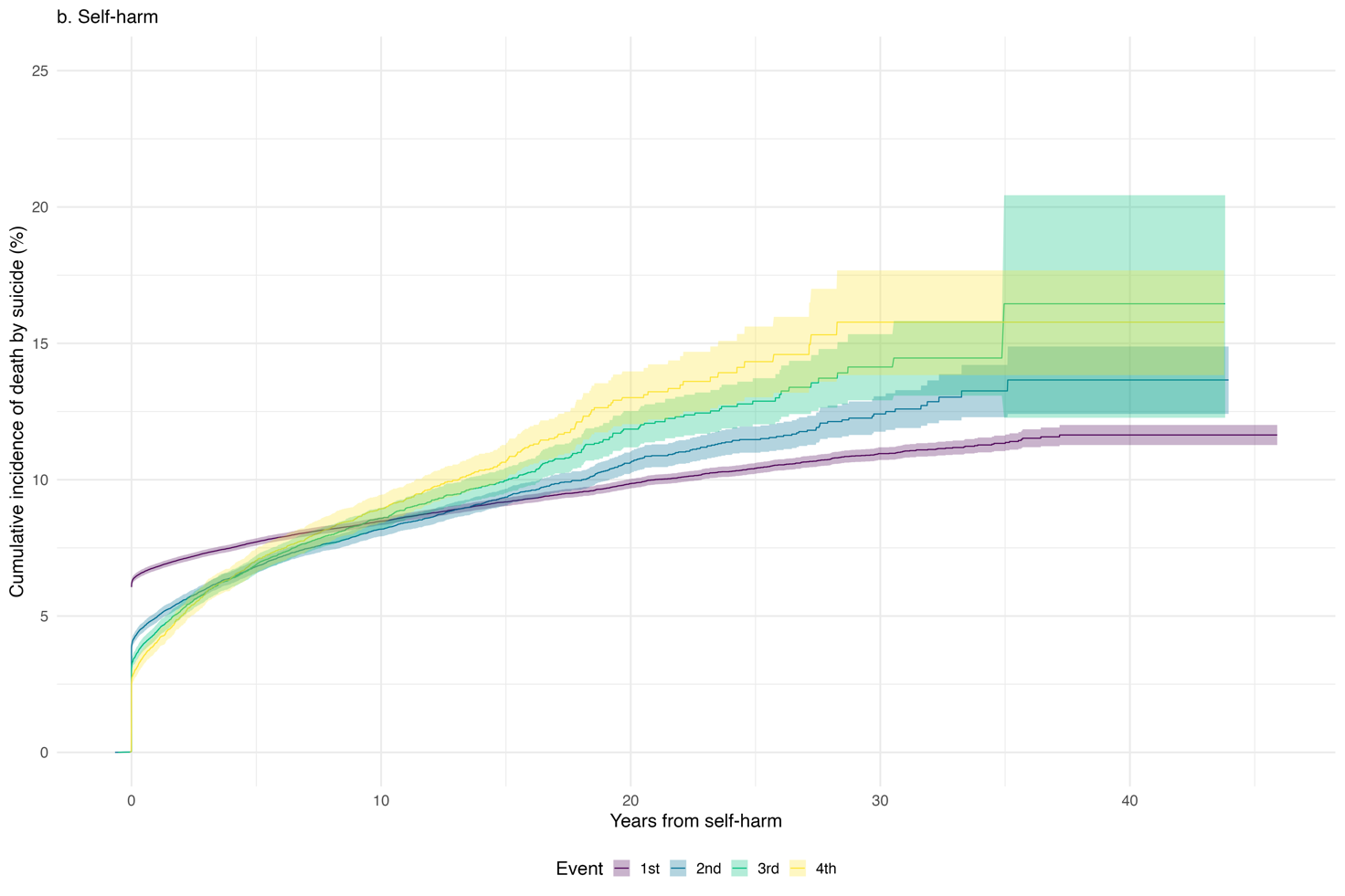

#### **Figure S14: Cumulative incidence of suicide mortality**

*Data for* ***a.*** ***Suicide attempt*** *and* ***b. Self-harm****. Cumulative incidence of suicide mortality since the suicide attempt/self-harm estimated using Kaplan-Meier method. Lines show the proportion of individuals who died by suicide since the 1^st^, 2^nd^, 3^rd^, and 4^th^ suicide attempt/self-harm. Shaded regions show 95% confidence interval of the cumulative incidence.*

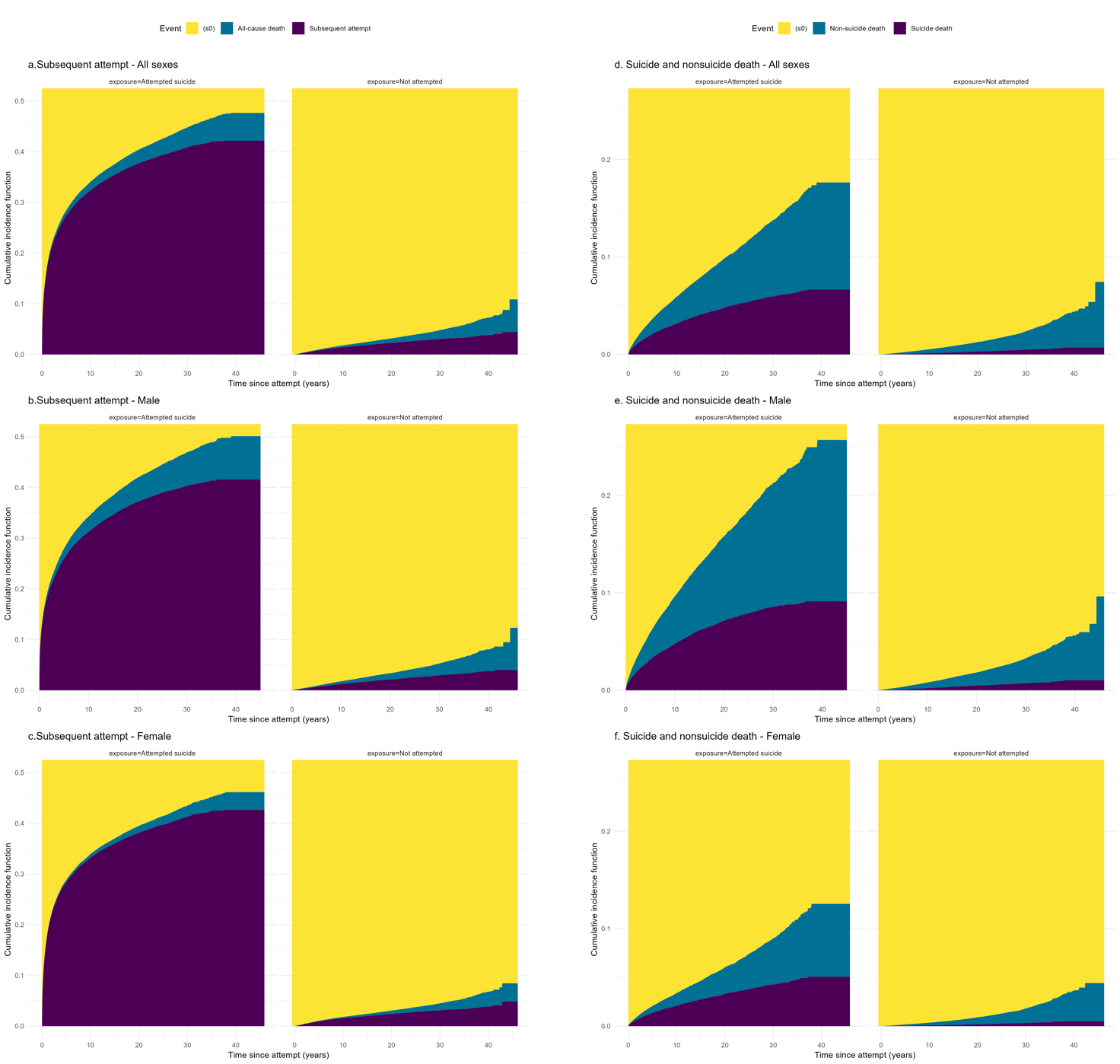

#### **Figure S15: Cumulative incidence of subsequent attempts and suicide mortality considering competing risk**

***a, b, c) Cumulative incidence of subsequent attempt considering all-cause mortality as competing risk for all population, males and females, respectively.***

***d, e, f) Cumulative incidence of suicide mortality considering non-suicide mortality as competing risk for all population, males and females, respectively.***

*Data for individuals with suicide attempts, estimated using Aalen-Johansen estimator. The lines show the proportion of individuals with suicide attempts having the outcome at each year after the initial attempt.*

4.	*A Package for Survival Analysis in R* [computer program]. Version survival-package2023.

5.	*Utility Functions, Datasets and Extended Examples for Survival Analysis* [computer program]. 2022.

16.	Gvion Y, Levi-Belz Y. Serious Suicide Attempts: Systematic Review of Psychological Risk Factors. *Frontiers in Psychiatry.* 2018;9.
